# Temporal associations between pain and anxiety/depression in young people: A systematic review and meta-analysis of longitudinal, prospective, population- and community-based studies

**DOI:** 10.64898/2026.10.20.26366024

**Authors:** Thi Viet Hai Tran, Rebecca P. Harding, Annabel Rowney-Smith, Beril Can, Joshua Reynolds, Faith Matcham, Kathryn J. Lester, Ellen J. Thompson

**Author notes:** **Corresponding author:** Ellen J. Thompson. **Email address:** Thi Viet Hai Tran, Rebecca P. Harding, Annabel Rowney-Smith, Beril Can, Joshua Reynolds, Faith Matcham, Kathryn J. Lester.

## Abstract

Pain, anxiety, and depression are common among young people, frequently co-occur with each other, and may be longitudinally related. This systematic review and meta-analysis aimed to synthesise the direction and strength of the temporal associations between pain and anxiety/depression among young people. PsycINFO, Web of Science, Scopus, PubMed, Proquest Dissertations and Theses, Google Scholar, and OSF Preprints were searched from inception until 3 July 2026. Additional studies were identified through citation searching and references of existing systematic reviews. Longitudinal, prospective, population-or community-based studies examining temporal associations between pain and anxiety/depression among young people (0-24 years) were included. Two independent reviewers screened studies, extracted data, assessed risk of bias using ROBINS-E, and evaluated certainty of evidence using GRADE. Data were synthesised using three-level meta-analysis and narrative synthesis. Forty-two studies from 31 longitudinal cohorts were included. Anxiety, depression, and combined anxiety/depression were positively associated with subsequent pain. However, after adjustment for confounders, only depression remained significantly associated. Pain was positively associated with subsequent anxiety, depression, and combined anxiety/depression both before and after confounder adjustment. Heterogeneity was moderate to considerable and was partly explained by sex, baseline age, follow-up duration, pain type, pain informant, pain and anxiety/depression operationalisation, and risk of bias. Two of 14 outcomes had high quality of evidence, whereas the remainder ranged from very low to moderate quality. These findings highlight longitudinal associations between pain and anxiety/depression among young people and should be considered in the screening, treatment, and public health management of these problems.

## 1. Introduction

Pain is common in young people, with current estimates of up to 20% reporting chronic pain [18] and up to 93% reporting acute pain [105]. Anxiety and depression are also common, affecting 1.32-4.58% and 0.08-3.85% young people respectively [57]. Pain and anxiety/depression are among the leading causes of disability and can result in significant distress, poorer quality of life, and a broad range of negative physical, psychological, and social outcomes [22,35,118,132].

Pain frequently co-occurs with anxiety and depression. Approximately one in three young people (< 25 years) experiences chronic pain and anxiety, and one in eight live with both pain and depression [23]. Among youths with chronic pain, anxiety and depression show moderate-to-large associations with greater disability, pain interference, pain catastrophising, and reduced quality of life [42]. Anxiety/depression can impede pain recovery, whilst pain may hinder identification of anxiety/depression, leading to undertreatment [131]. Therefore, understanding the co-occurrences of pain and anxiety/depression is important for developing prevention strategies and interventions for those at risk.

Longitudinal studies have suggested temporal associations between pain and anxiety/depression, but findings remain mixed. Whilst some studies found that pain is associated with subsequent anxiety/depression symptoms or disorders in young people [48,83,112], others reported the reverse [7,68,98]. Several studies found no associations [31,33,107]. These inconsistencies highlight the need to synthesise the available evidence to clarify the strength and direction of these associations.

Several syntheses have been conducted, but important gaps remain. Many have examined anxiety/depression as risk factors for pain and found that they increase the risk of pain onset and persistence in young people [19,45,49–51,54]. However, fewer studies have examined whether pain is associated with subsequent anxiety/depression. To our knowledge, only one review investigated the prevalence of anxiety and depression in youths with chronic pain [23], and none evaluated the association. Although other reviews have examined general associations between pain and anxiety/depression in young people [10,15,27,94], no temporality could be established due to the inclusion of cross-sectional studies.

In addition, existing reviews have included studies using retrospective measures, which are vulnerable to recall bias and reduced reliability [125], especially when children recall past experiences [20]. Furthermore, they have synthesised evidence from both clinical and population-and/or community-based studies, despite evidence showing that the strength of the association between pain and anxiety/depression may vary across settings [23]. Given that pain, anxiety, and depression are major public health issues [24,74], a synthesis of longitudinal, prospective studies with population-and community-based samples is warranted to better understand their development in the broader population of young people.

Overall, this systematic review and meta-analysis aimed to investigate the direction and strength of the temporal associations between pain and anxiety/depression. Following the most recent commissioned report on paediatric pain [24], we focused on young people from birth to age 24. We synthesised evidence exclusively from longitudinal, prospective, population-and community-based studies. An improved understanding of this relationship can ultimately guide future prevention and treatment strategies for pain, anxiety, and depression in young people.

## 2. Methods

The study protocol was prospectively registered with Prospero (CRD420251018369). This systematic review and meta-analysis adhered to the Preferred Reporting Items for Systematic Review and Meta-analysis (PRISMA 2020) [86] and to the Meta-analyses of Observational Studies in Epidemiology (MOOSE) [116].

### 2.1. Search strategy

Published and unpublished literature was searched using Proquest-interface of PsycINFO, Web of Science, Scopus, and PubMed from inception until 3 July 2026. These databases were chosen as they provided the greatest access to relevant publications in the topic area. The search included terms for pain, anxiety, depression, longitudinal prospective design, and young people. Full search strategies for each database are in Supplementary Material. Grey literature was also searched from Proquest Dissertations and Theses, Google Scholar, and OSF Preprints from inception until 3 July 2026, to increase the review’s comprehensiveness, timeliness, and reduce publication bias. The same search strategy was used for grey literature. For Google Scholar, the first 300 results were sorted by relevance and selected as recommended by Haddaway and colleagues [38]. We also did backwards and forward citation searching using Web of Science and Scopus, following TARCiS recommendations [47]. References of relevant systematic reviews were also hand-searched for additional eligible studies.

### 2.2. Eligibility criteria

#### 2.2.1. Source

Only articles written in English were included.

#### 2.2.2. Population

Studies with human participants aged 0-24 years were eligible. Studies including participants older than 24 were included only if separate analyses were reported for participants aged 0-24. We included studies that recruited participants from the general population or the community and imposed no geographical restrictions. Studies recruiting participants from clinical (e.g., clinics, hospitals, primary care) or other special populations (e.g., youths with chronic pain or depressive disorders at baseline) were excluded. Studies that focused exclusively on pain or anxiety/depression associated with specific conditions (e.g., pregnancy, cancer) or circumstances (e.g., war) were also excluded.

#### 2.2.3. Phenotypes

##### Pain

Studies examining acute, chronic, or general pain conditions or symptoms were eligible. Pain must be assessed prospectively. Studies based solely on retrospective recall or historical records (e.g., insurance claims or health records), or involving experimentally induced pain were excluded.

##### Depression and anxiety

Anxiety and depression symptoms or disorders must be assessed prospectively using psychometrically validated measures (e.g., a clinical structured interview or validated questionnaires). Where a cut-off point was used, it must have been psychometrically validated. Studies using non-validated measures/cut-off points or solely retrospective measures were excluded.

#### 2.2.4. Design

Longitudinal studies examining unidirectional and/or bidirectional temporal associations between pain and anxiety/depression in young people were eligible. By temporal, measures of pain and anxiety/depression were assessed at two or more distinct time points. No restrictions were placed on the intervals between assessments. Studies must report effect sizes or sufficient quantitative or descriptive data to infer the direction and strength of the associations. Studies without original empirical data (e.g., reviews, meta-analyses, commentaries, editorials), qualitative studies, cross-sectional studies, and studies lacking sufficient data were excluded.

When multiple studies were found for the same cohort, all studies using different waves were included. When studies used datasets from the same waves but with different sample sizes, the study with the largest sample size was selected. An exception was when studies’ sample sizes were not entirely overlapping with one another and/or when they addressed different research questions. In those circumstances, all studies were included. Where sample sizes were identical and the same research questions were addressed, the study with the most robust methodology was included.

### 2.3. Study selection process

Search results were imported to EndNote 21 [119]. Duplicates were removed following Falconer [26]. When a study was reported in both a thesis/dissertation and a peer-reviewed publication, in line with Cochrane recommendations [63], information from both reports was collated, and the thesis/dissertation was selected because it had the most complete information. Titles were screened by the lead author (TVHT) in EndNote, with 10% independently double-screened by a second reviewer (RPH/JR). Abstracts were screened in the same manner using Rayyan [85]. All full texts were independently screened by TVHT and ARS/JR using Rayyan. Interrater reliability was assessed via Cohen’s Kappa [77]. Discrepancies were resolved through discussion and, where necessary, consultation with a third reviewer (EJT). Study authors were contacted when additional information was required to determine eligibility.

### 2.4. Data extraction

Data extraction was conducted using a standardised spreadsheet. A full list of extracted information is in Supplementary Material. Pilot testing of the data extraction spreadsheet was performed on a subset of five studies to refine the process and ensure comprehensiveness. Two reviewers (TVTH & BC/JR) independently extracted data from eligible studies to ensure consistency. Study authors were contacted twice, two weeks apart, to request any missing data.

### 2.5. Risk of bias assessment

Risk of bias was assessed using ROBINS-E tool [44], a specialised tool for evaluating observational epidemiologic studies on seven domains of bias: confounding, exposure measurement, outcome measurement, selection, attrition, presence of post-exposure interventions, and reporting. Where a paper investigated more than one relevant association, each association was assessed separately. Ratings included low, some concerns, high, or very high risk of bias. Two reviewers (TVHT & BC/JR) independently applied the tool to each included study. Any discrepancies were discussed and, if necessary, resolved with a third reviewer (EJT).

### 2.6. Data synthesis

#### 2.6.1. Statistical analyses

Three-level meta-analyses were conducted to examine the direction and strength of the temporal associations between pain and anxiety/depression in young people. Six meta-analyses were performed: three between anxiety, depression, combined anxiety/depression separately with subsequent pain, and three between pain with anxiety, depression, and combined anxiety/depression separately. Only studies that reported effect sizes, or sufficient data to calculate them, were included. A minimum of two effect sizes from at least two studies was required for each meta-analysis. Effects that were not included in the meta-analysis were synthesised narratively.

Meta analyses were conducted using the {metafor} package [124] in R 4.5.2 [96]. Three-level models were used to account for dependencies among multiple effect sizes from the same study [40]. Heterogeneity variance was estimated using restricted maximum likelihood estimator (REML) [123]. Confidence intervals and significance tests were based on t-distributions to reduce risk of inflated Type I error rates [124].

Unadjusted and adjusted results were synthesised separately. For unadjusted analyses, all effects were converted to odds ratio because it was the most commonly reported measure [92]. For adjusted analyses, continuous and binary outcomes were meta-analysed separately using linear regression coefficients and odds ratios, respectively. All estimates were synthesised irrespective of the confounders included in individual studies. Result of meta-analyses were visually presented with forest plots.

Heterogeneity was assessed using within-study and between-study I^2^ statistic and interpreted according to Cochrane guidance (0-30%: might not be important; 30-50%: moderate; 50-75%: substantial; 75-100%: considerable) [78]. We also reported *τ̂*^2^ and 95% confidence intervals, which reflect uncertainty in heterogeneity estimates. In addition, 95% prediction intervals were reported to indicate the expected range of effects in future studies. Prediction intervals containing null value suggest greater uncertainty in the estimated pooled effect [40].

#### 2.6.2. Meta-regression

Meta-regressions were conducted to examine potential moderators of effect sizes within each three-level meta-analysis. We considered the following factors: sex, mean baseline age, age group (children/adolescents), pain type, pain operationalisation (binary/continuous), pain informant (youth/others), anxiety/depression operationalisation (binary/continuous), anxiety/depression informant (youth/others), follow-up duration, and risk of bias (low, some concerns, high/very high). Mean baseline age and follow-up duration was entered as continuous moderators, whilst others were entered as categorical moderators. Musculoskeletal (MSK) pain was defined according to the International Association for the Study of Pain and included back, limb, neck/shoulder, and general MSK pain [53]. Meta-regression was conducted only when at least two effects from at least two studies were available in at least two categories (or values) of a moderator. For pain types, due to insufficient effect sizes with pain types other than MSK pain, mixed pain, and headache, they were combined into one category. All meta-regression used the same dataset as the corresponding meta-analyses, except analyses of sex, which included only sex-specific associations.

### 2.7. Publication bias

To assess publication bias because of small-study effects, we visually inspected funnel plots and contour-enhanced funnel plots [40,90]. Contour-enhanced funnel plots were used to distinguish potential publication bias from other causes of funnel plot asymmetry. Specifically, gaps in the non-significance area suggest publication bias, whilst gaps in significance areas indicate other causes of asymmetry. Because inferences from funnel plots tend to be subjective [40], we also conducted Peters’ regression test [89]. Although Egger’s regression test [25] was initially chosen in the PROSPERO protocol, Peter’s test was selected because it accounts for situations where effect sizes and their standard errors are mathematically correlated, inflating false positive rates, as in the case of outcomes measured with odds ratios [89]. Evidence of publication bias due to small-study effects was inferred from significant Peters’ test and gaps in non-significance area of contour-enhanced funnel plot.

### 2.8. Certainty of evidence assessment

The GRADE (Grading of Recommendation, Assessment, Development, and Evaluation) framework [37] was used to assess the certainty of evidence for each outcome. Certainty was classified as high, moderate, low, or very low. Detailed GRADE rating criteria are provided in the Supplemental Material. Two reviewers (TVHT & EJT) independently assessed the overall certainty of evidence for each outcome. Discrepancies were resolved via discussion. GRADE ratings were integrated into the interpretation and conclusions of the review.

## 3. Results

### 3.1. Study selection

We identified a total of 35,552 records via databases and registers between inception and 3 July 2026. After duplicates removal, we screened 22,581 records, from which we reviewed 189 full-text documents and included 35 studies. Citation searching resulted in additional 6,382 records from which we reviewed 28 full-text documents and included seven studies. In total, 42 studies from 31 longitudinal cohorts were included in this review. Figure 1 shows the PRISMA diagram, which was created using PRISMA2020 Shiny App on R [38]. Interrater reliability for title/abstract screening and full-text screening was moderate (κ = 0.52 and 0.49 respectively), but the overall agreement rates were 85.78% and 80.19% respectively, reflecting the high proportion of excluded records at both screening stages.

**Figure 1.**
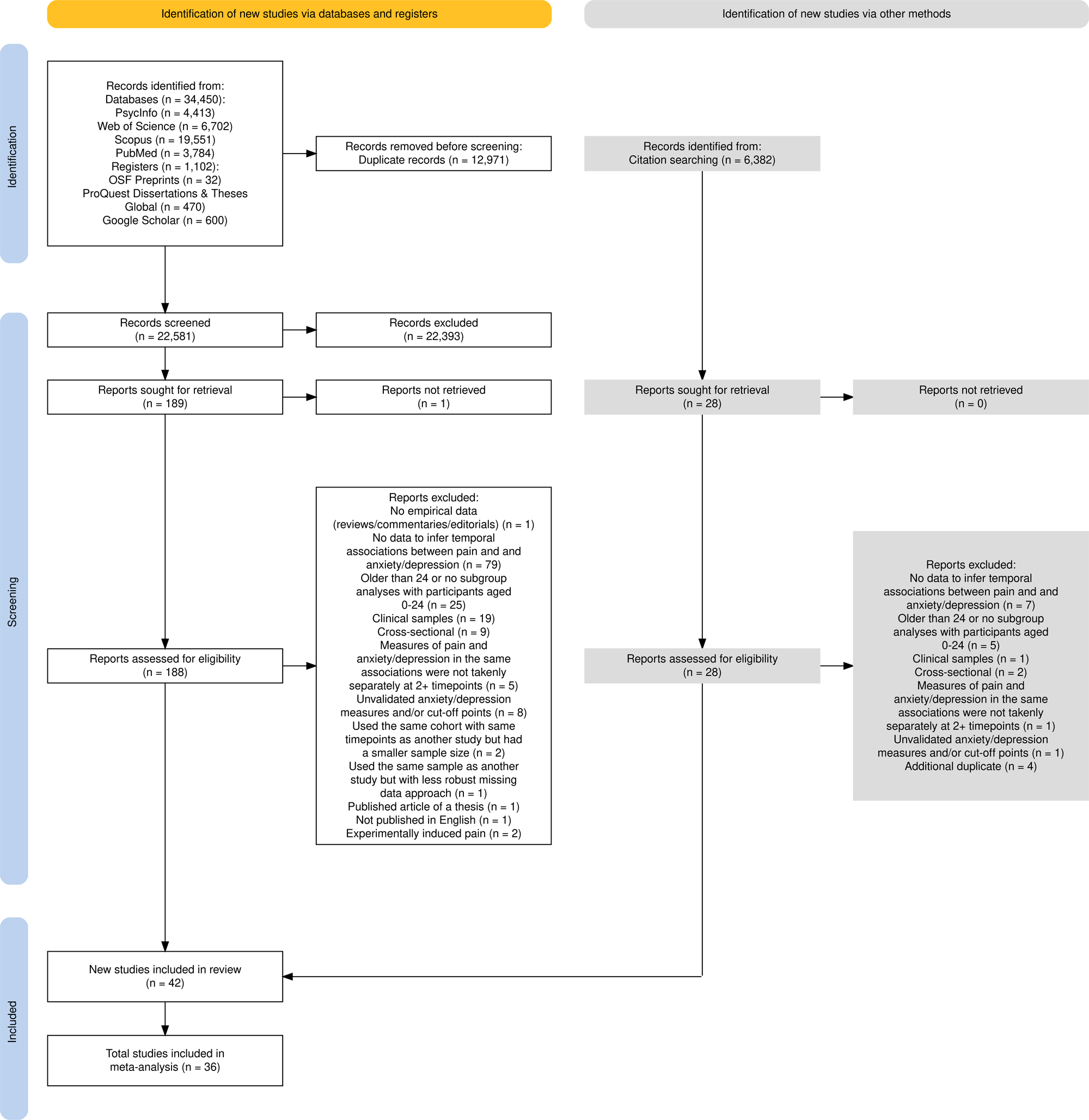
PRISMA 2020 flow diagram.

### 3.2. Study characteristics

Eligible studies were published between 2004 and 2026 (Table 1). Five studies investigated bidirectional associations between pain and anxiety/depression, and 37 examined unidirectional associations. Across studies, 14 examined anxiety and subsequent pain, 17 depression and subsequent pain, 14 anxiety/depression combined and subsequent pain, 10 pain and subsequent anxiety, 10 pain and subsequent depression, and six pain and subsequent anxiety/depression combined. Nearly half (42.3%) were females. Participants were only children (0-9) at baseline in nine studies, only adolescents (10-18) in 28 studies, only young adults (19-24) in one study, a mix of children and adolescents in three studies, and a mix of adolescents and young adults in one study. Follow-up durations ranged from three months to 15 years. Mixed pain was the most frequently studied (*k* = 15). Among specific pain types, musculoskeletal (MSK) pain was the most frequently examined (*k* = 14). There were 12 countries represented with most studies being conducted in Europe (*k* = 21), followed by North America (*k* = 12), Asia (*k* = 5), and Australia (*k* = 4).

**Table 1.**
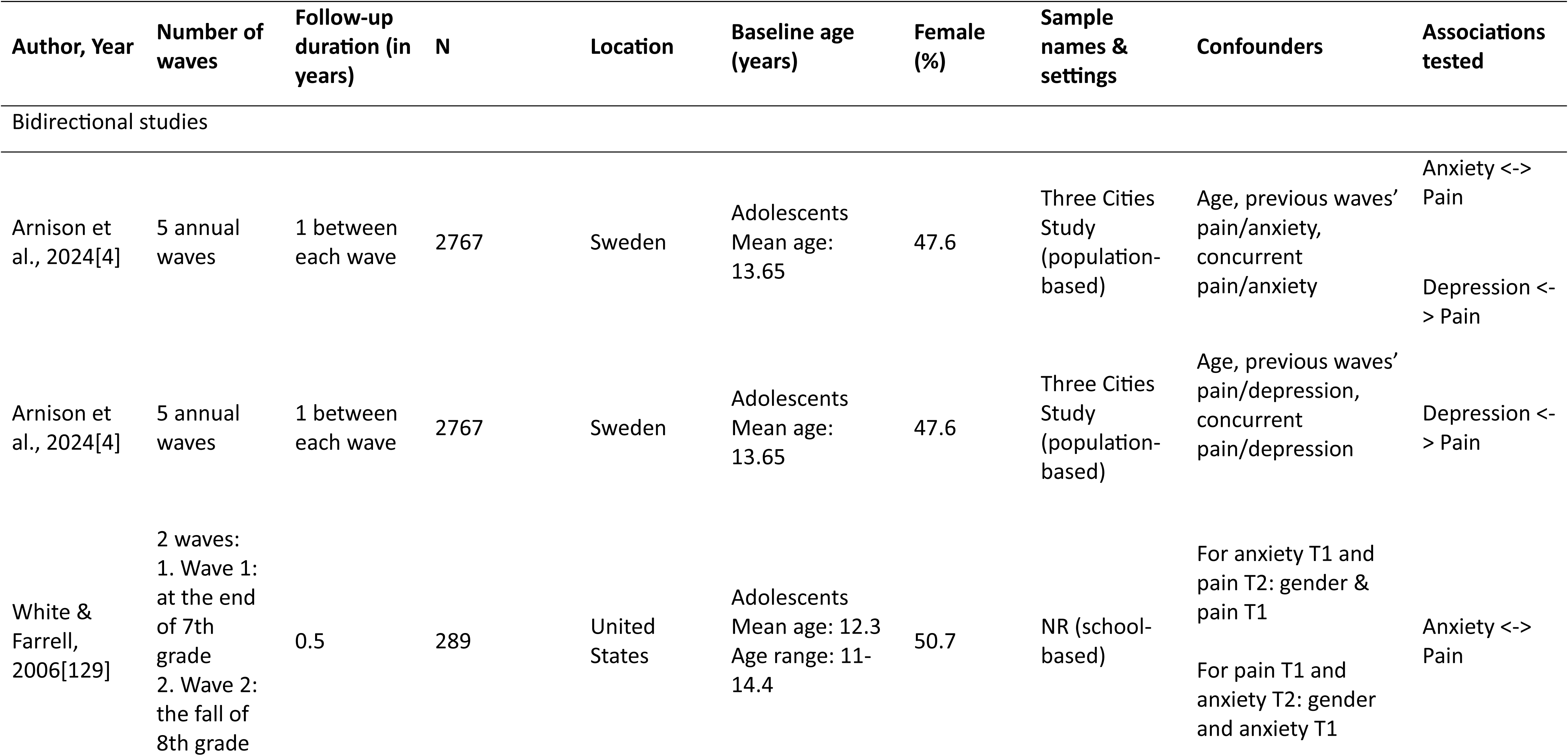
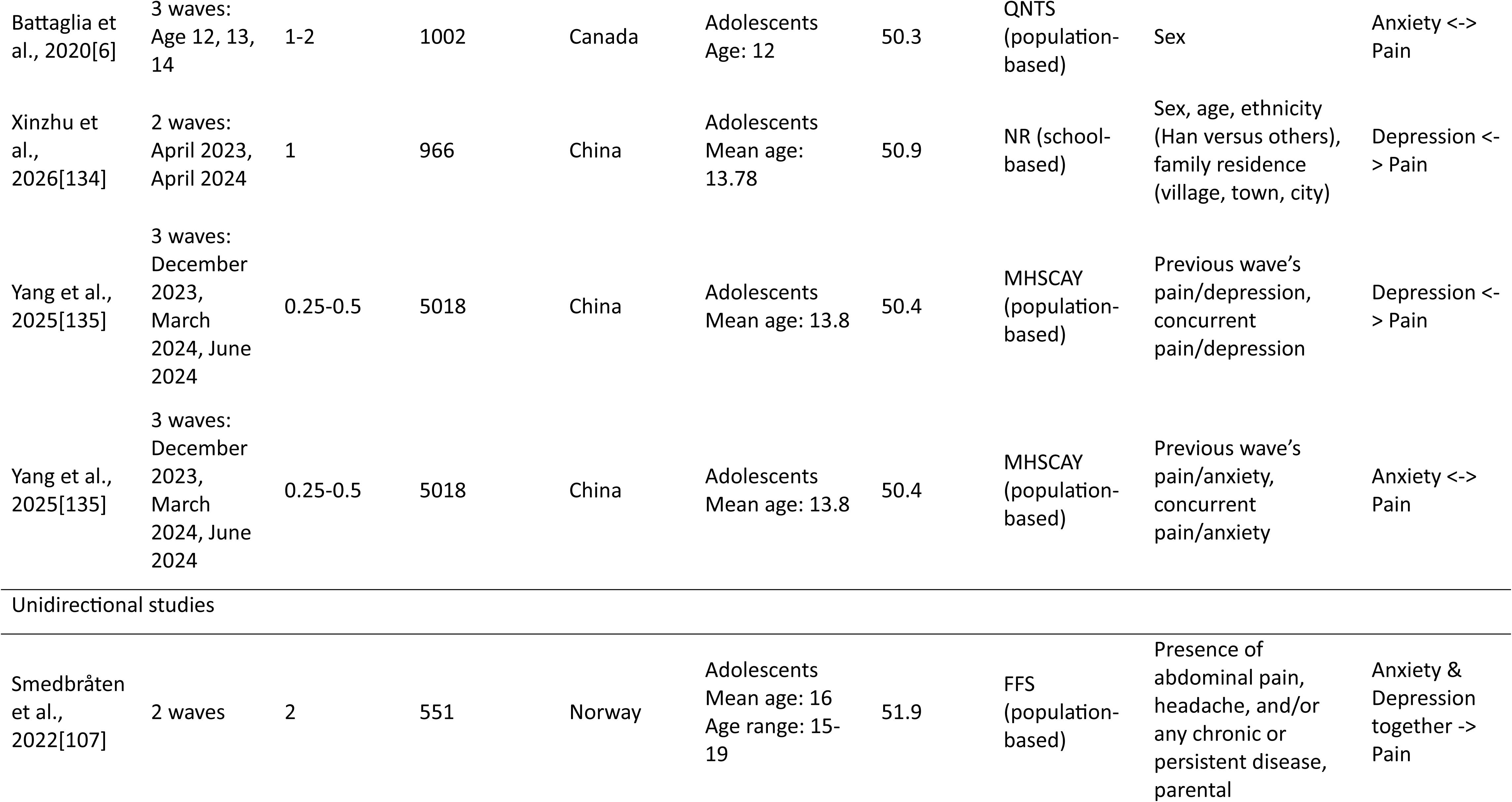
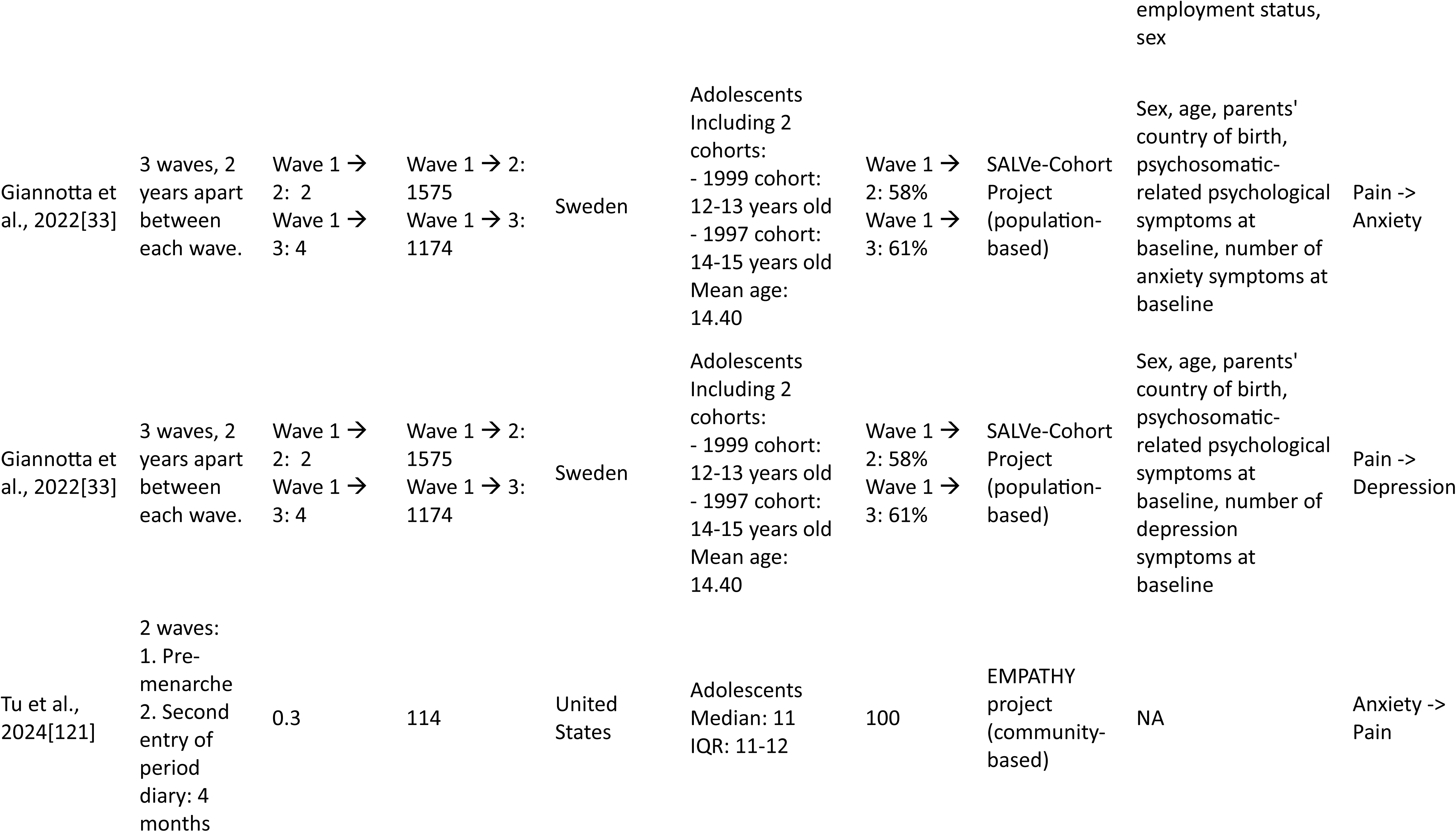
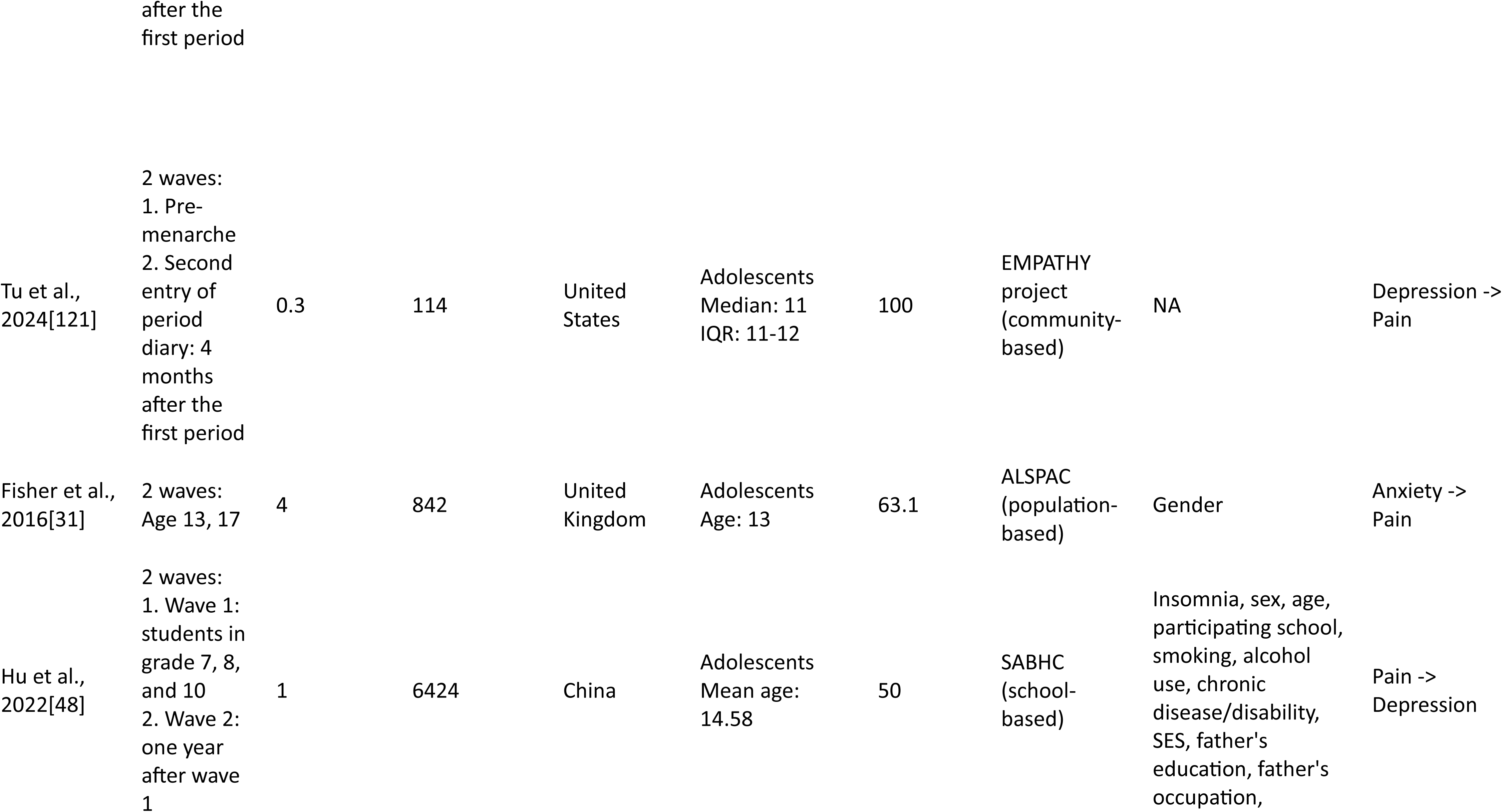
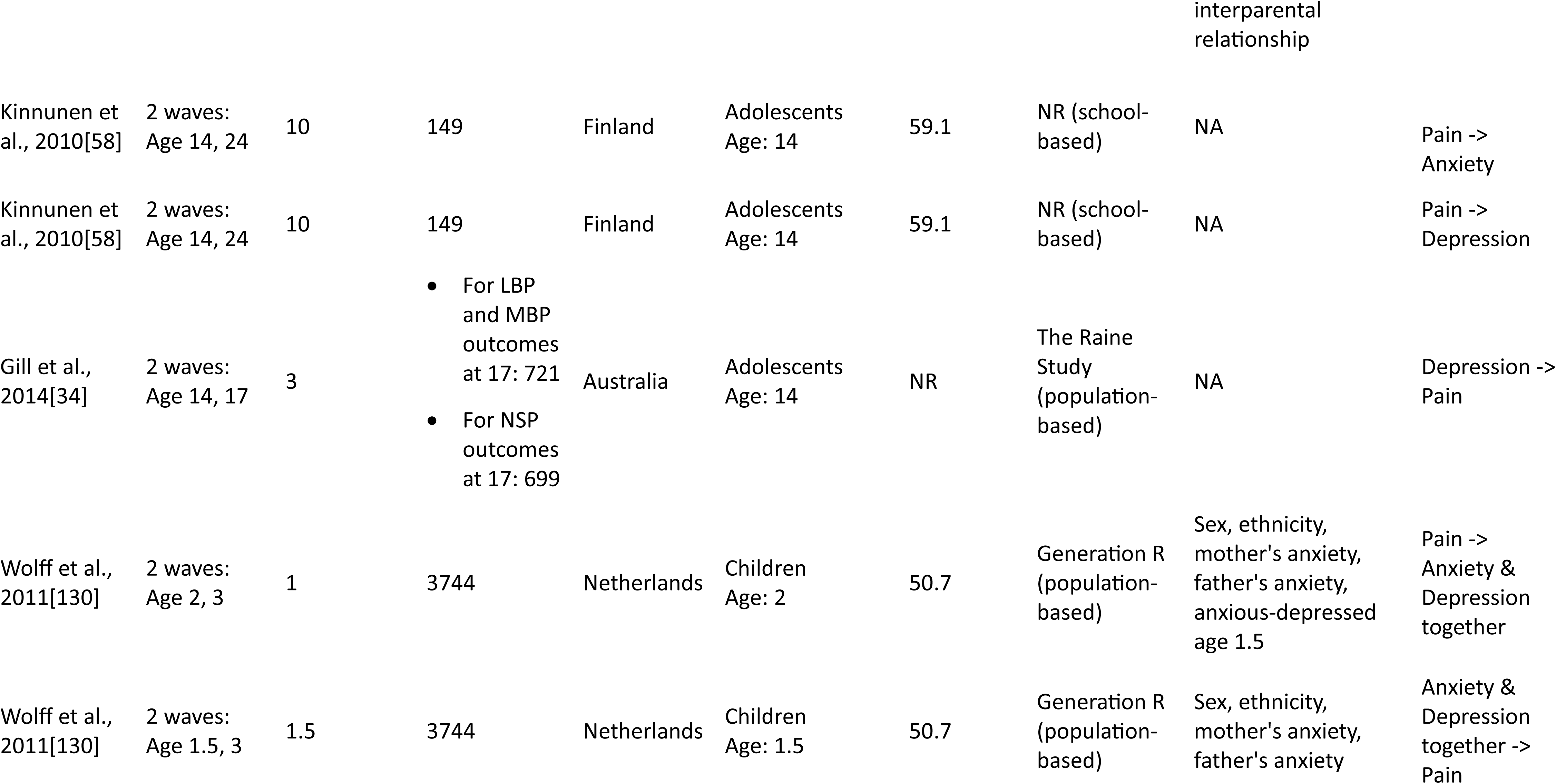
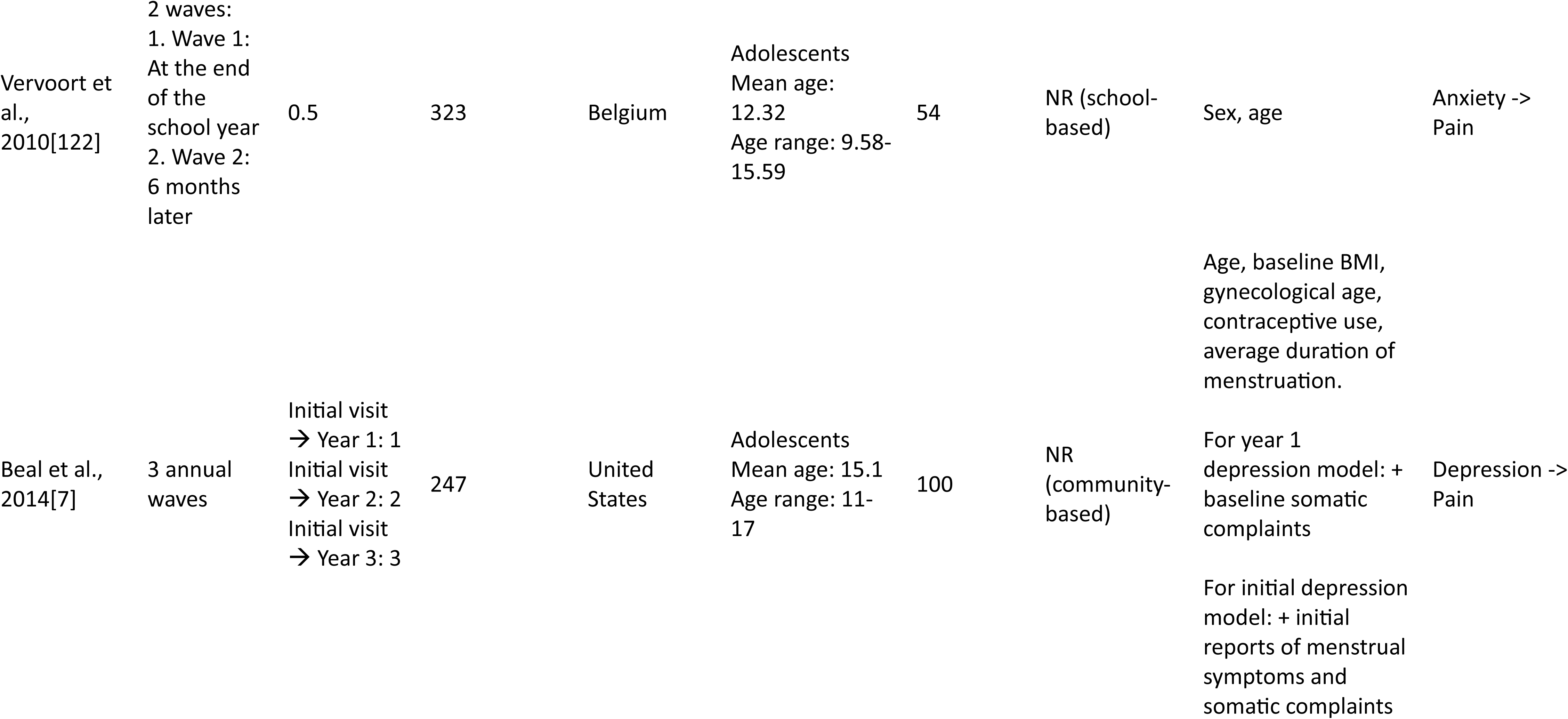
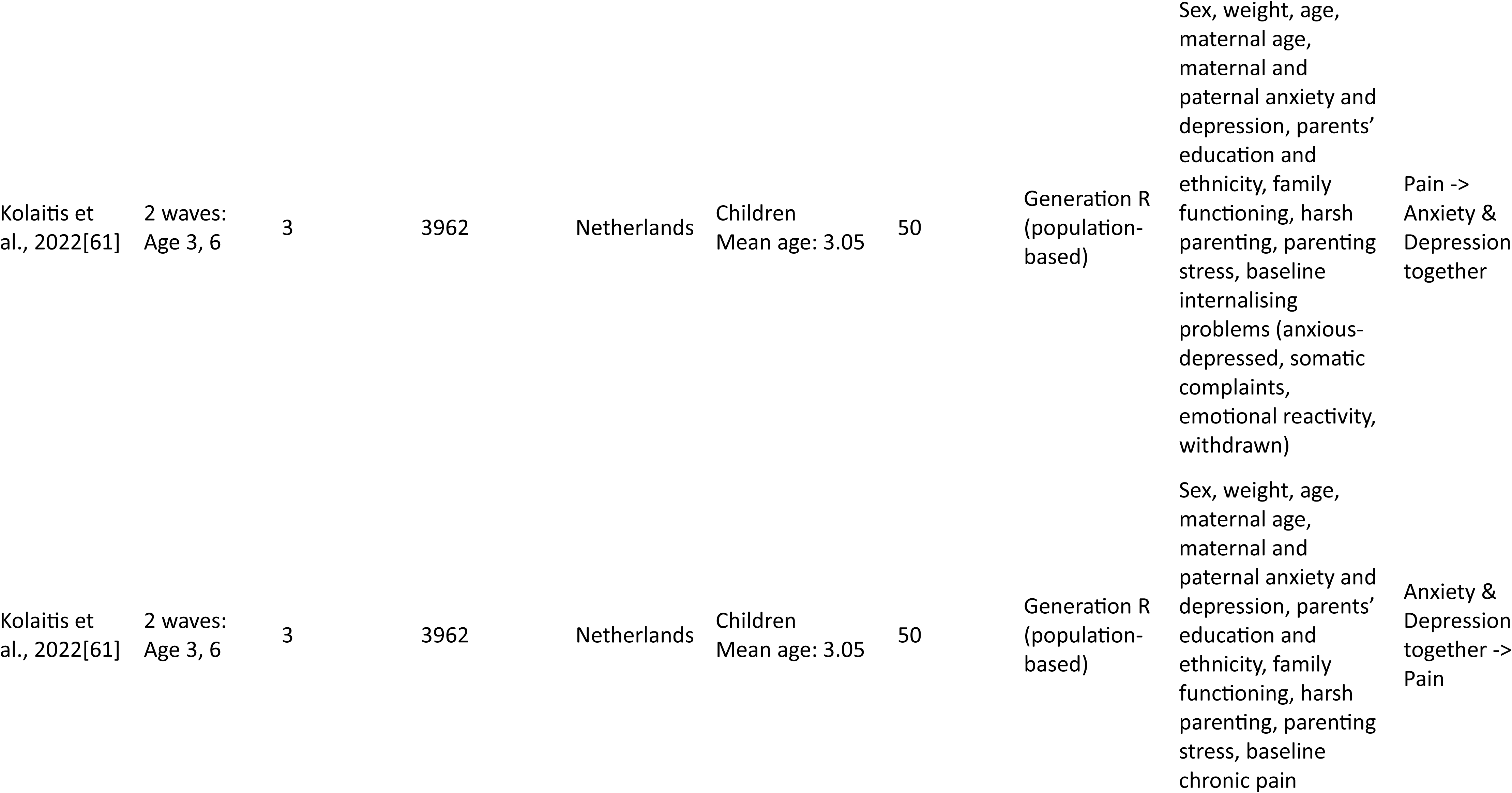
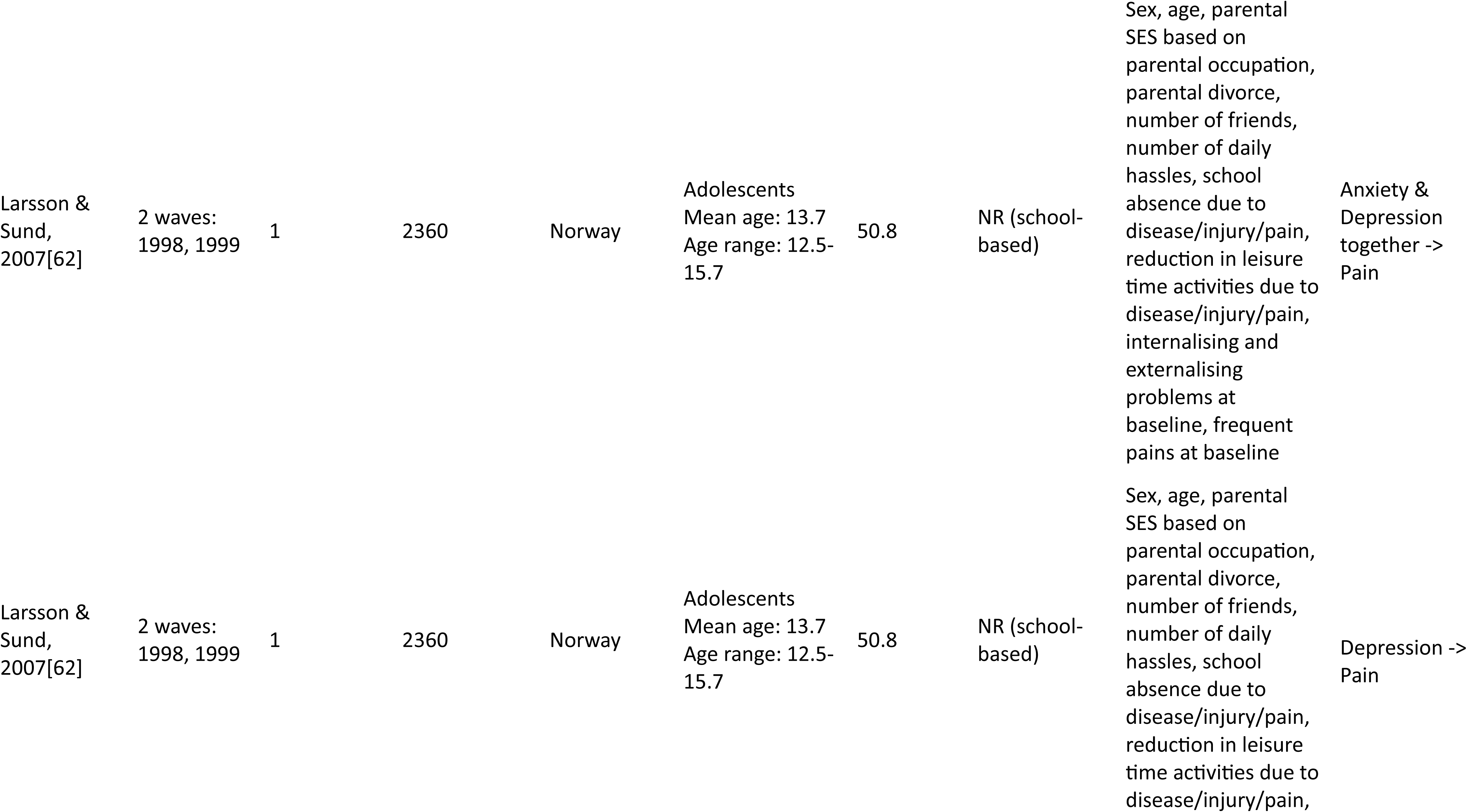
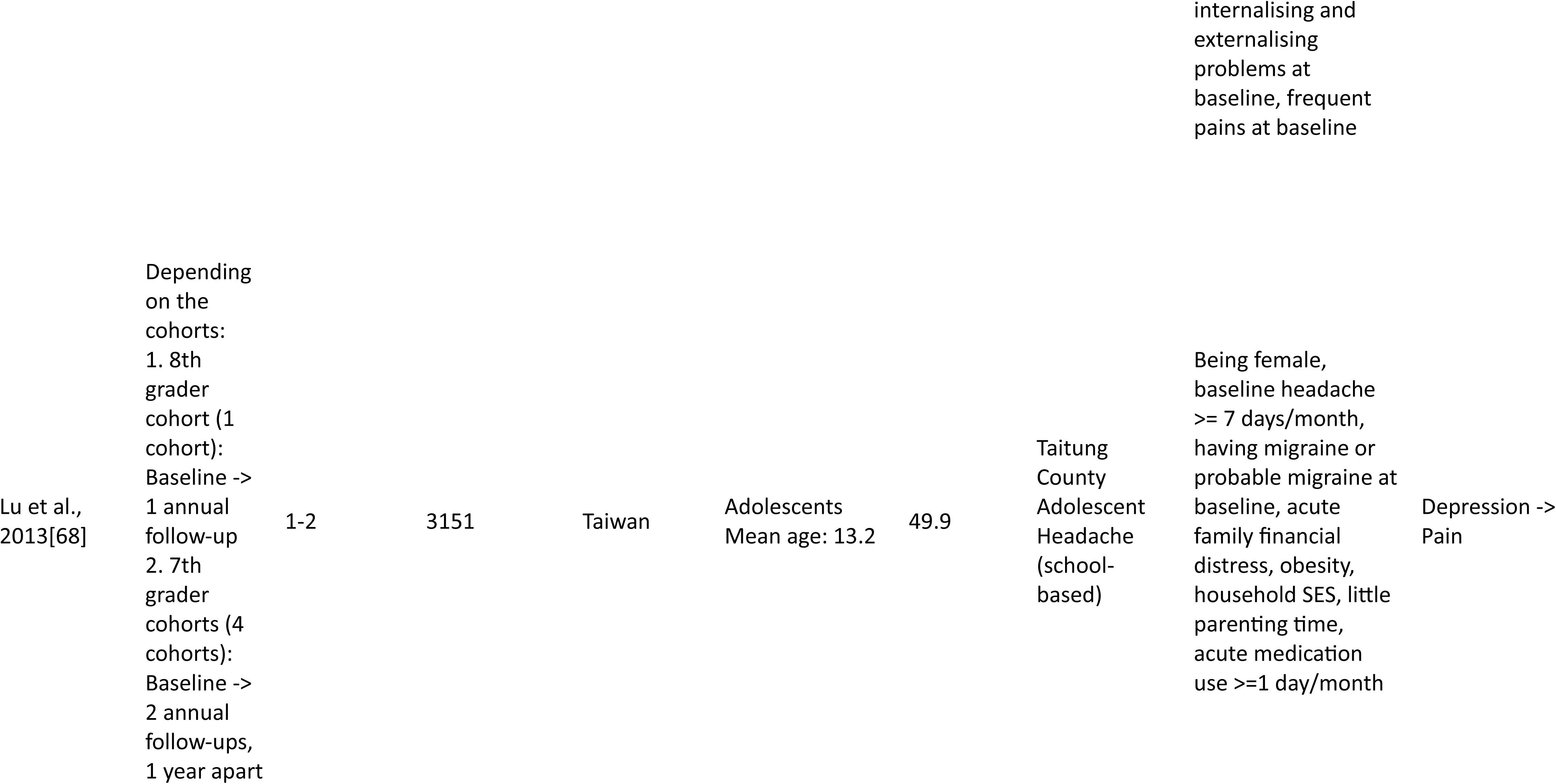
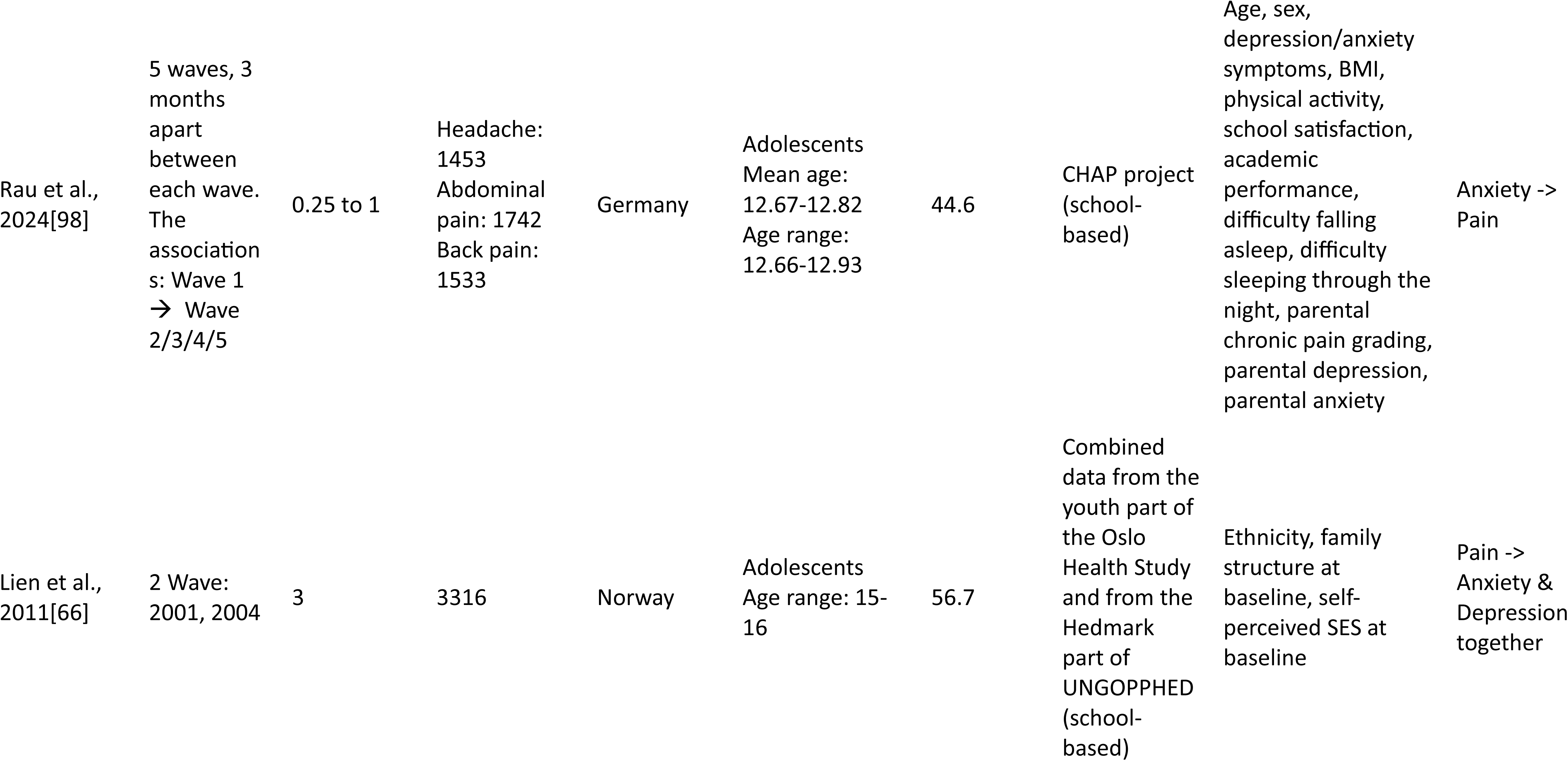
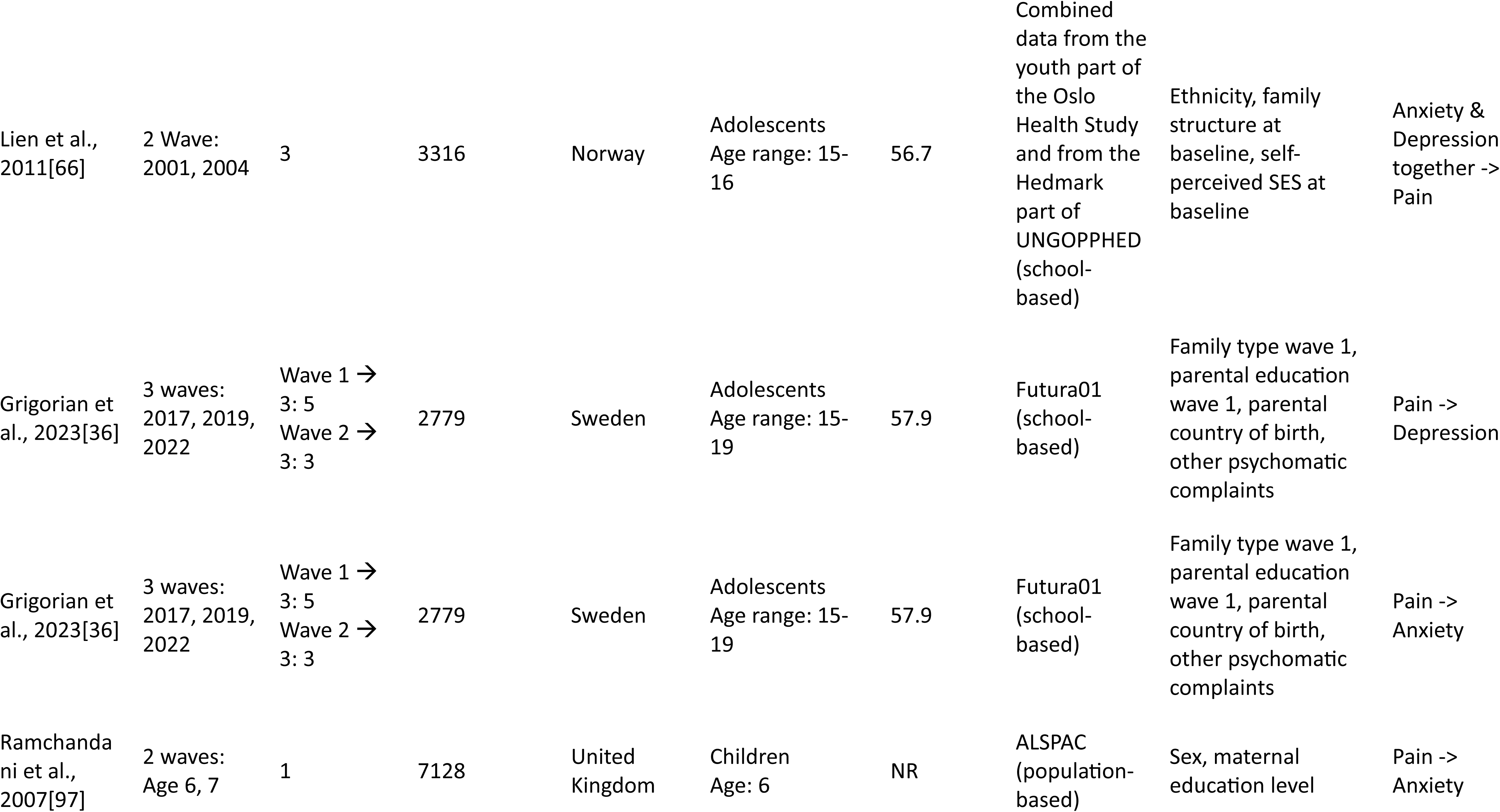
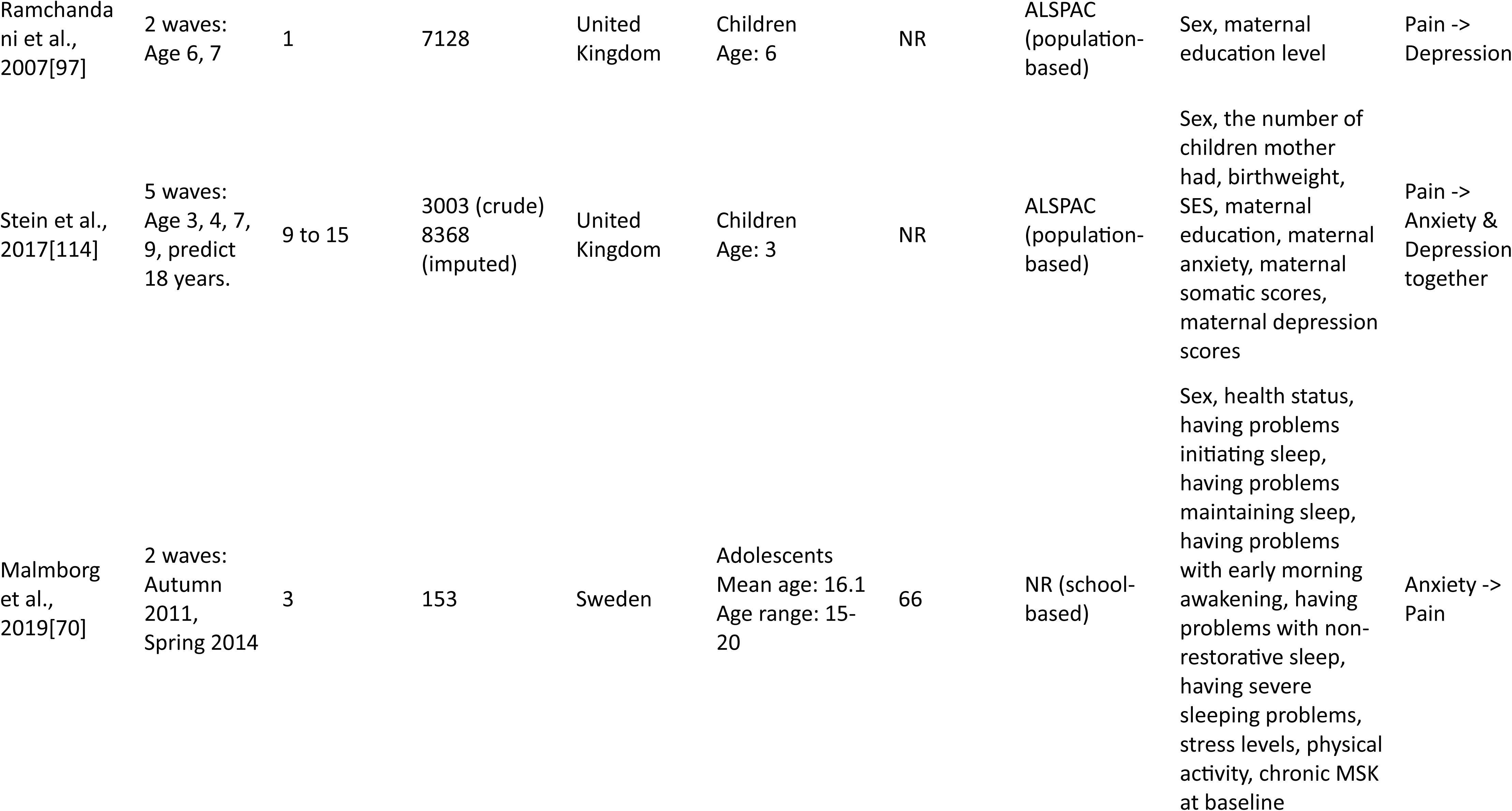
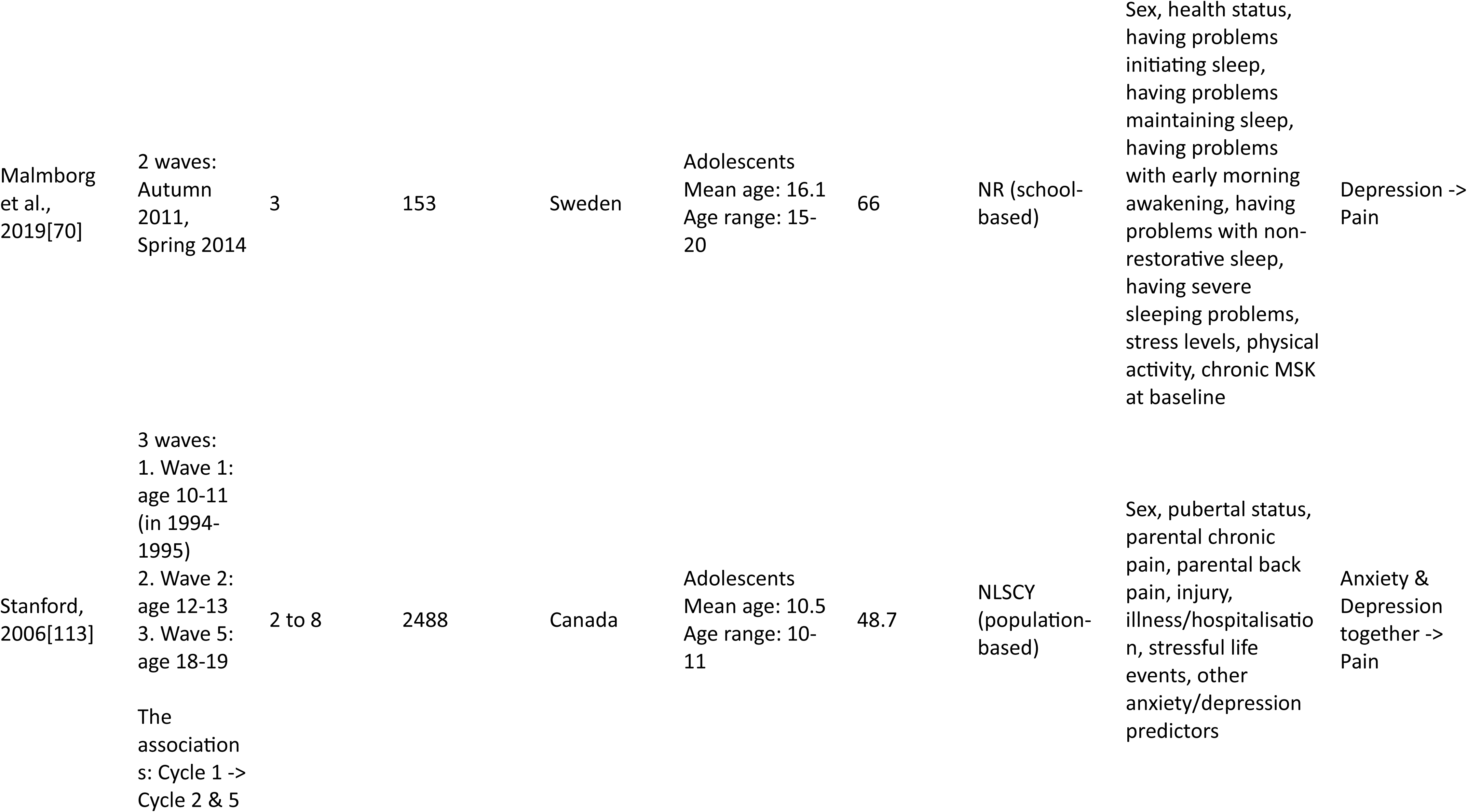
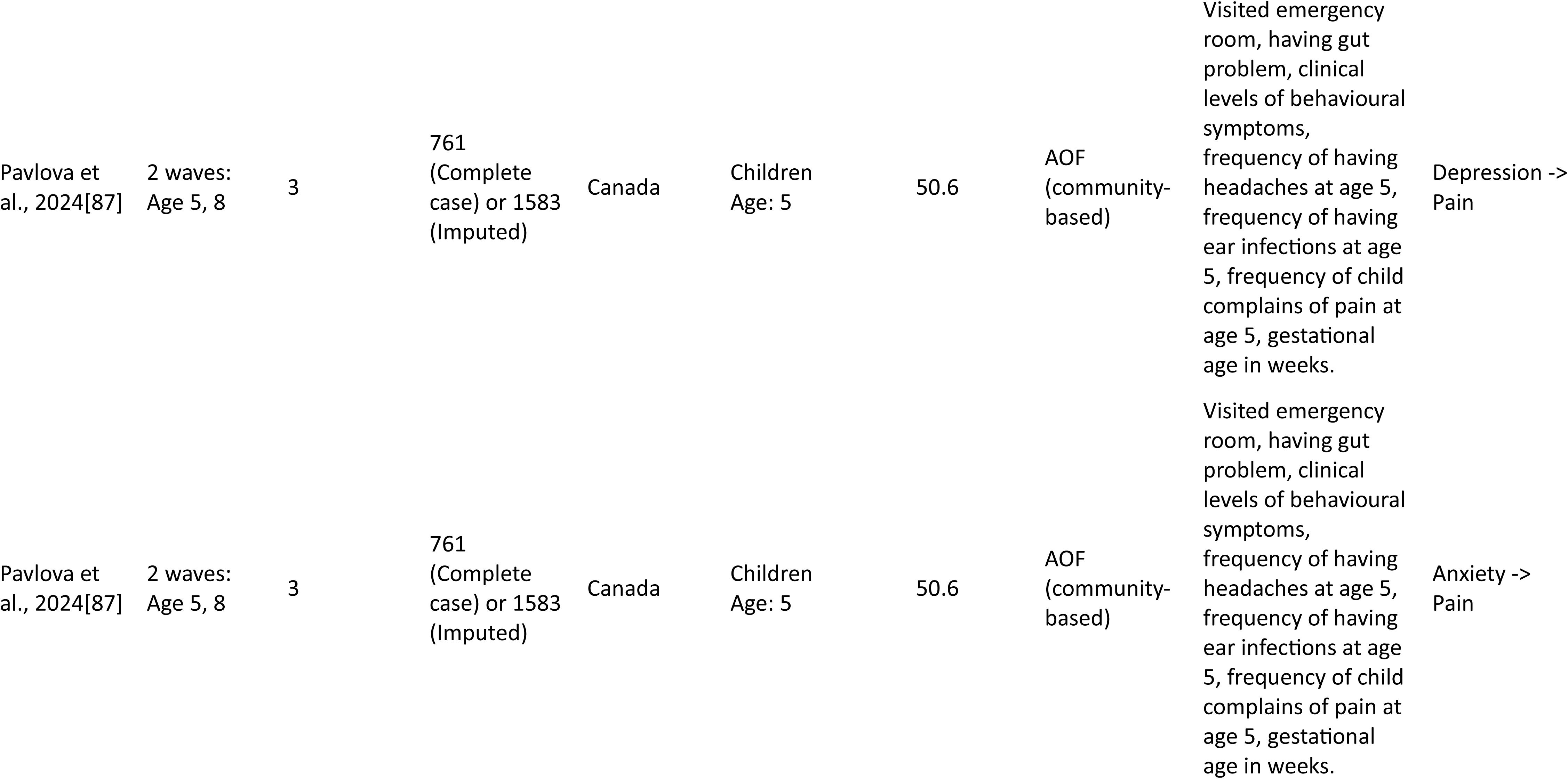
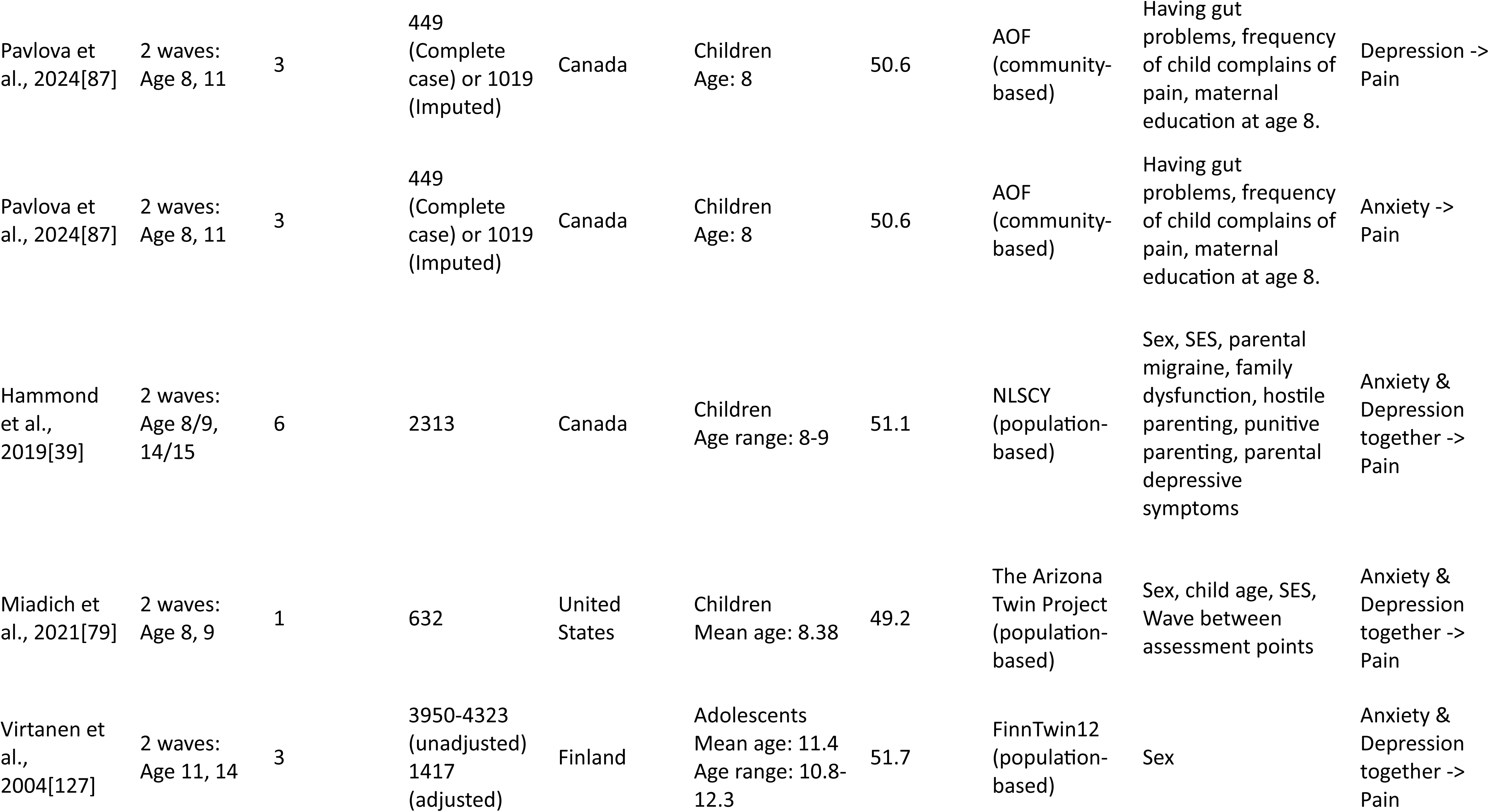
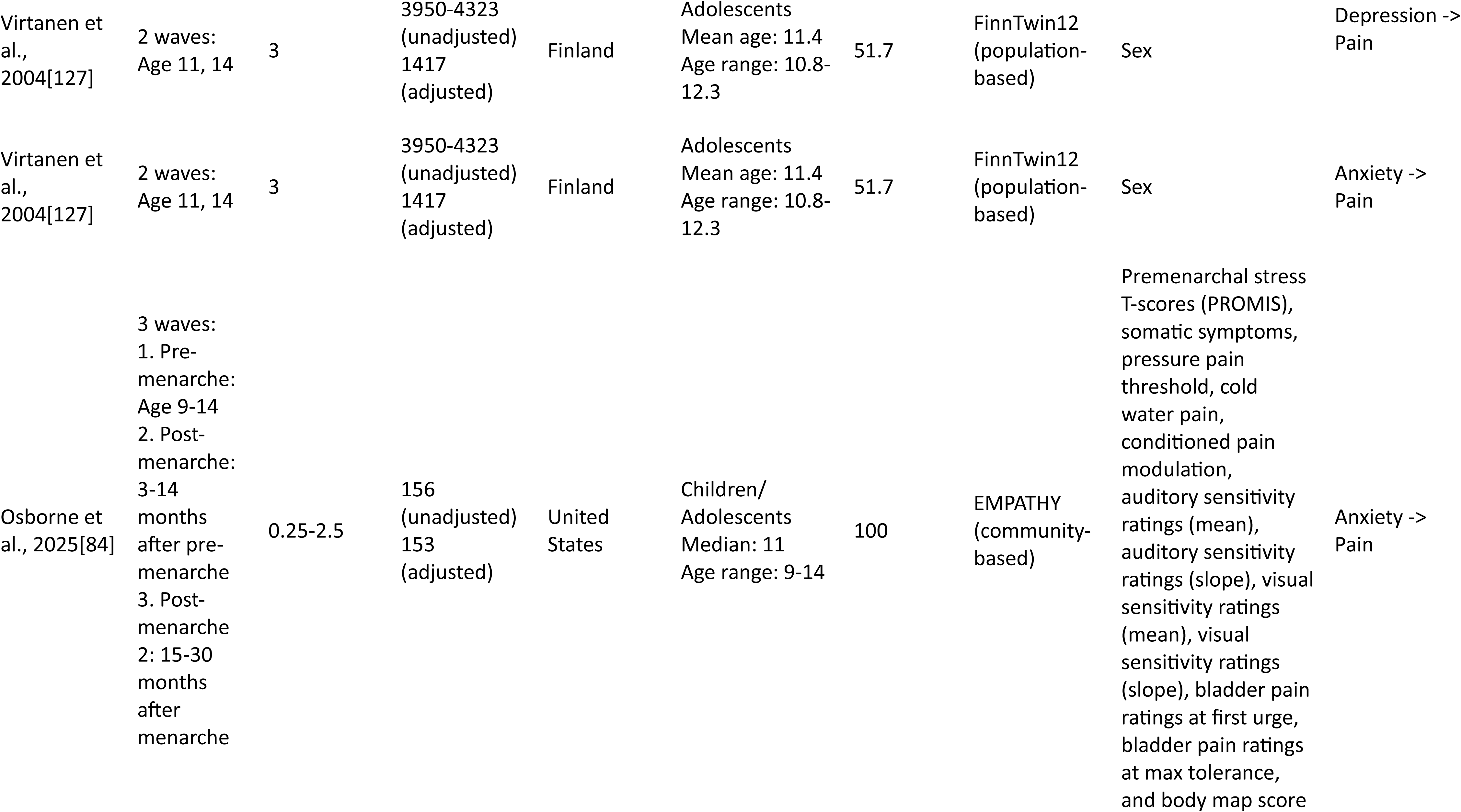
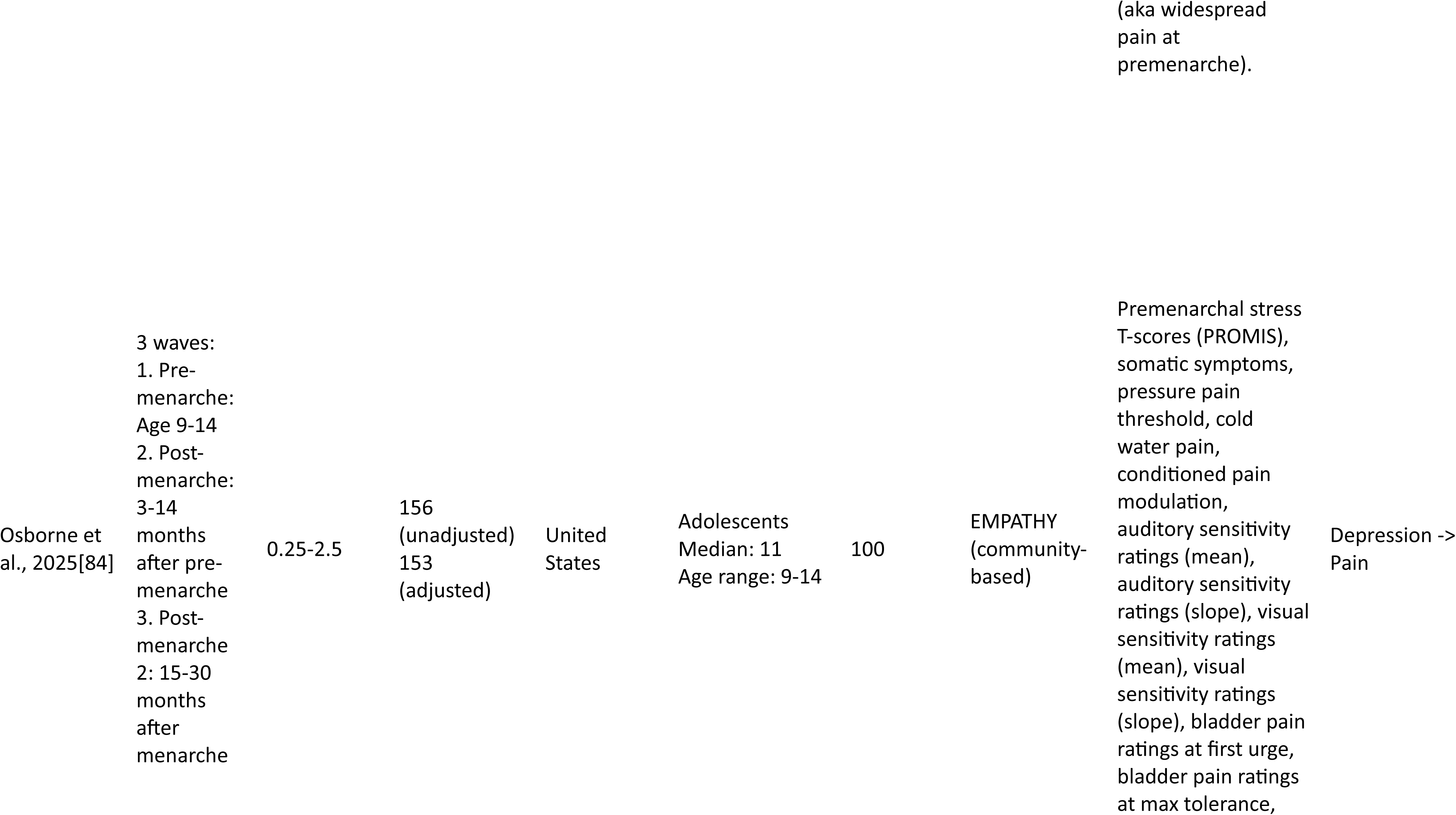
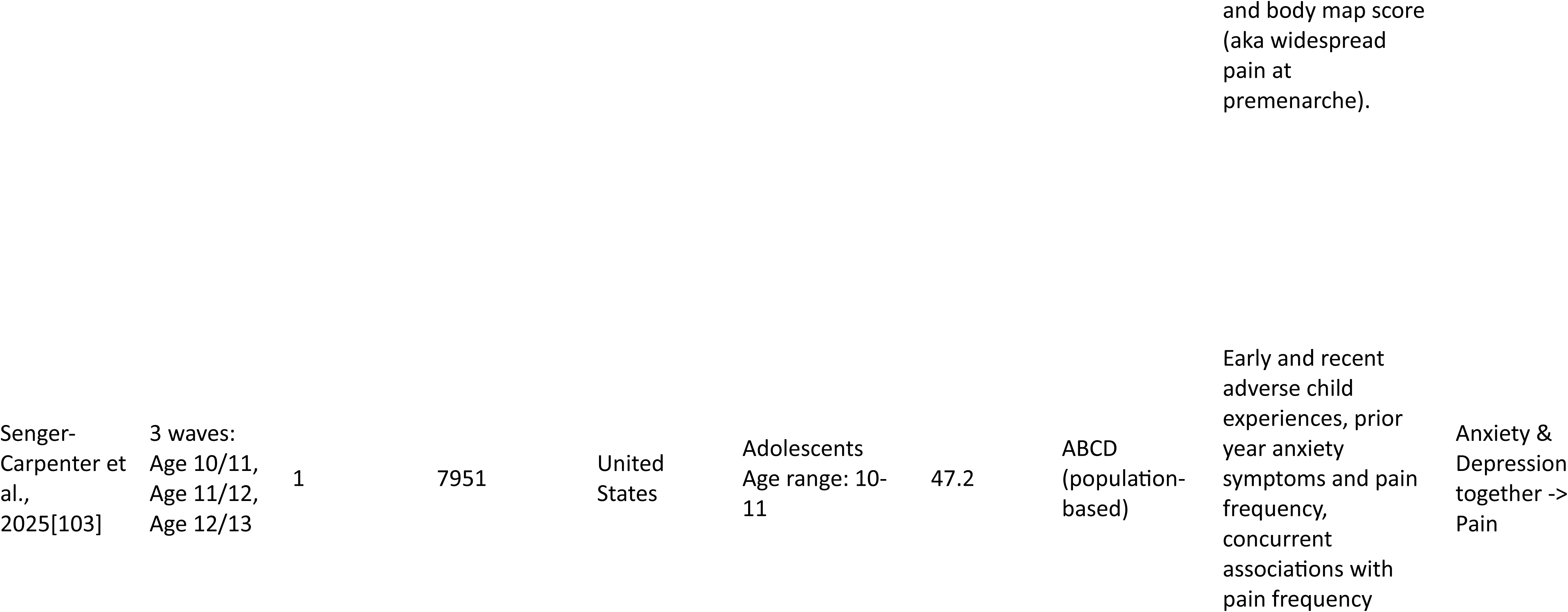
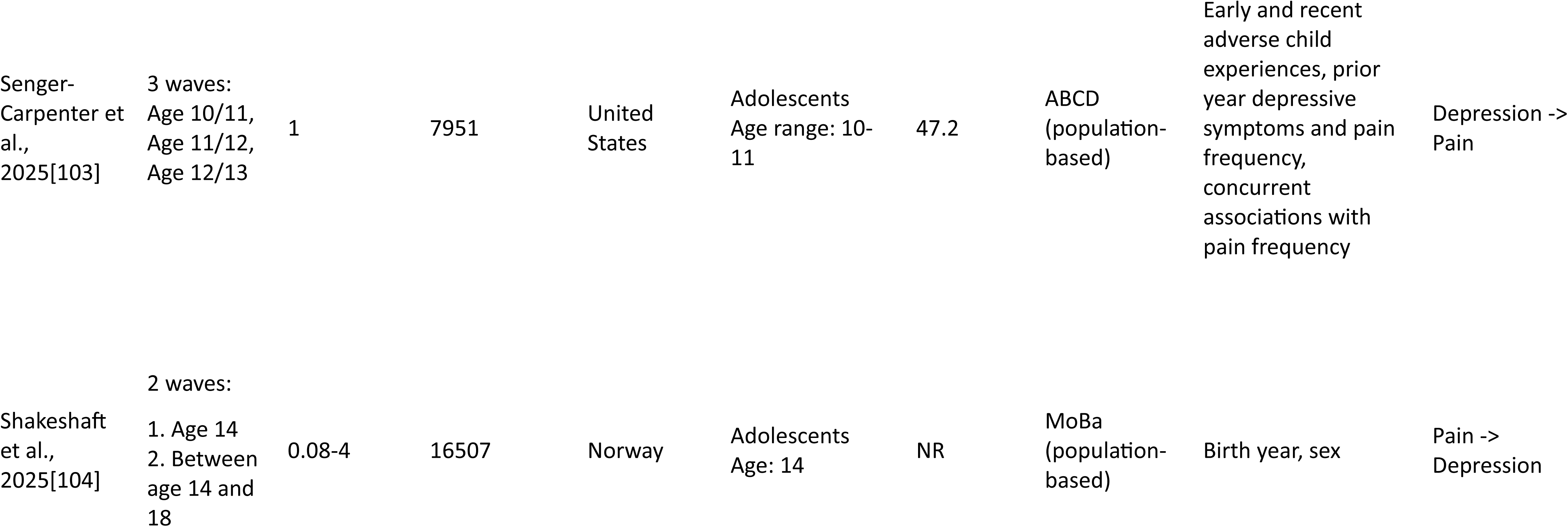
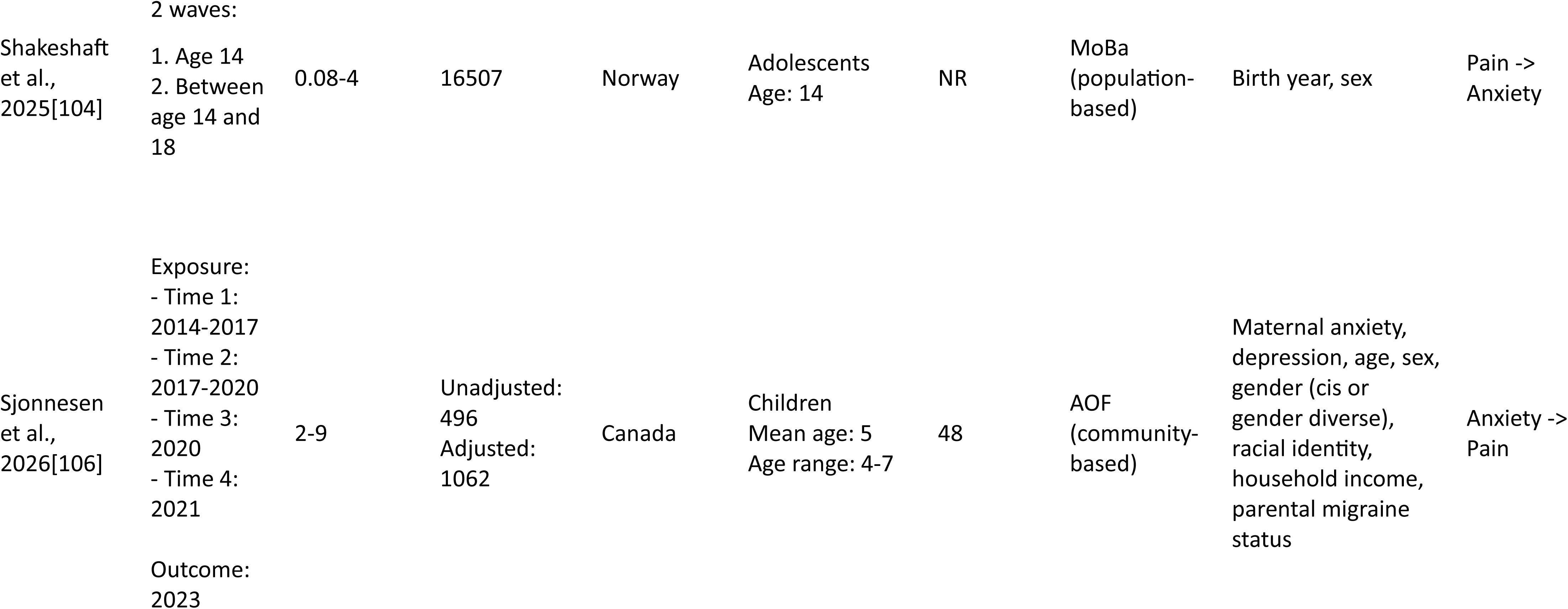
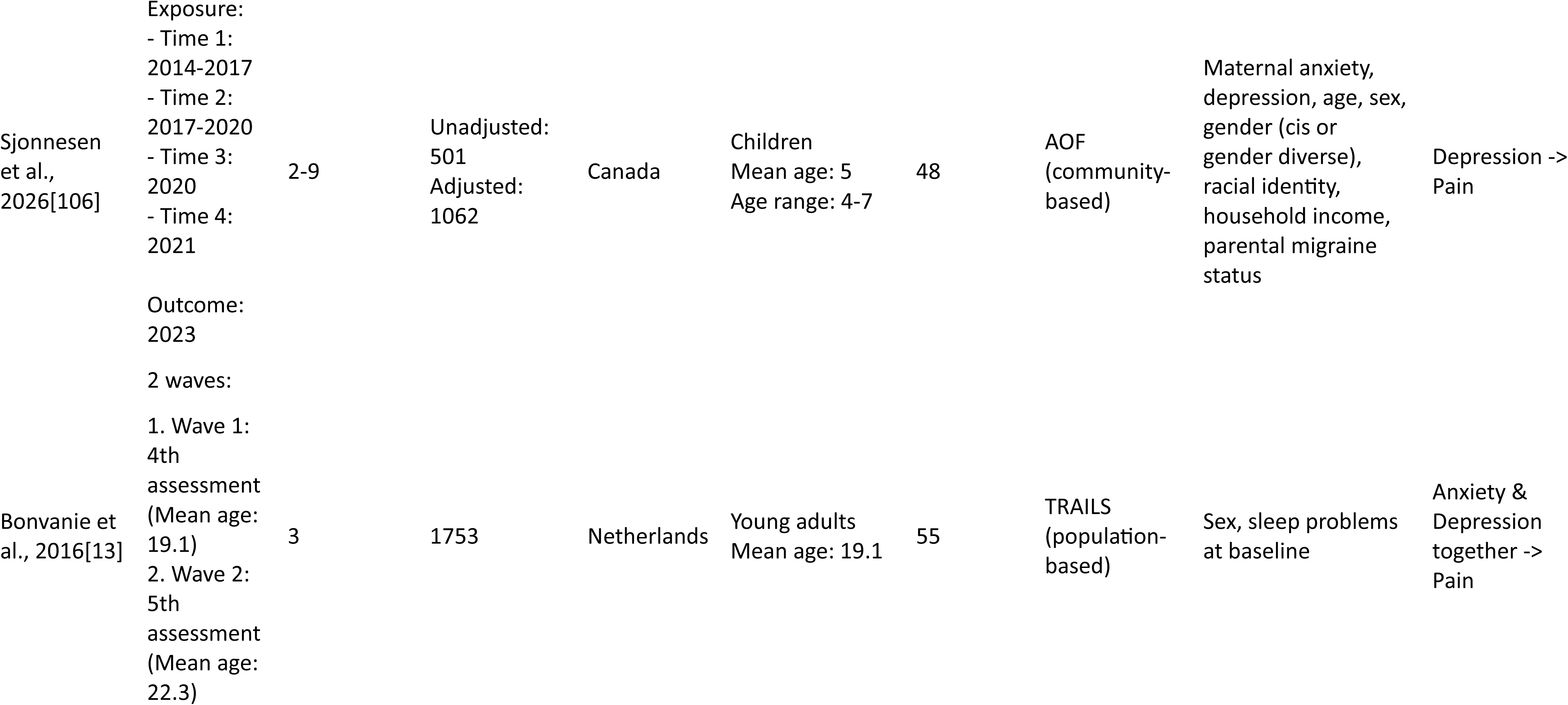
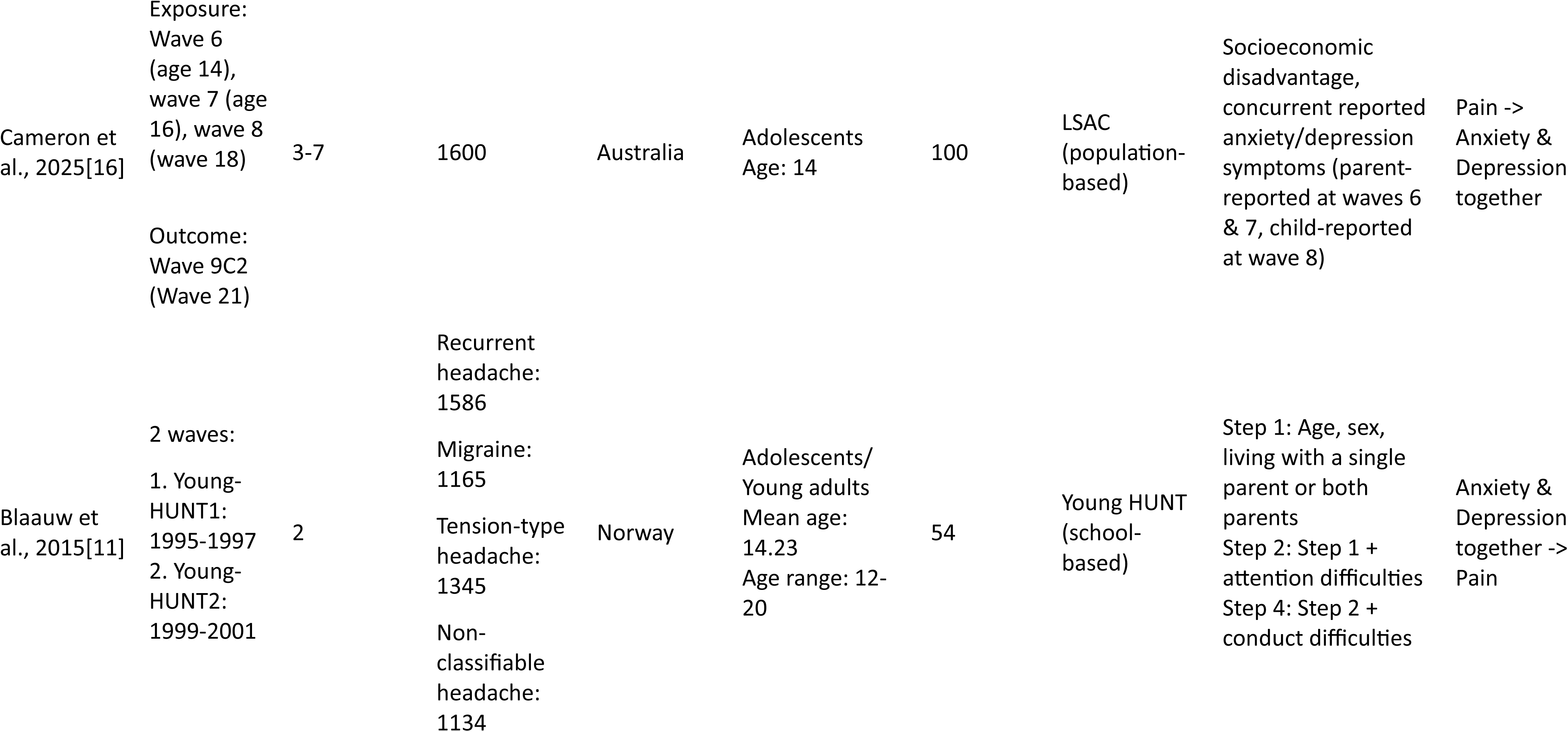
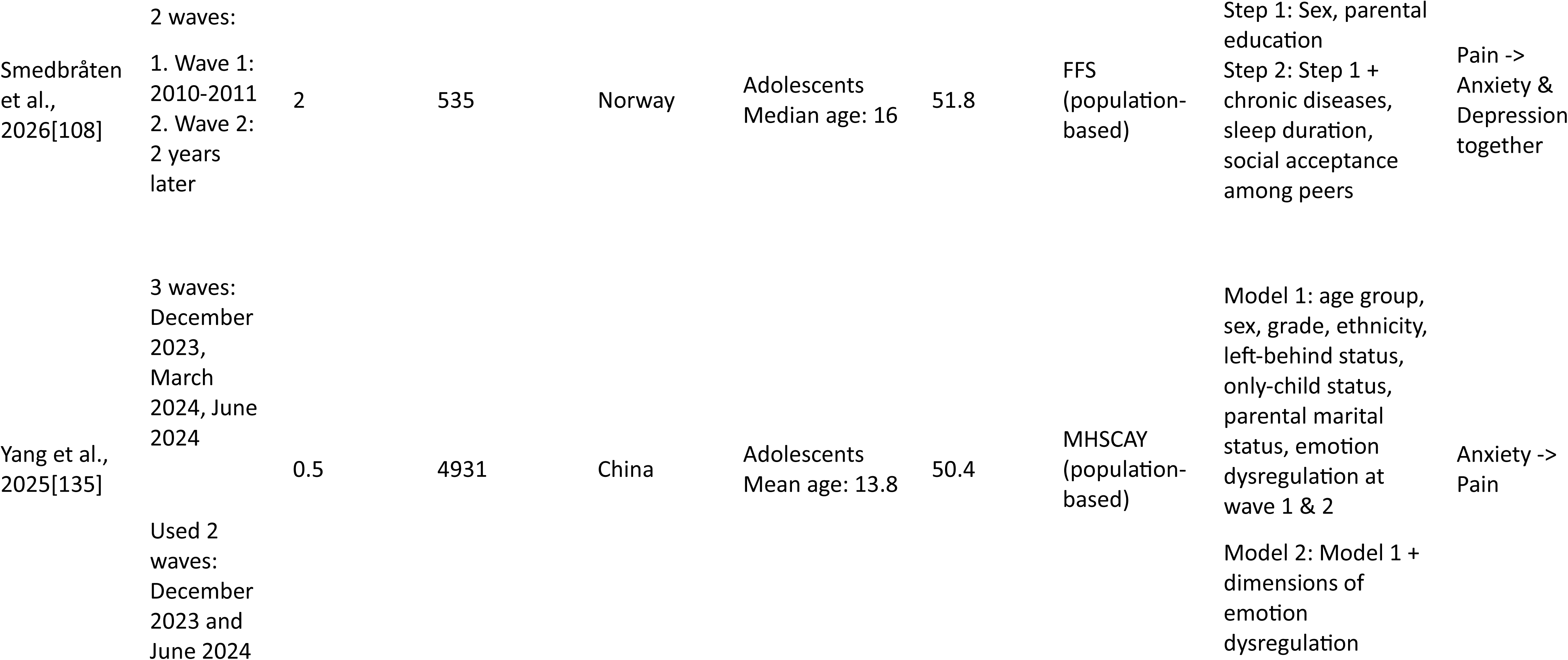
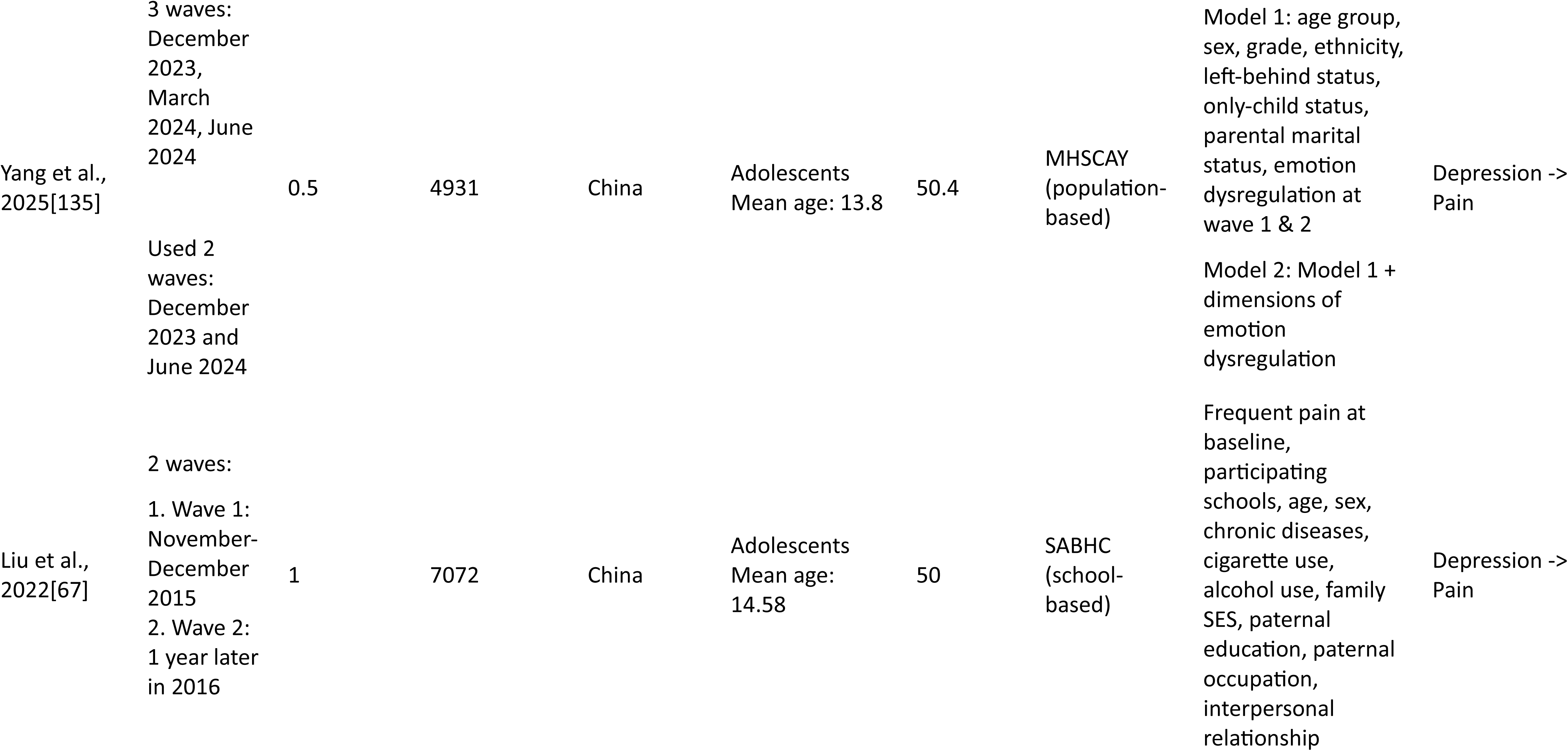
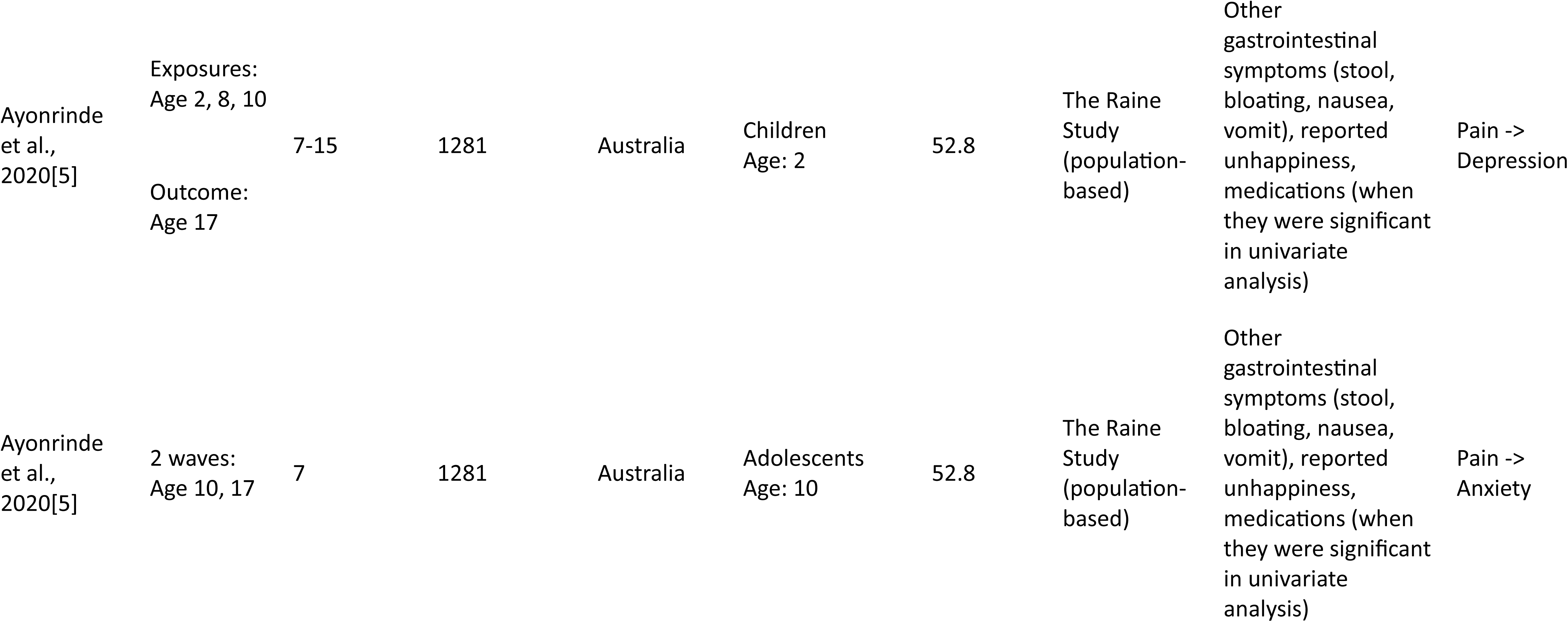
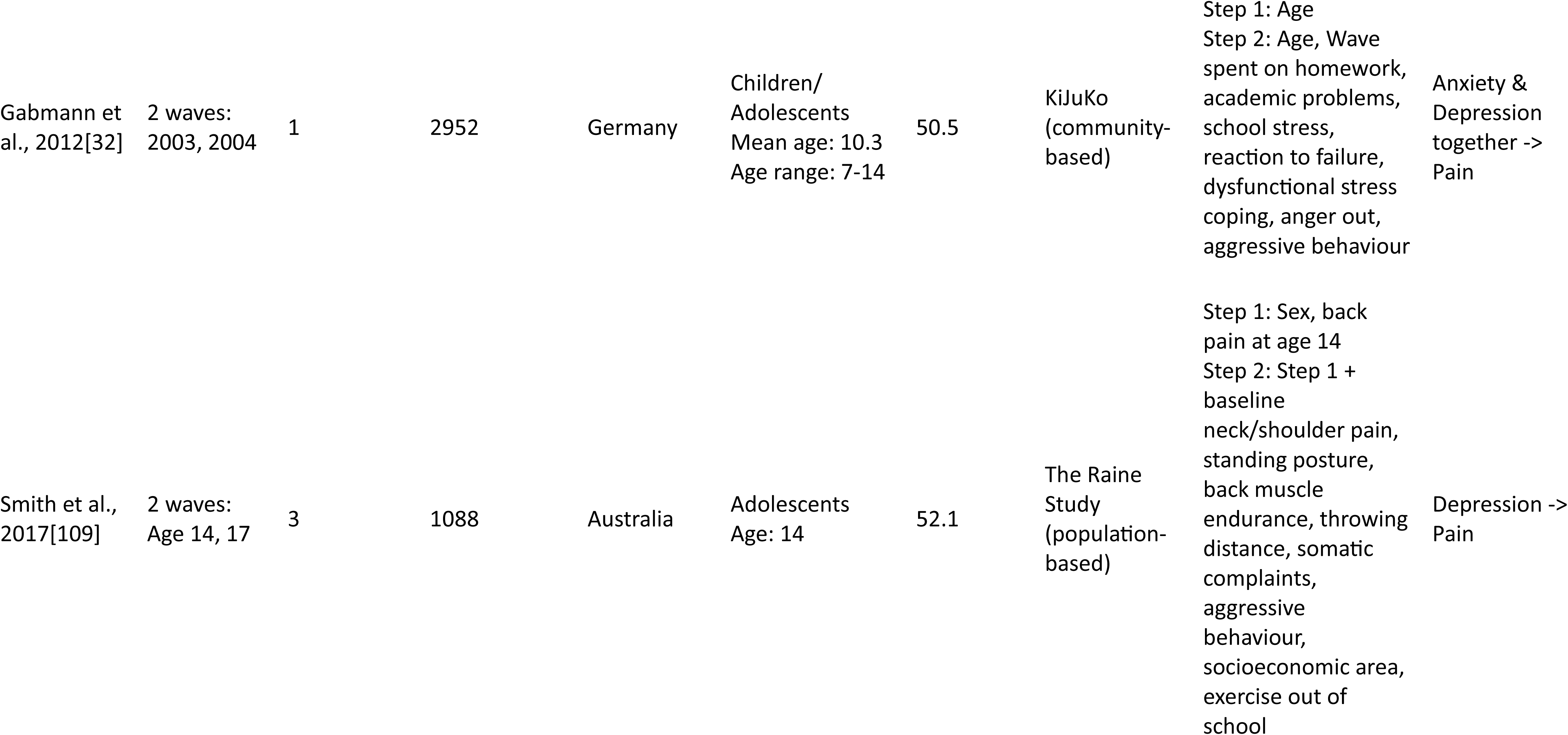
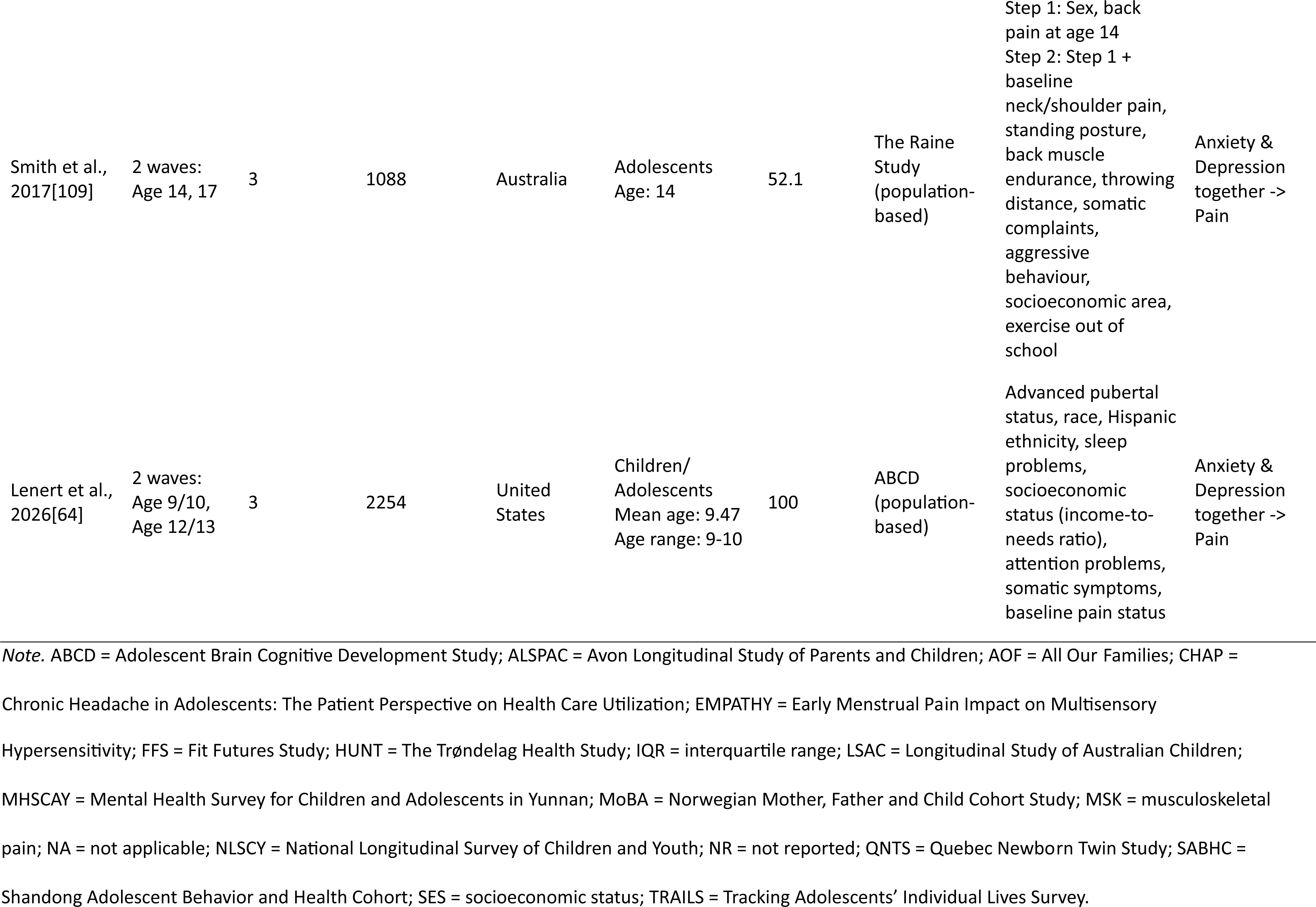
Characteristics of the included studies.

### 3.3. Measurement

Considerable variability was observed in the definition and measurement of pain, anxiety, and depression (Table 2). Pain was conceptualised as continuous symptoms (*k* = 15), binary presence within a specified period (*k* = 26), or ordinal indicators of pain (*k* = 2). One study used both binary and ordinal conceptualisations. There was little overlap in pain measures: three cohorts used the WHO Health Behaviour Survey of School-Aged Children; three used the Numeric Rating Scale; and all remaining cohorts used different instruments. Pain was generally self-reported by adolescents and young adults. It was mainly reported by parents and/or teachers for children.

**Table 2.**
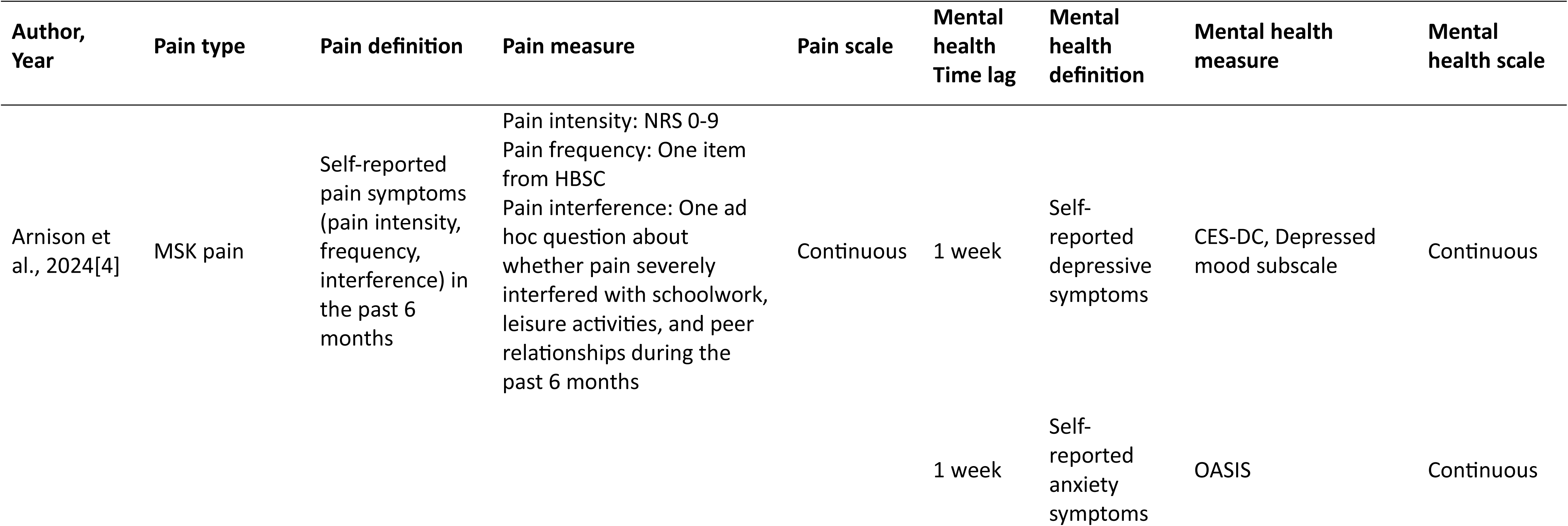
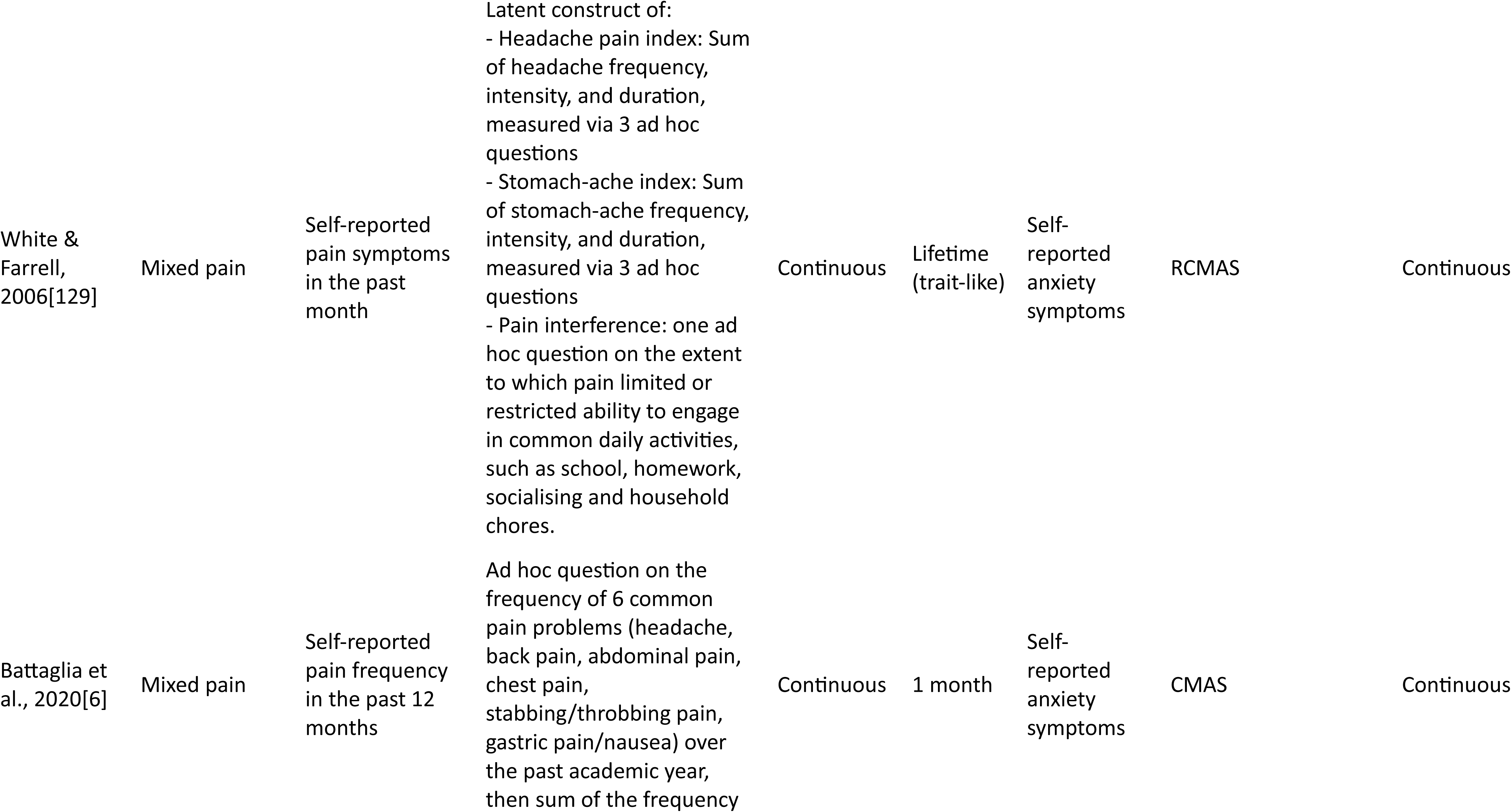
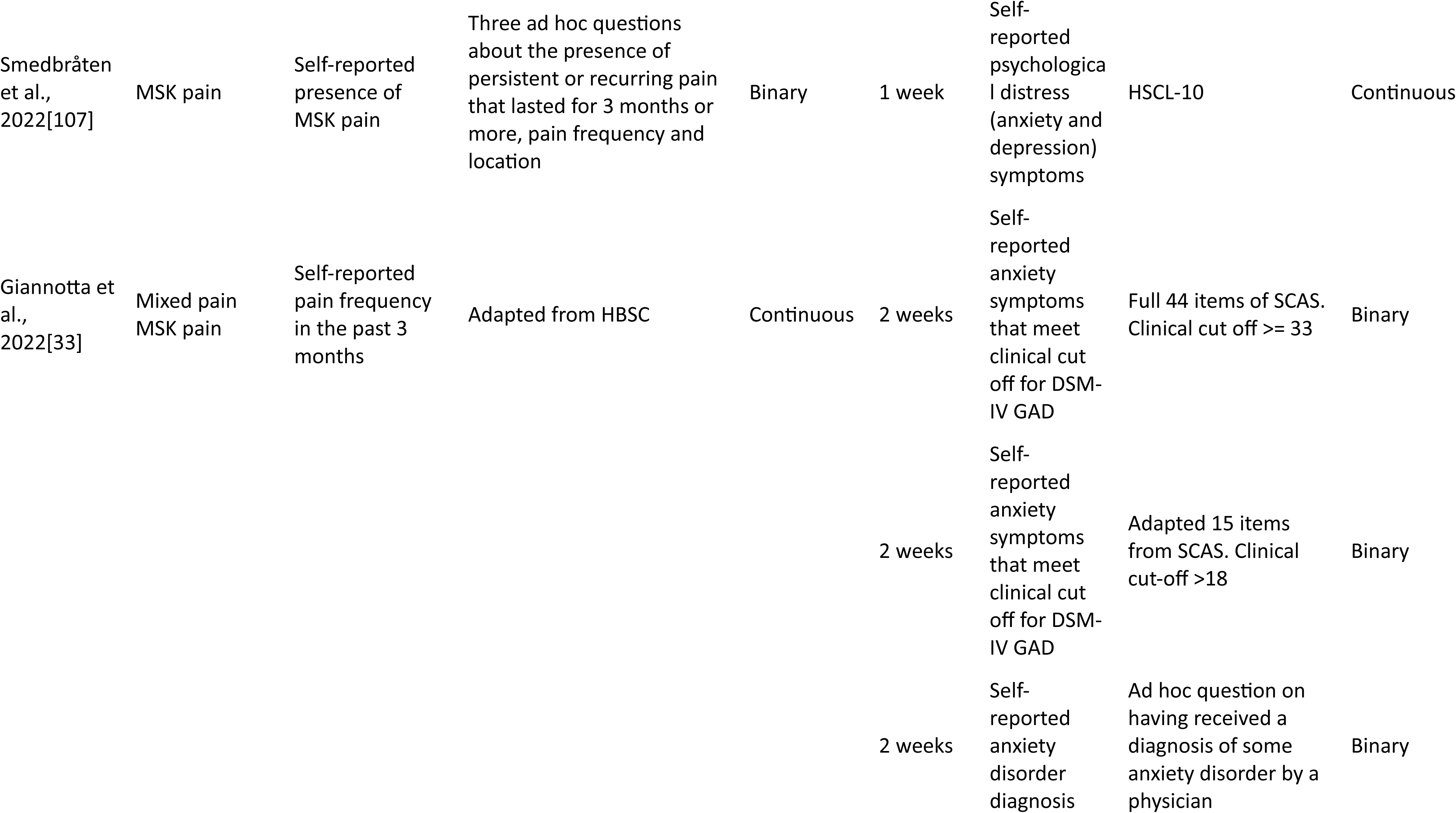
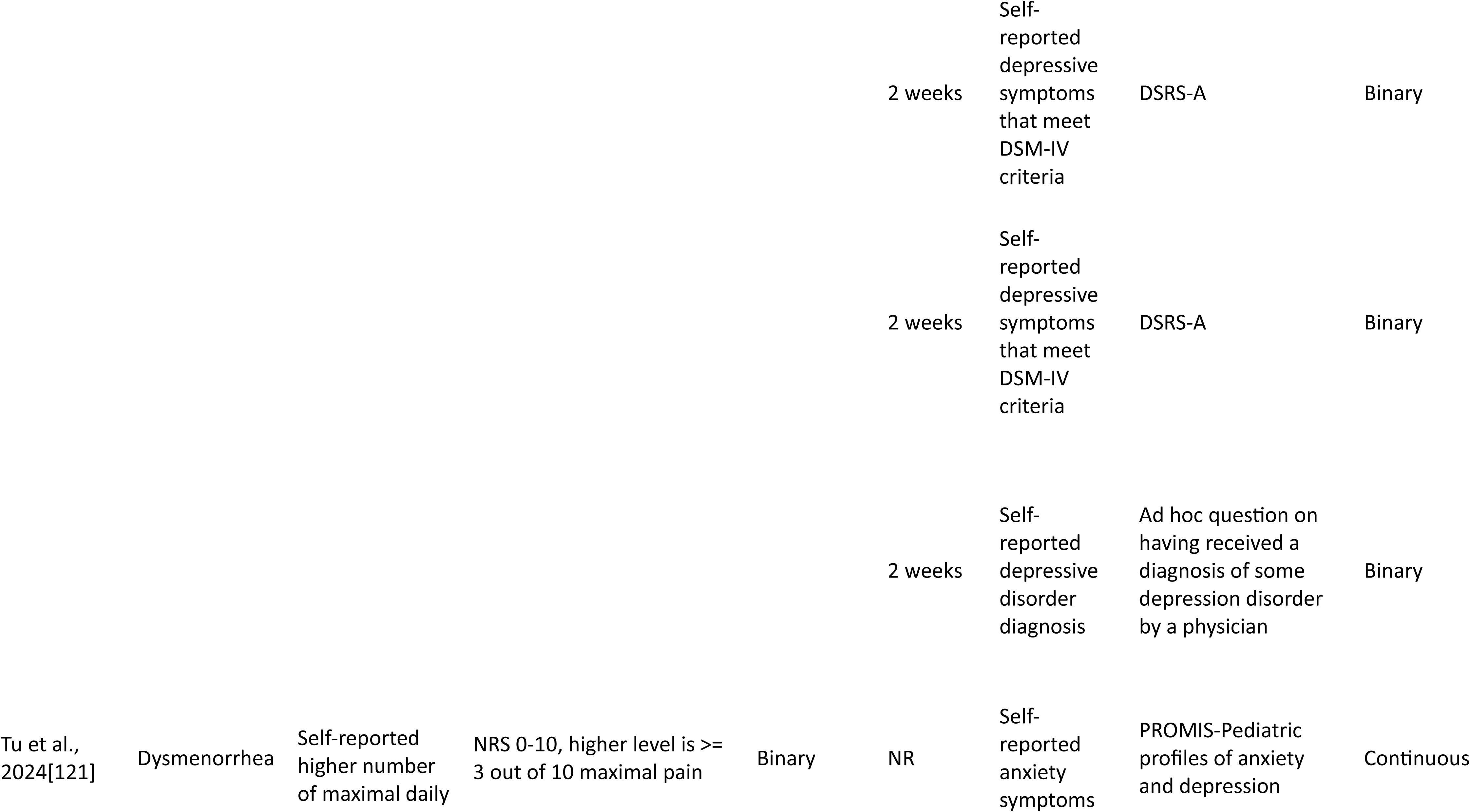
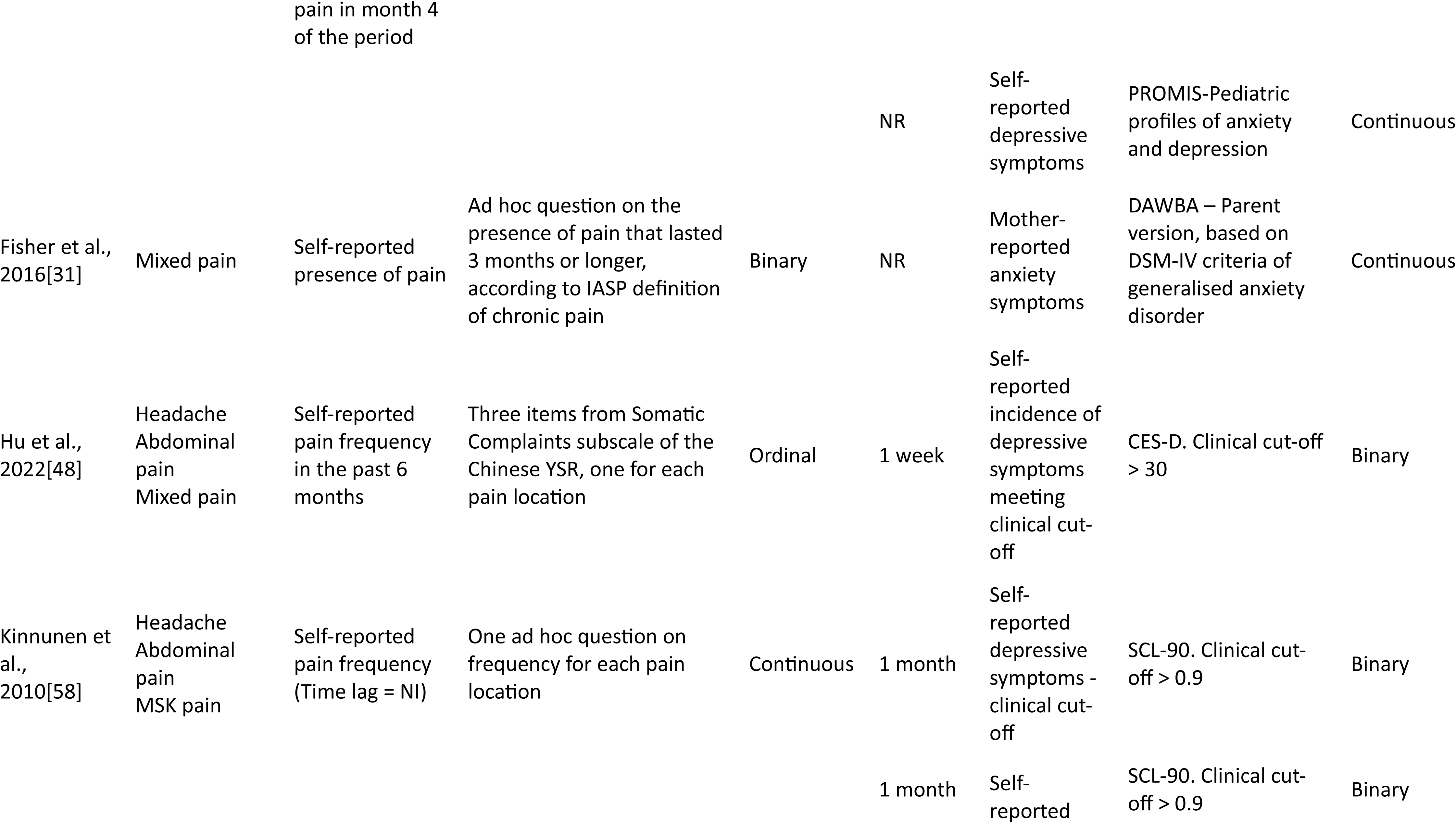
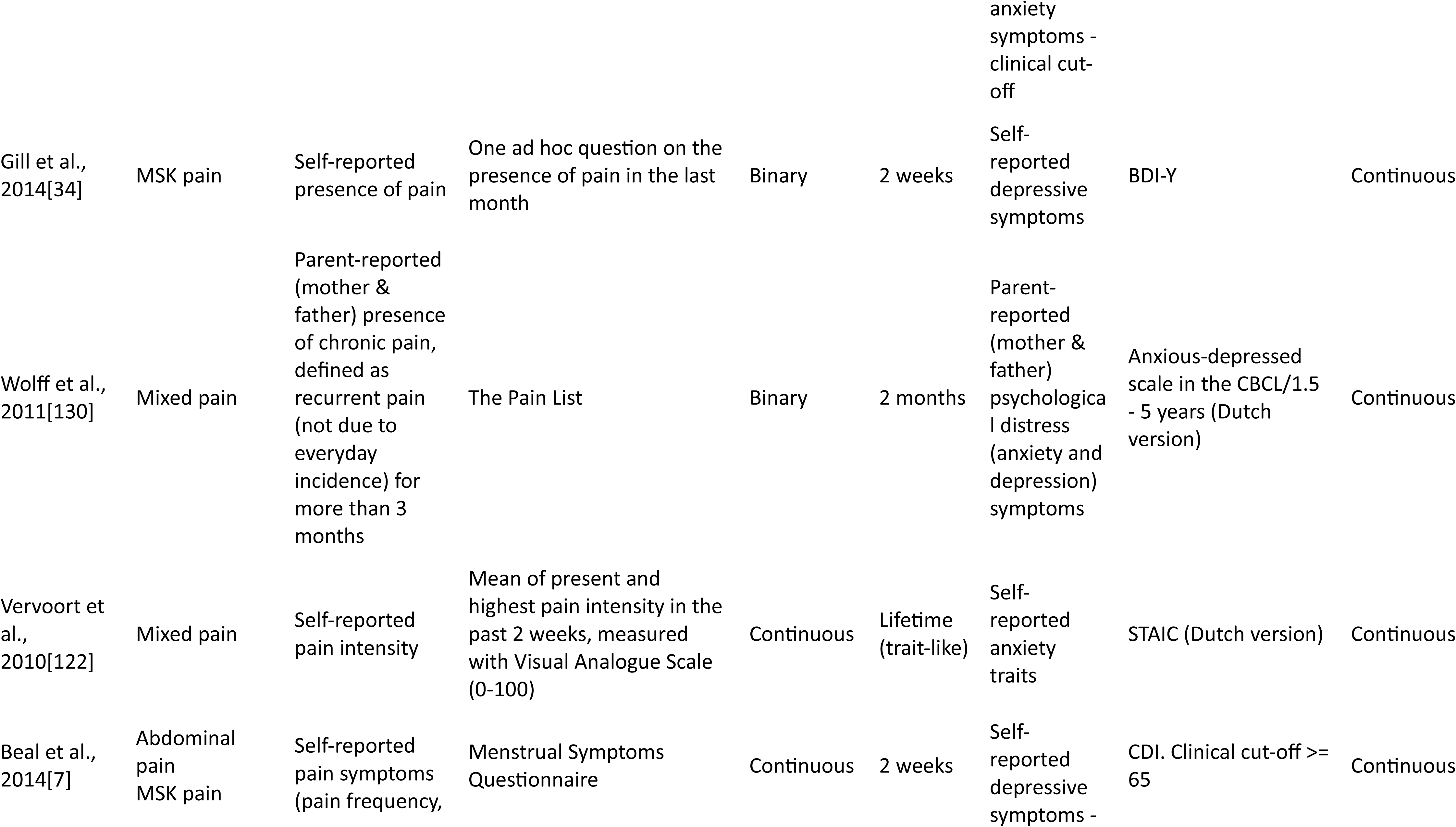
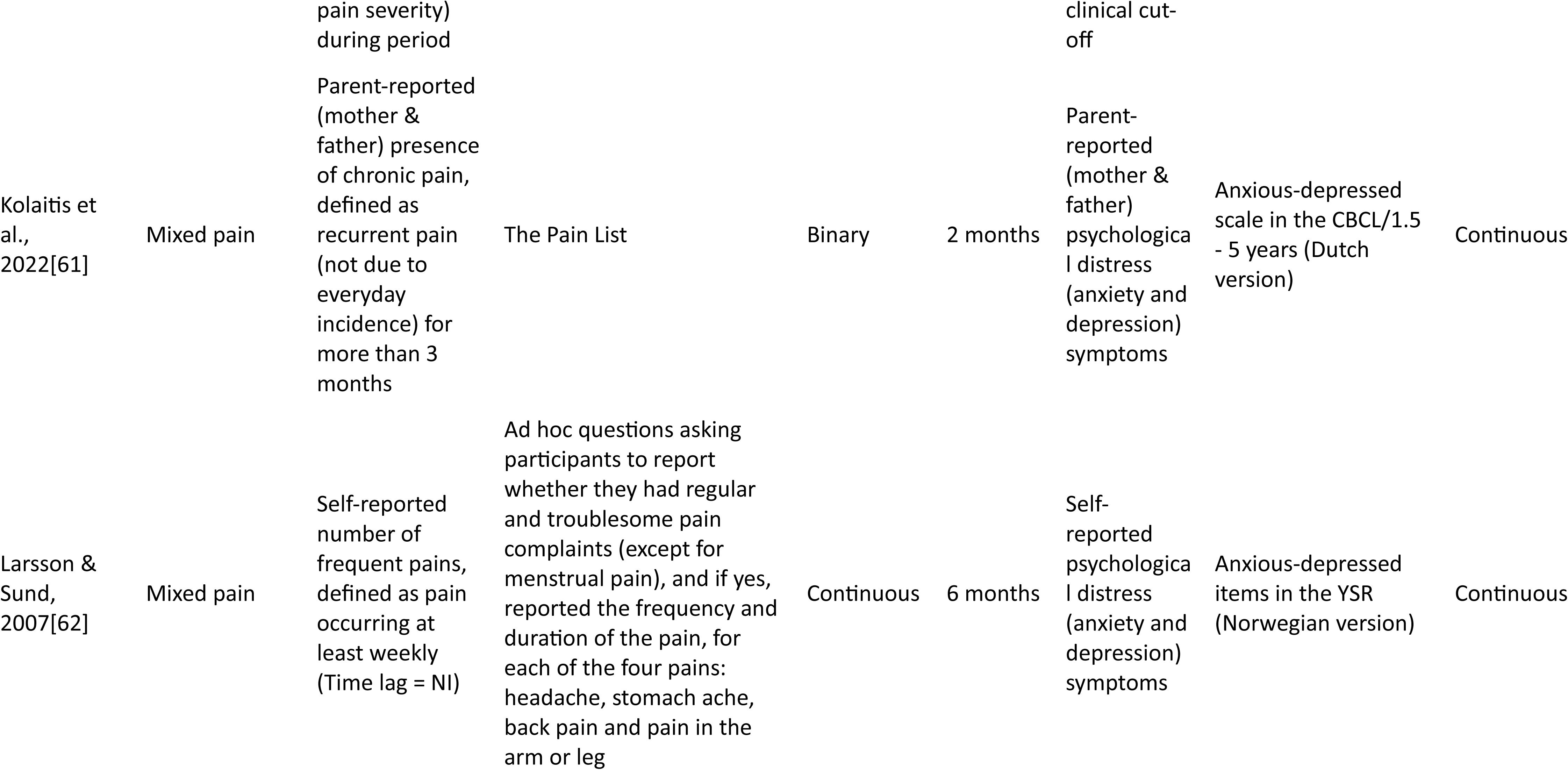
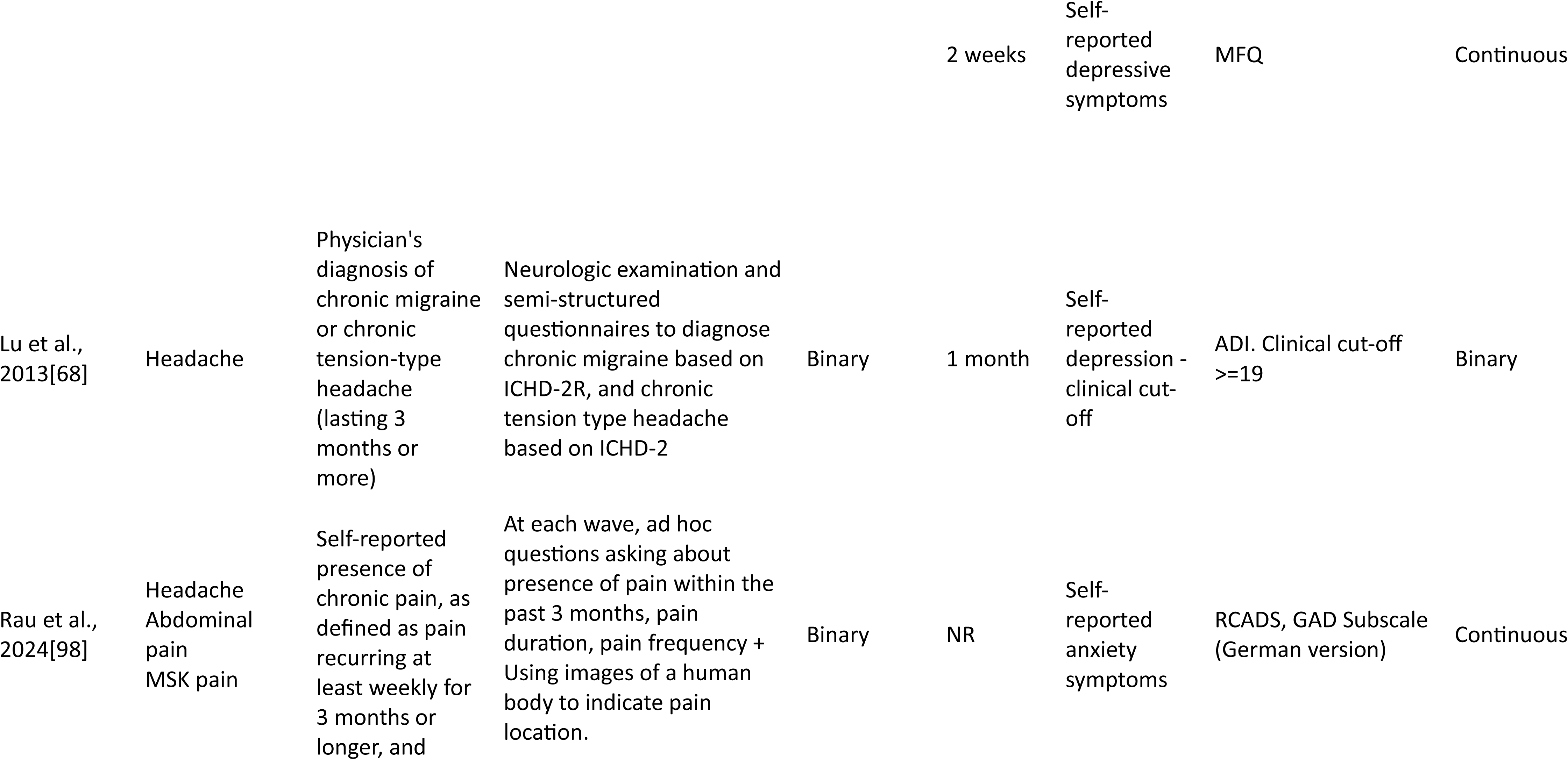
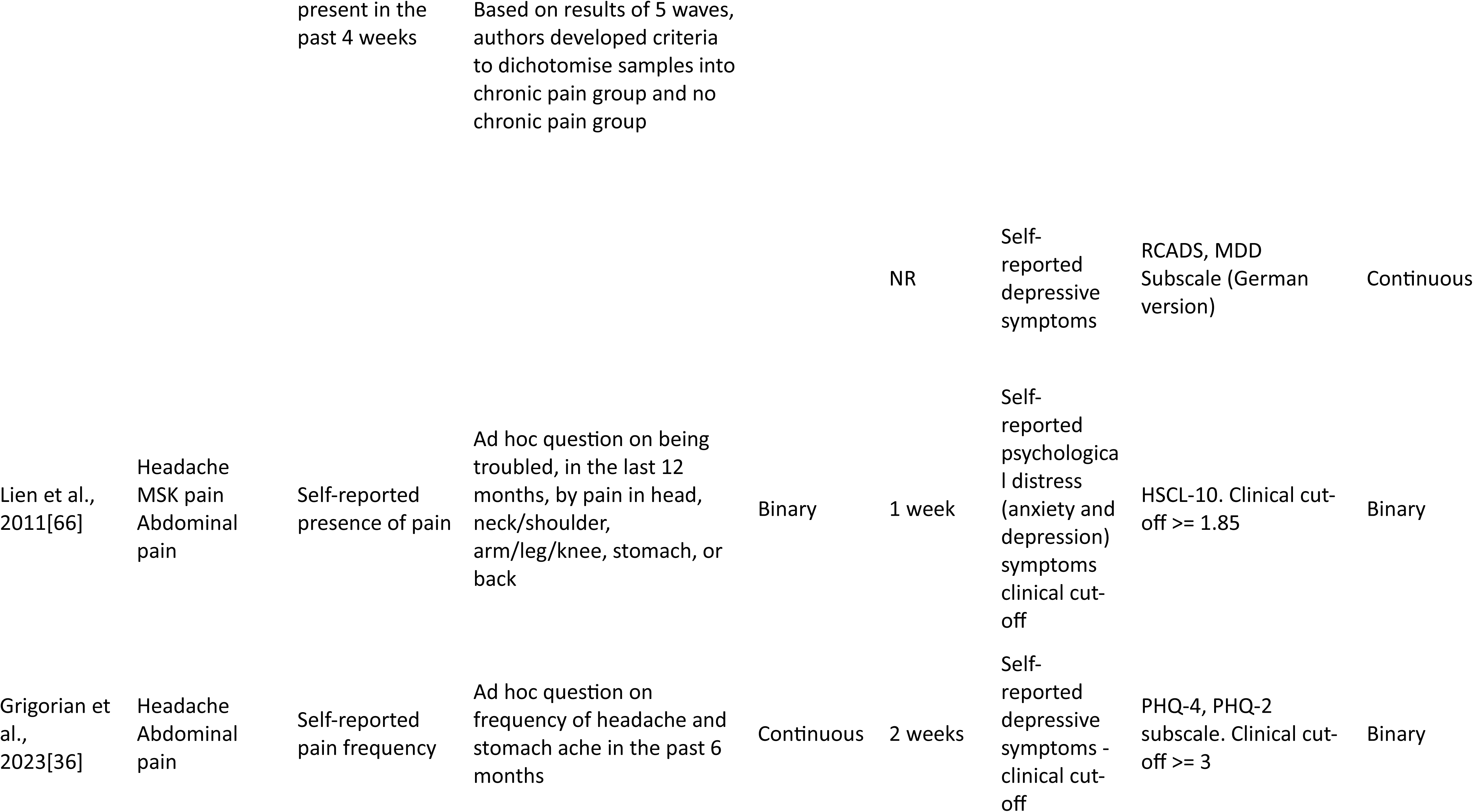
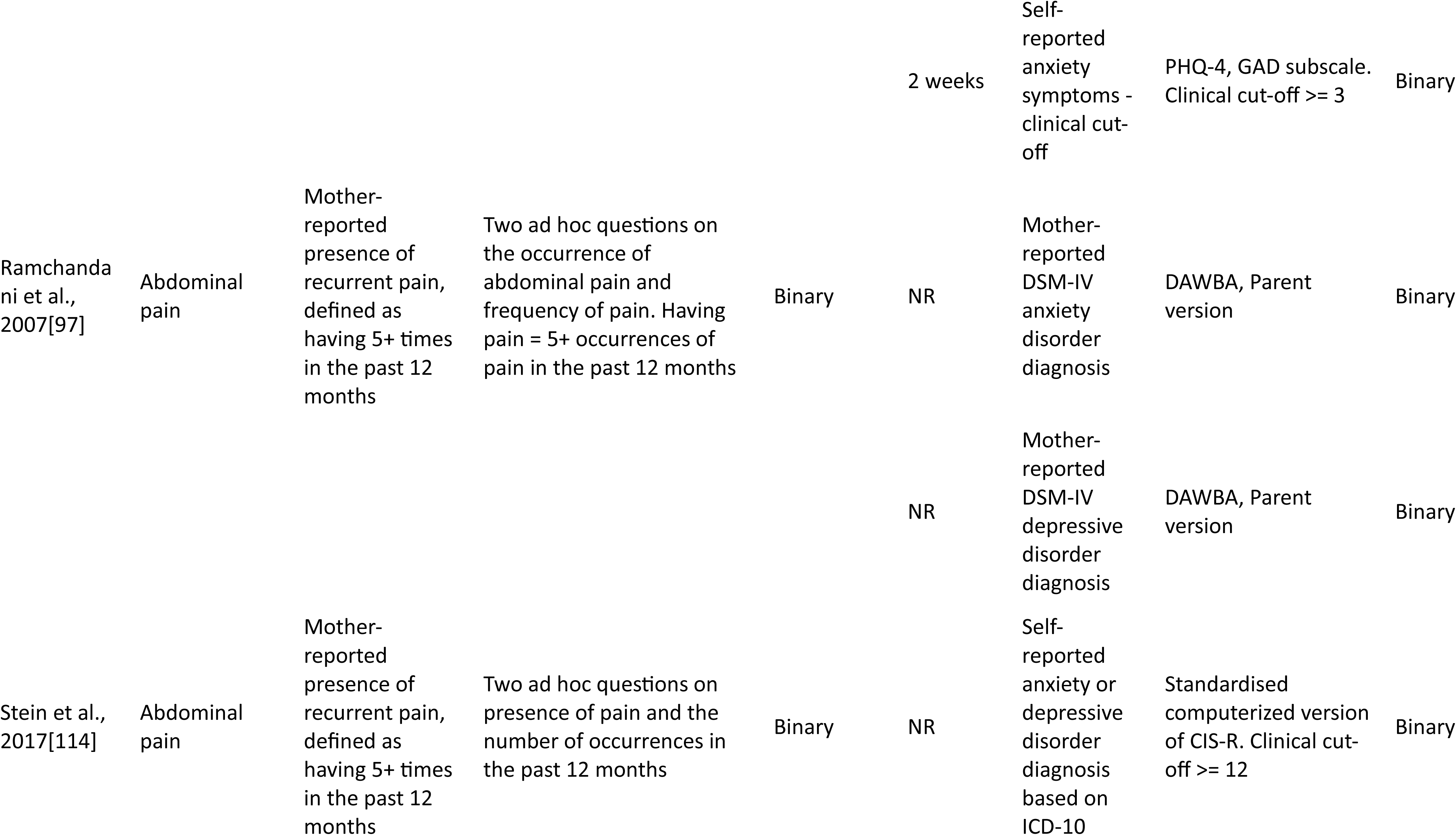
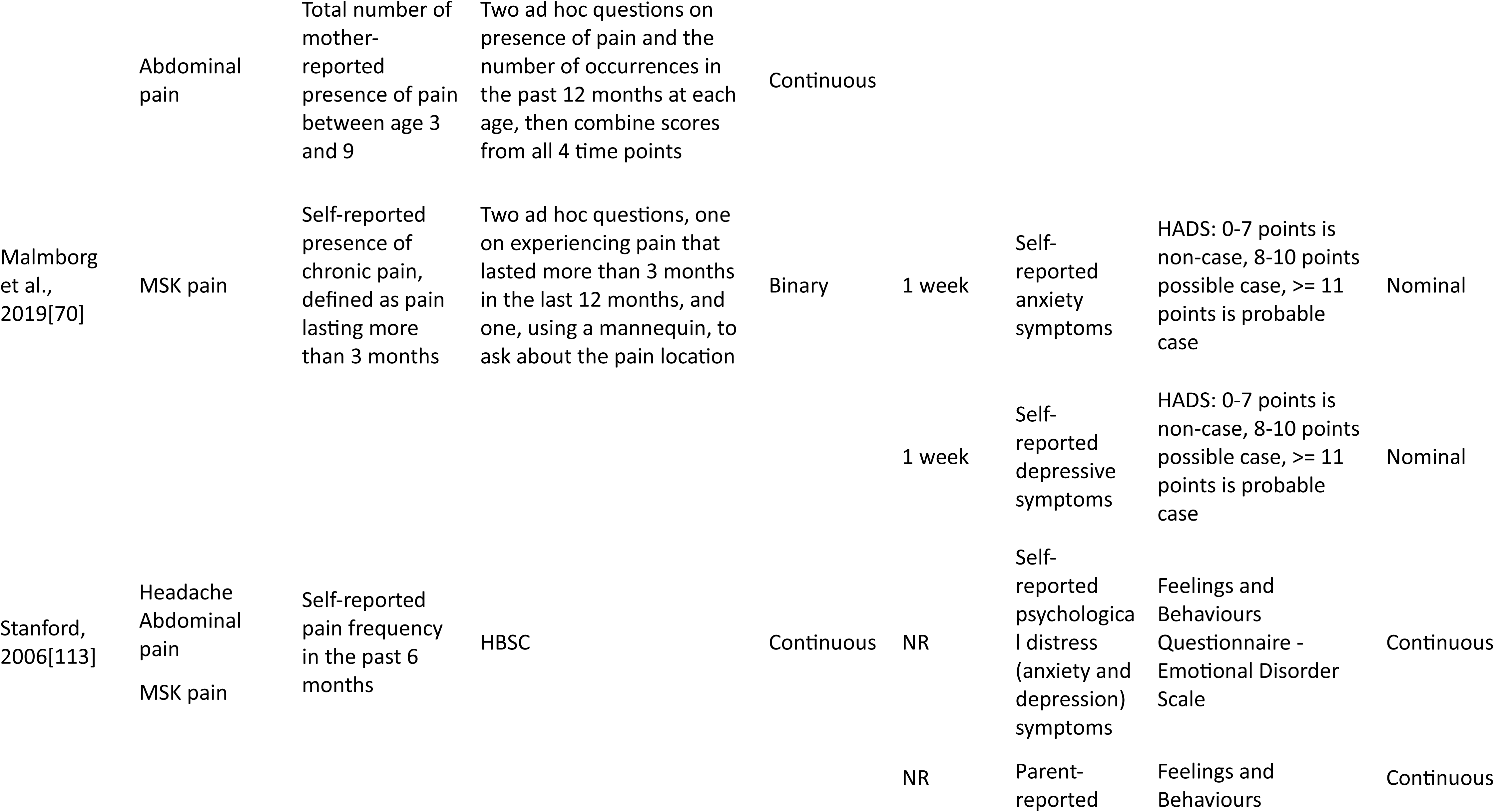
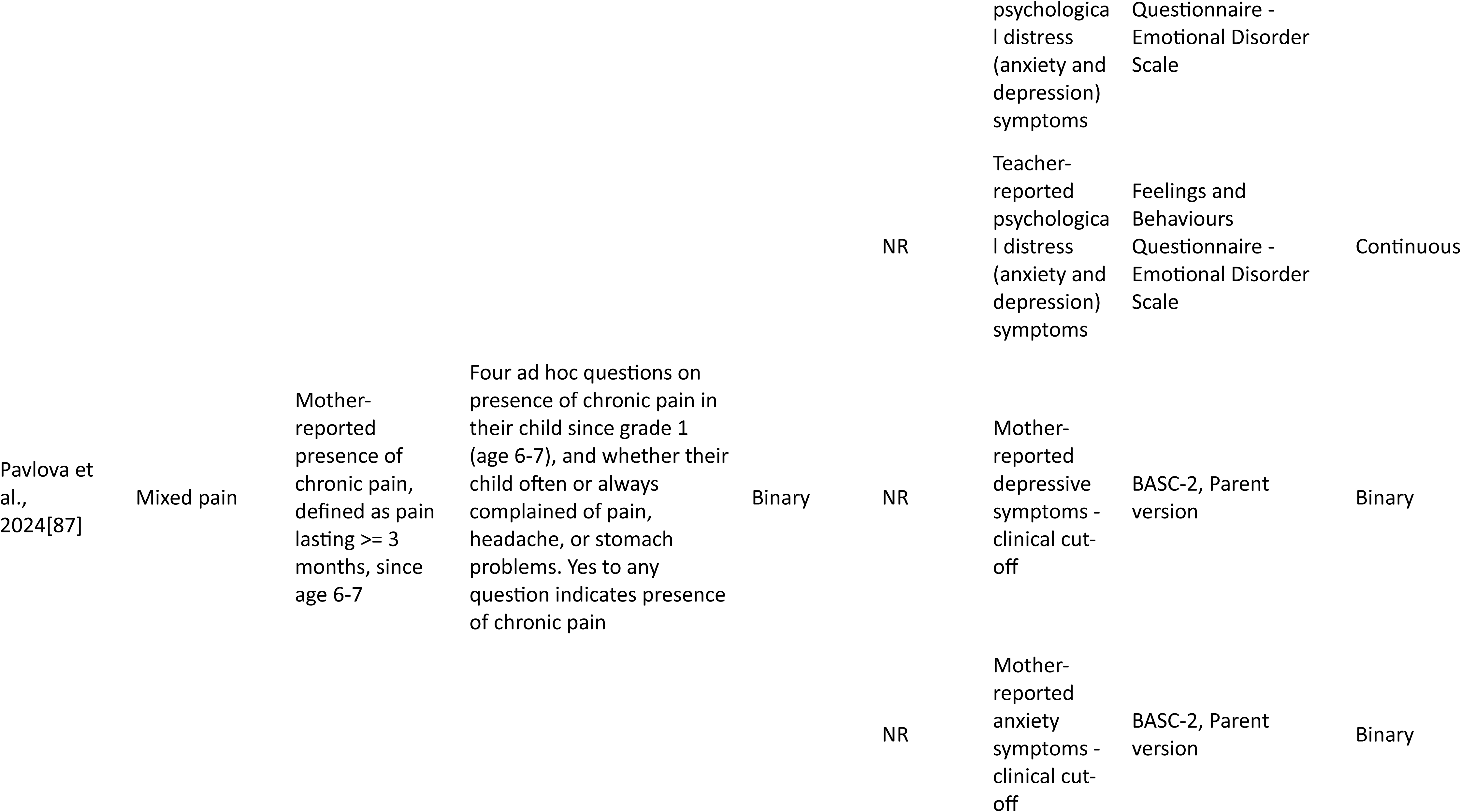
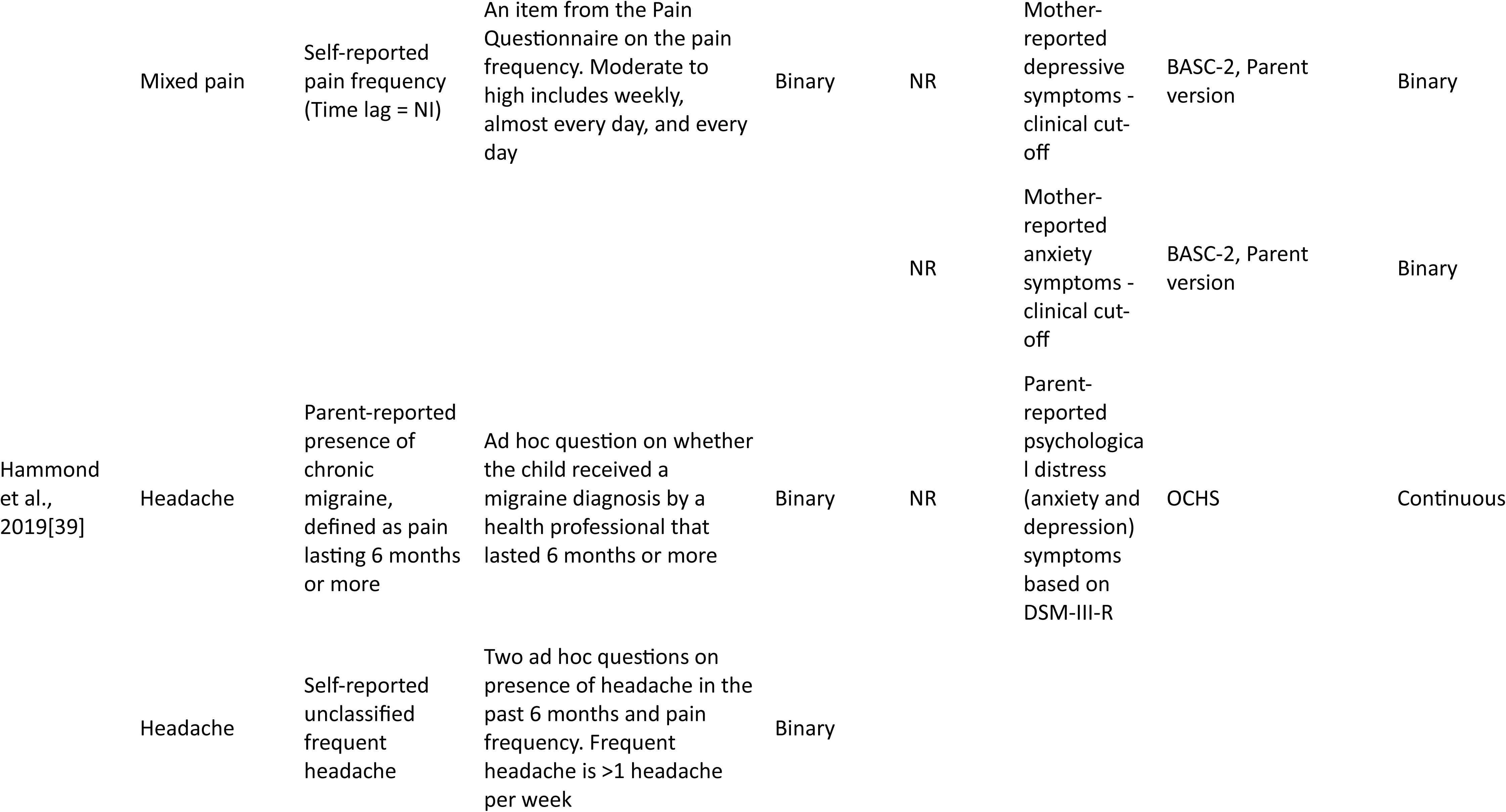
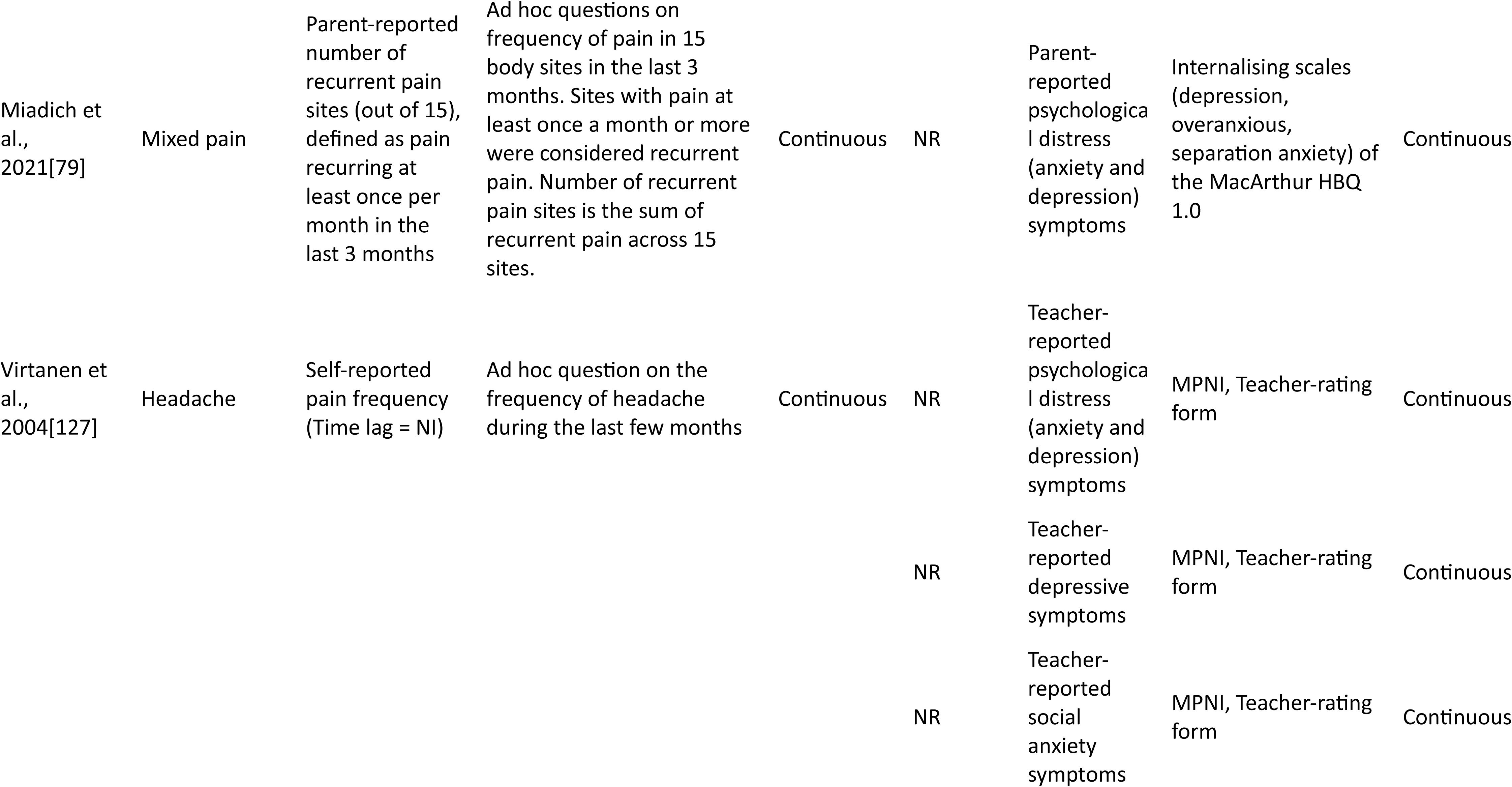
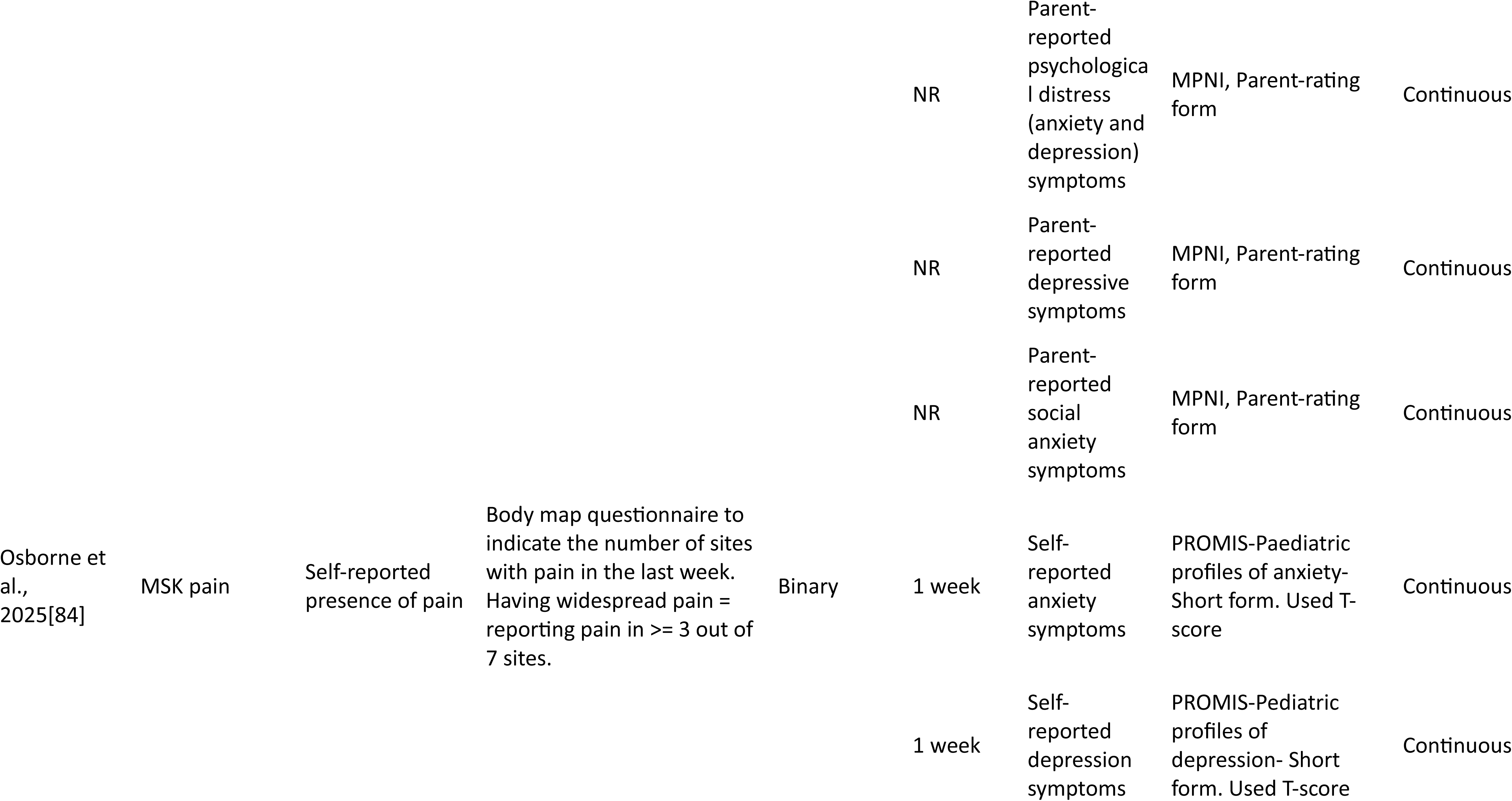
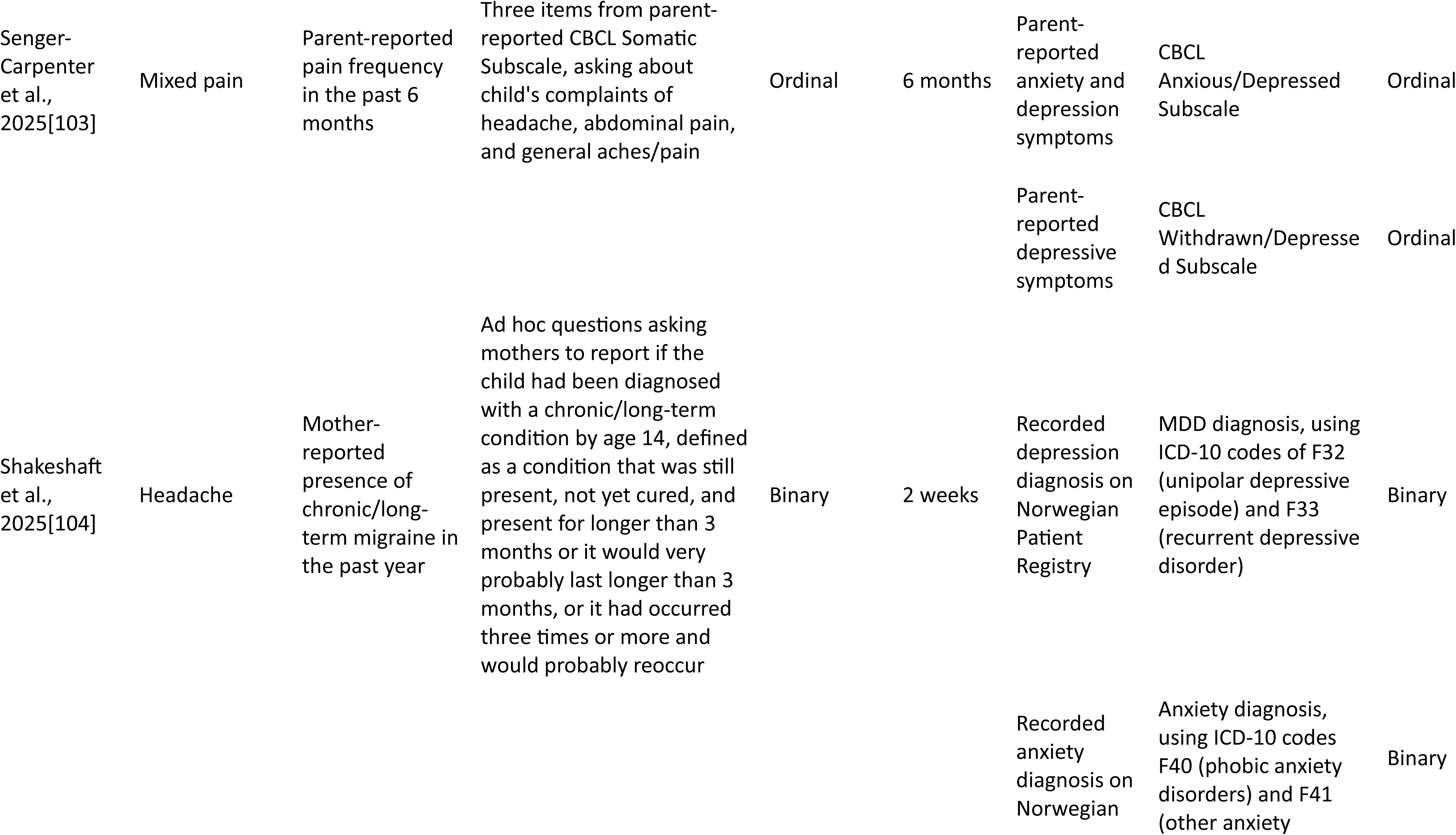
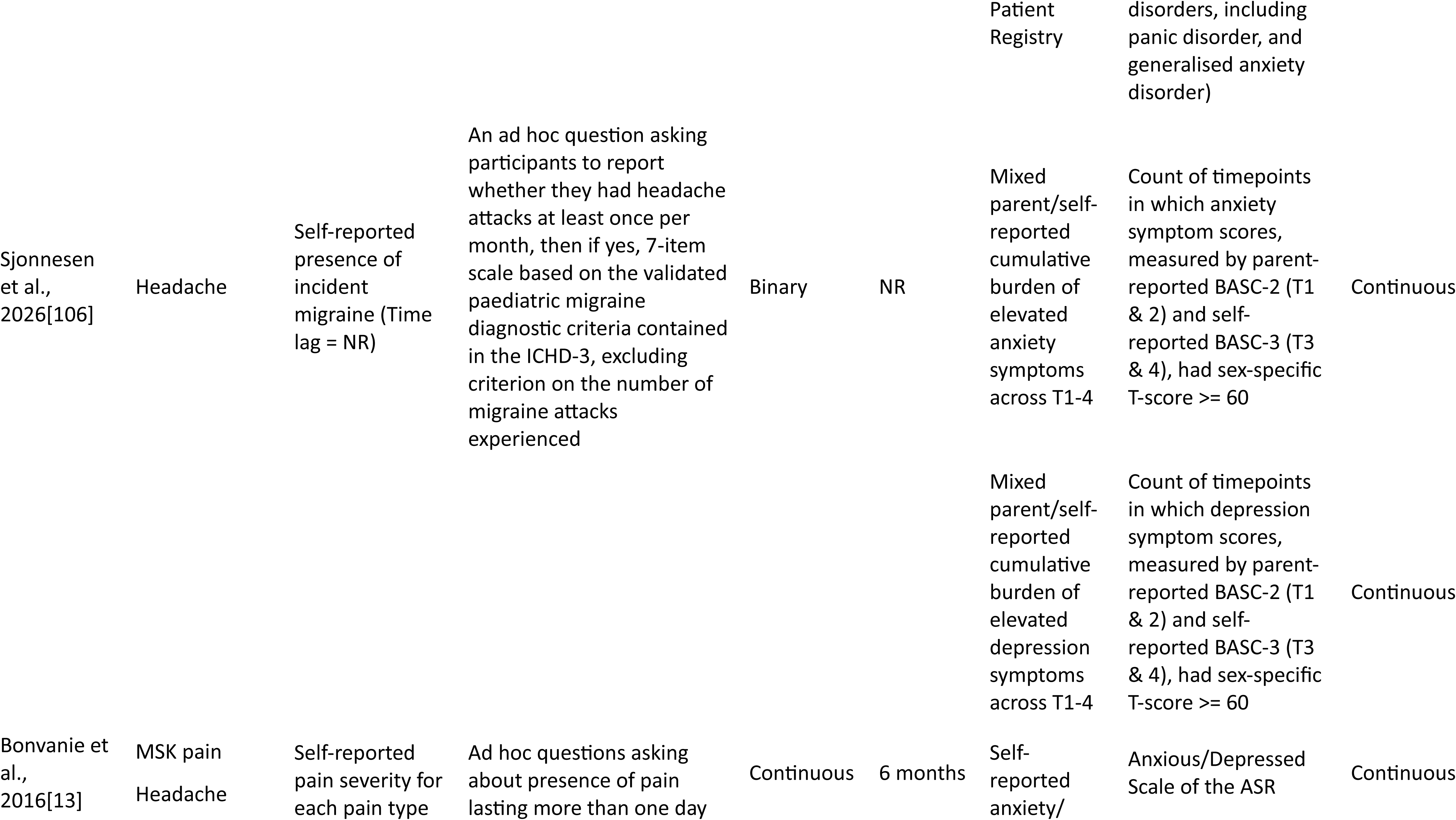
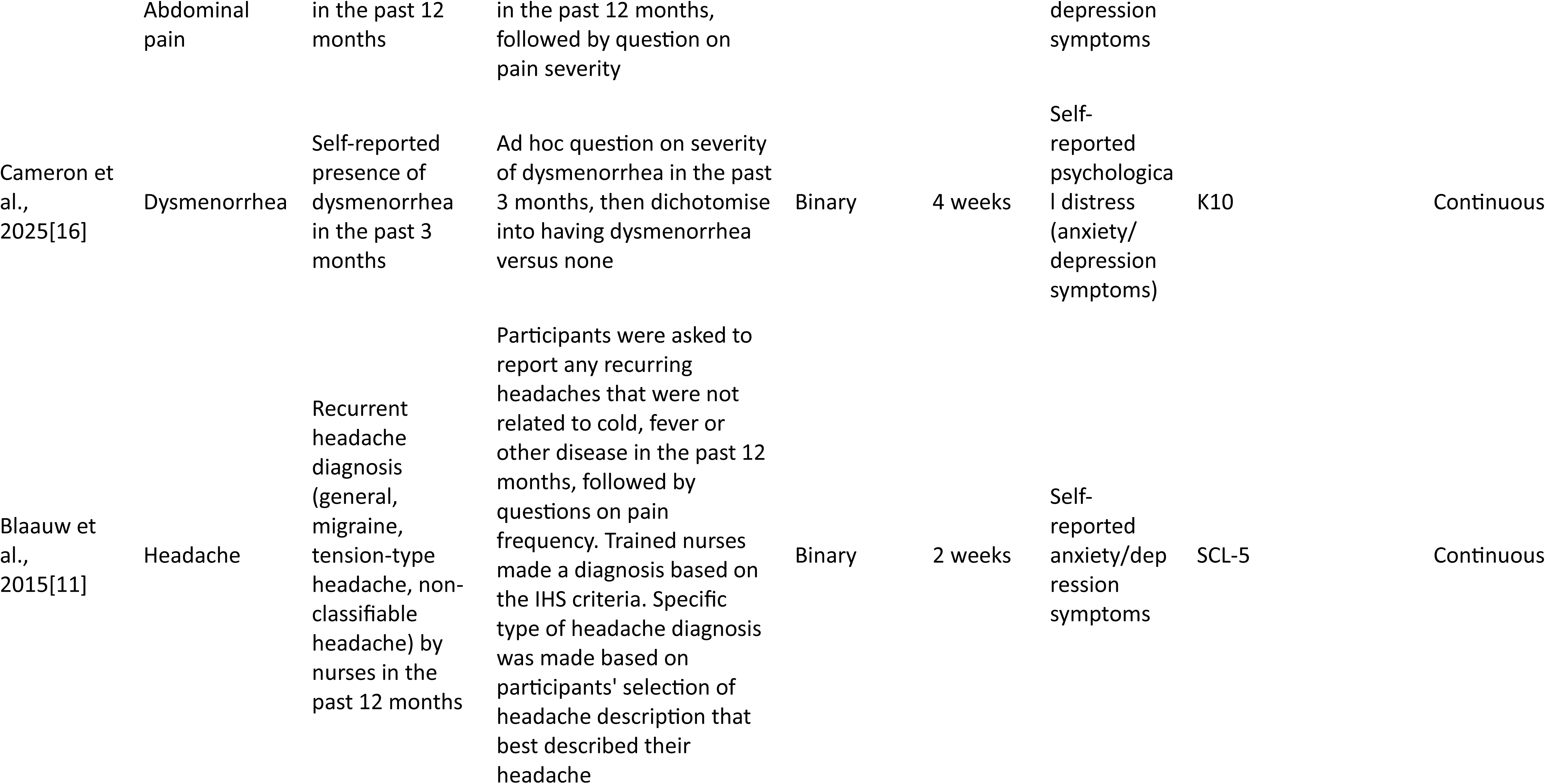
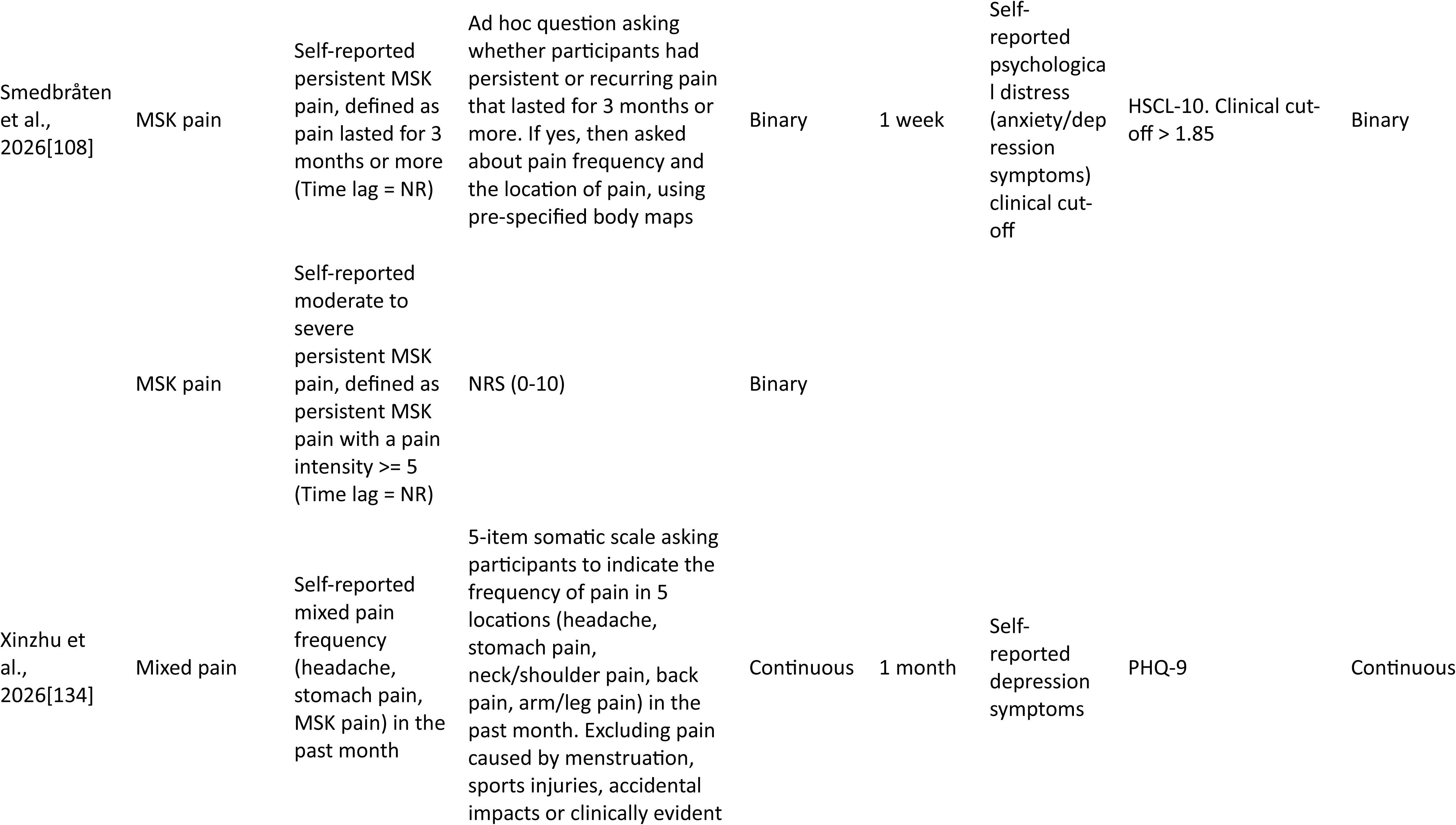
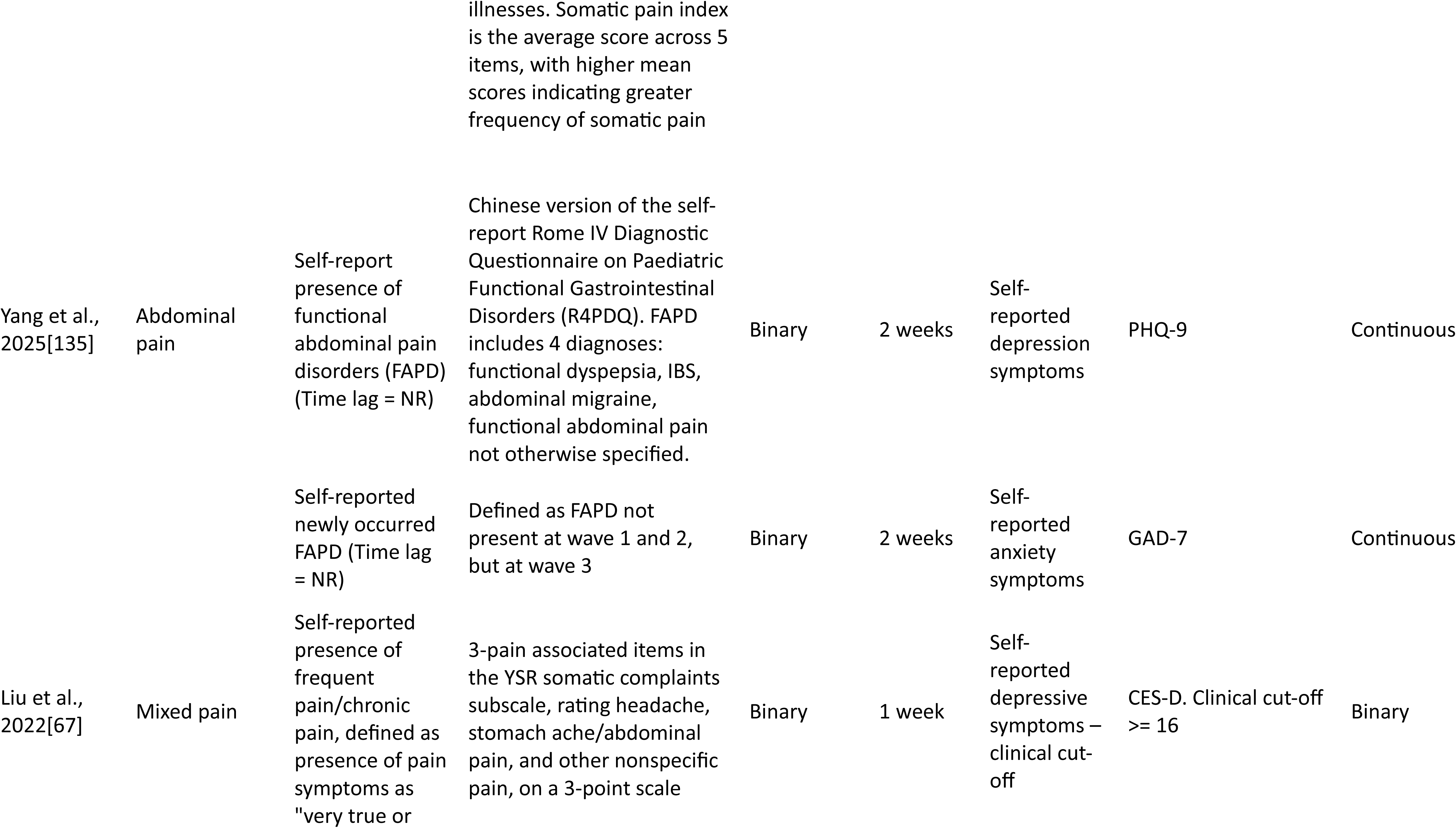
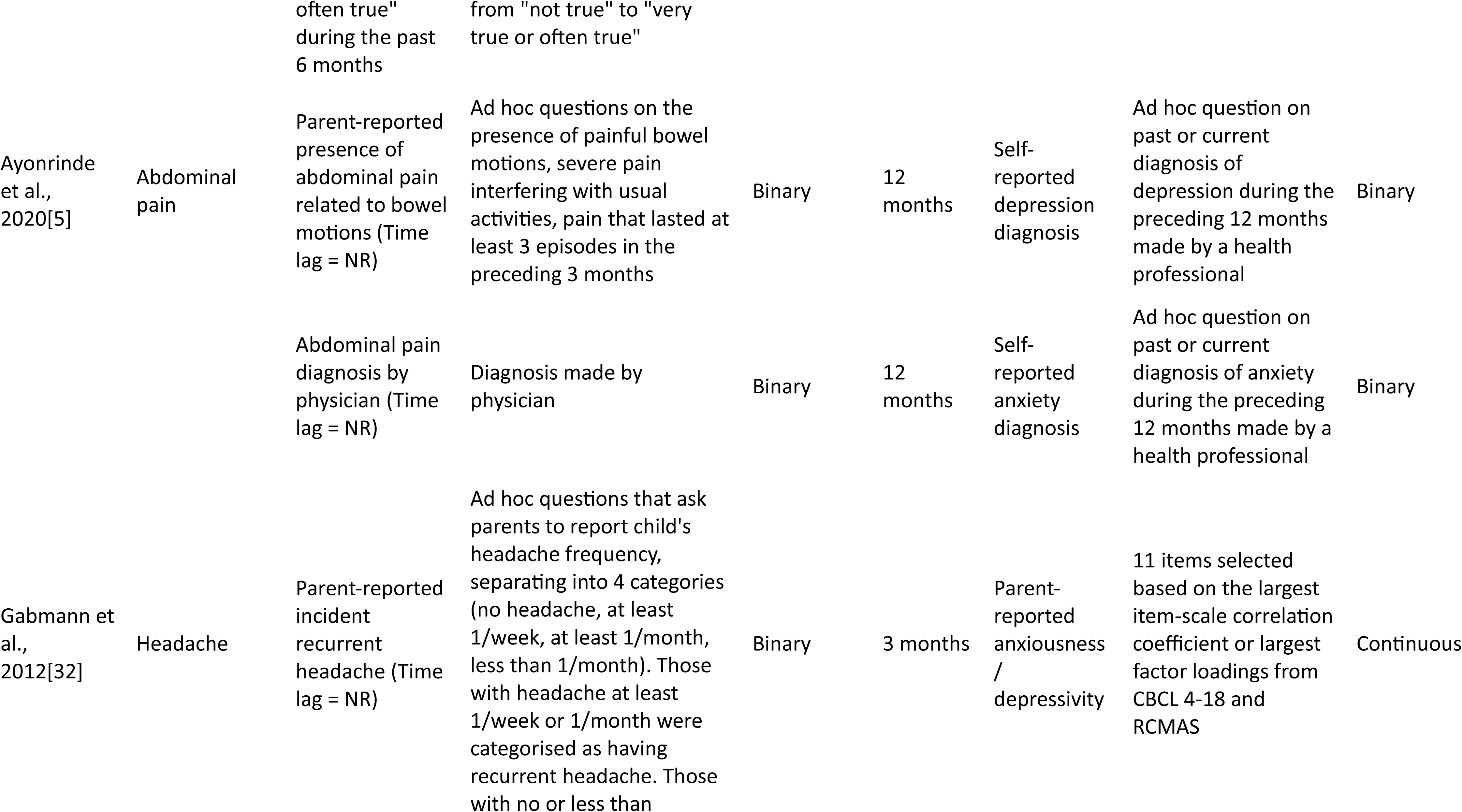
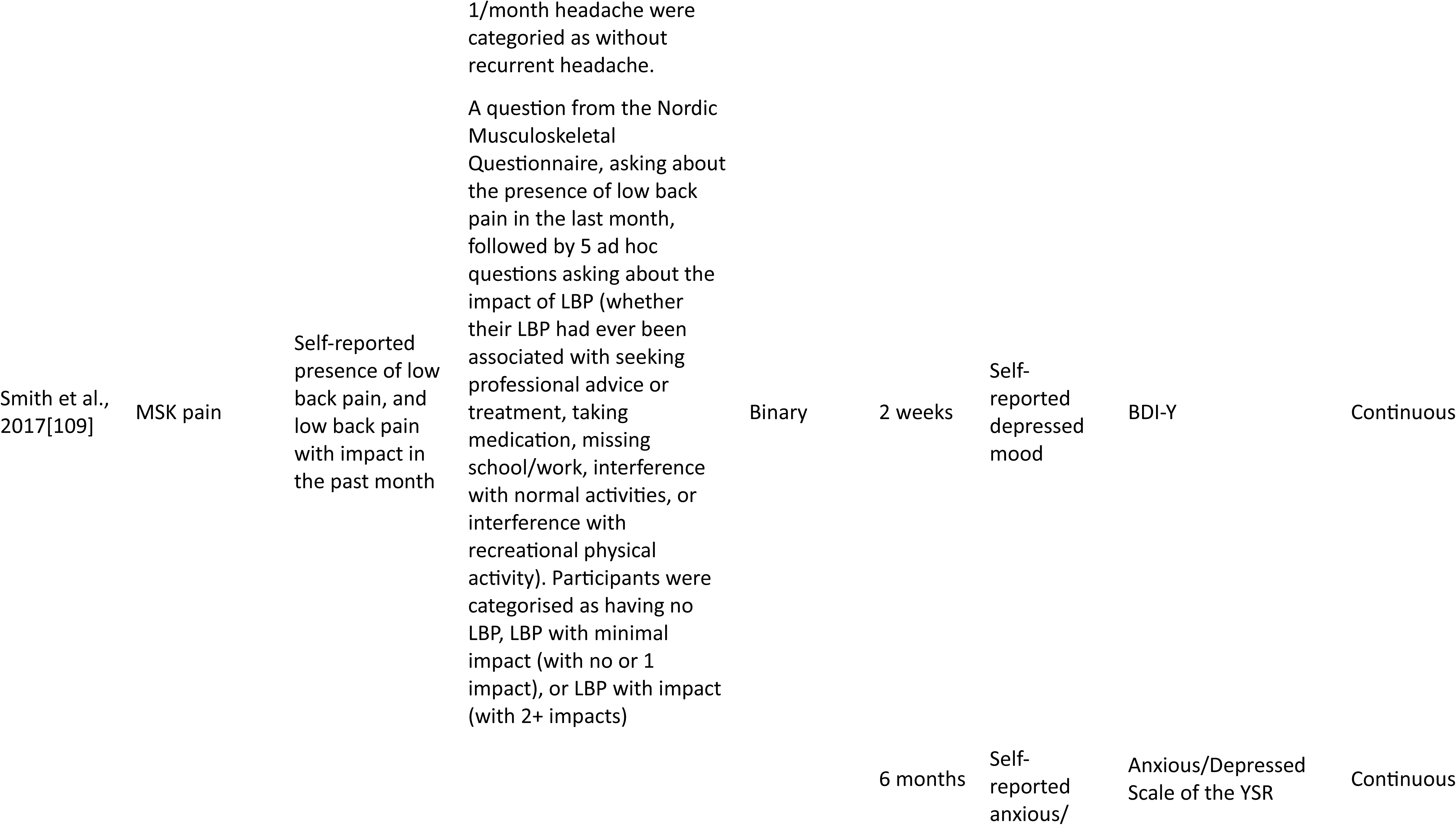
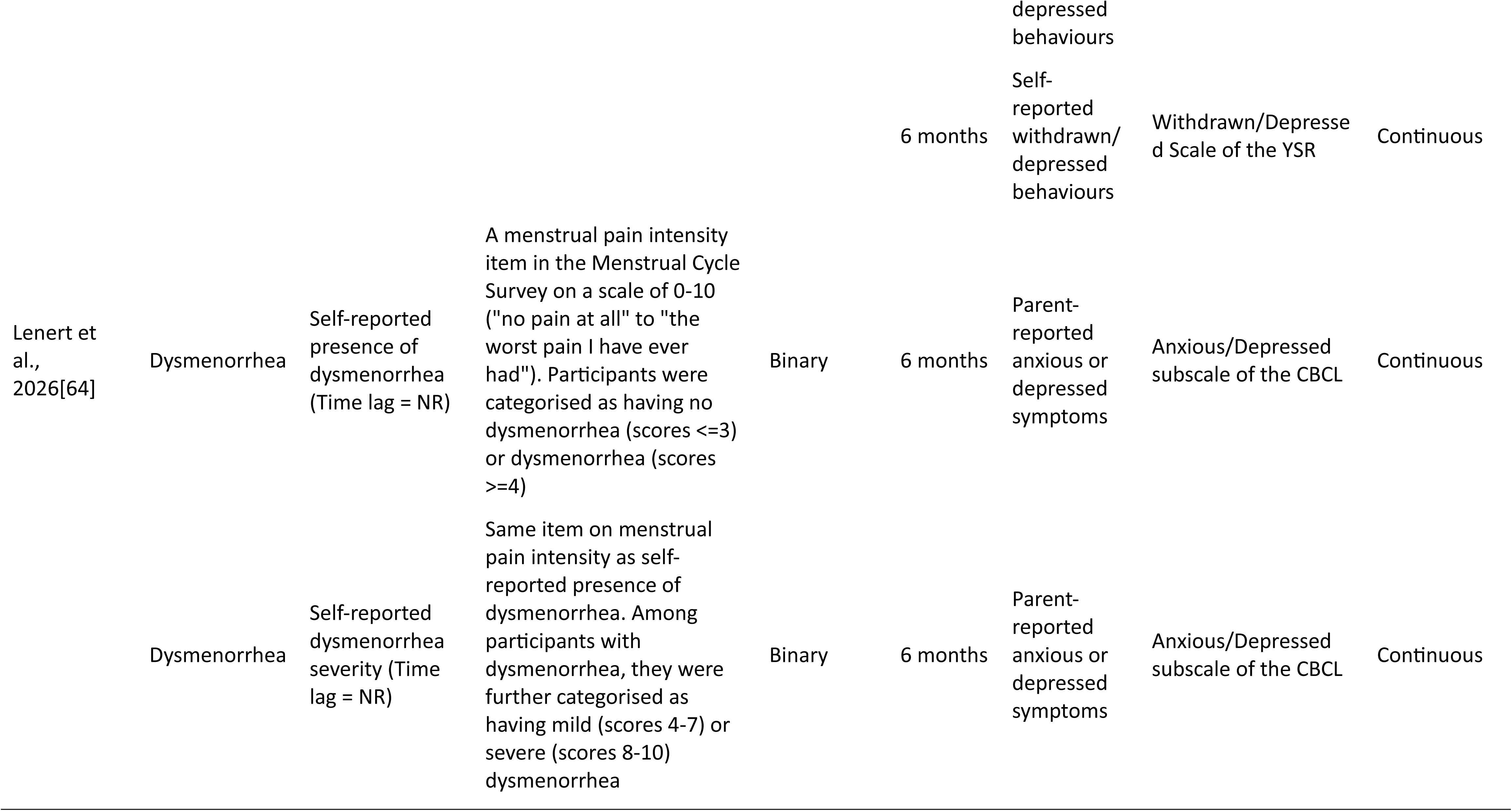
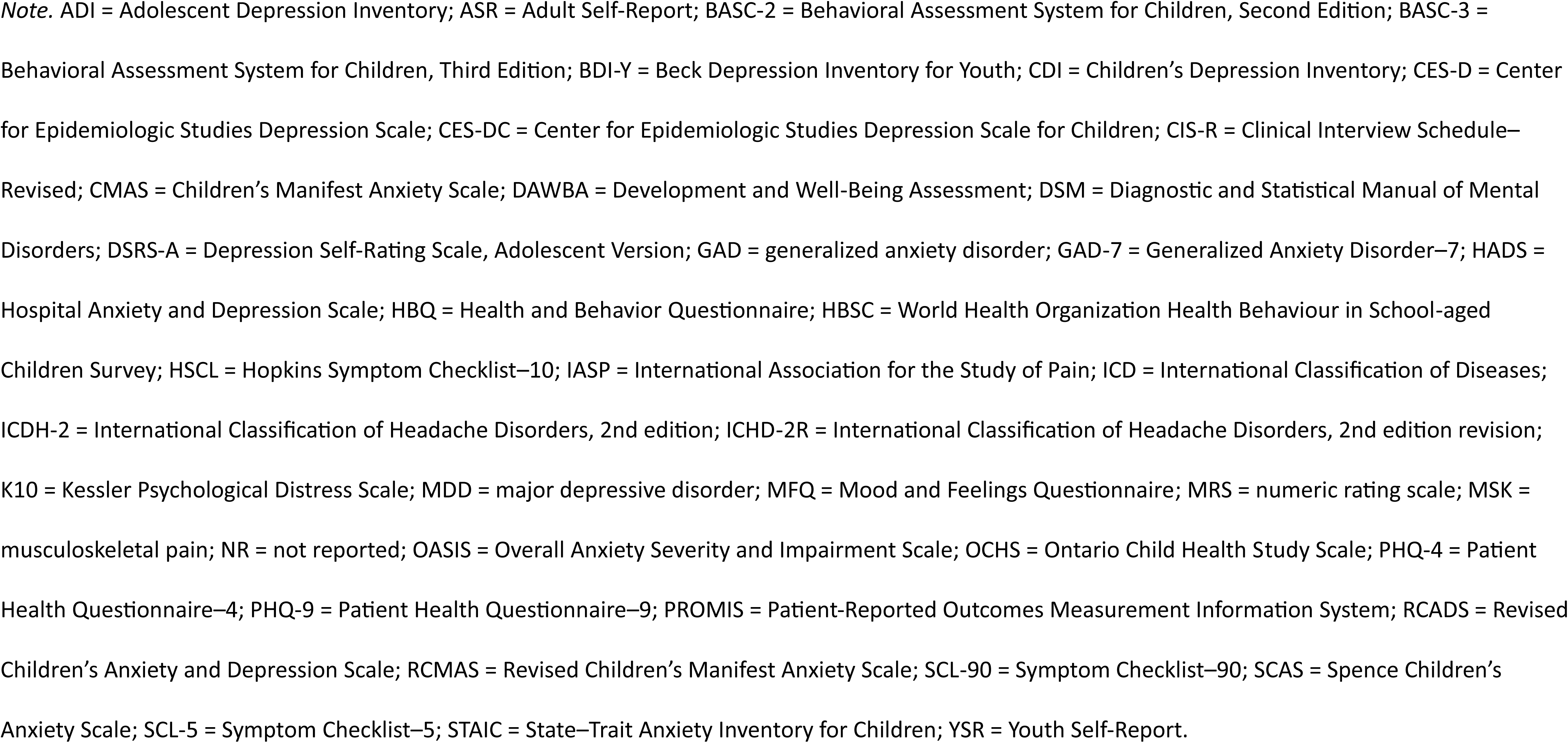
Definitions and measures used by included studies.

Similar variability was observed for anxiety and depression. They were conceptualised as binary diagnoses or clinical cut-offs (*k* = 13), continuous symptoms (*k* = 27), or nominal/ordinal indicators of frequency and severity (*k* = 2). They were typically reported by primary caregivers and/or teachers for children, and self-reported by adolescents and young adults.

### 3.4. Meta analysis

#### 3.4.1. Anxiety and subsequent pain

##### 3.4.1.1. Unadjusted association

Meta-analysis of 24 unadjusted associations from nine studies [4,6,84,98,106,106,121,122,127,135] showed that anxiety was significantly associated with subsequent pain (*n* = 15,087; *k* = 24; OR = 1.60; 95% CI: 1.19-2.15; *p* =.003; Figure 2A). There was no evidence of publication bias (Figure S1A, B; Peter’s test: *t(22)* = 0.63, *p* =.538). Heterogeneity was considerable (total *I^2^ =* 97.83%), and largely attributable to between-study variation (*I^2^_between_* = 95.12; *τ̂*^2^ = 0.1618; 95% CI: 0.0651-0.5383). The within-study variance was small (*I^2^_within_* = 2.72; *τ̂*^2^ = 0.0046; 95% CI: 0.0001-0.0220). The prediction interval crossed the null value (OR = 0.66-3.91), indicating greater uncertainty in the estimated effect. Meta-regressions were conducted with sex, pain type, risk of bias, anxiety informant, pain operationalisation, mean baseline age, and follow-up duration (Table S1). Associations were weaker when pain was operationalised as a categorical outcome and follow-up durations were longer. They were also weaker in males than females.

**Figure 2.**
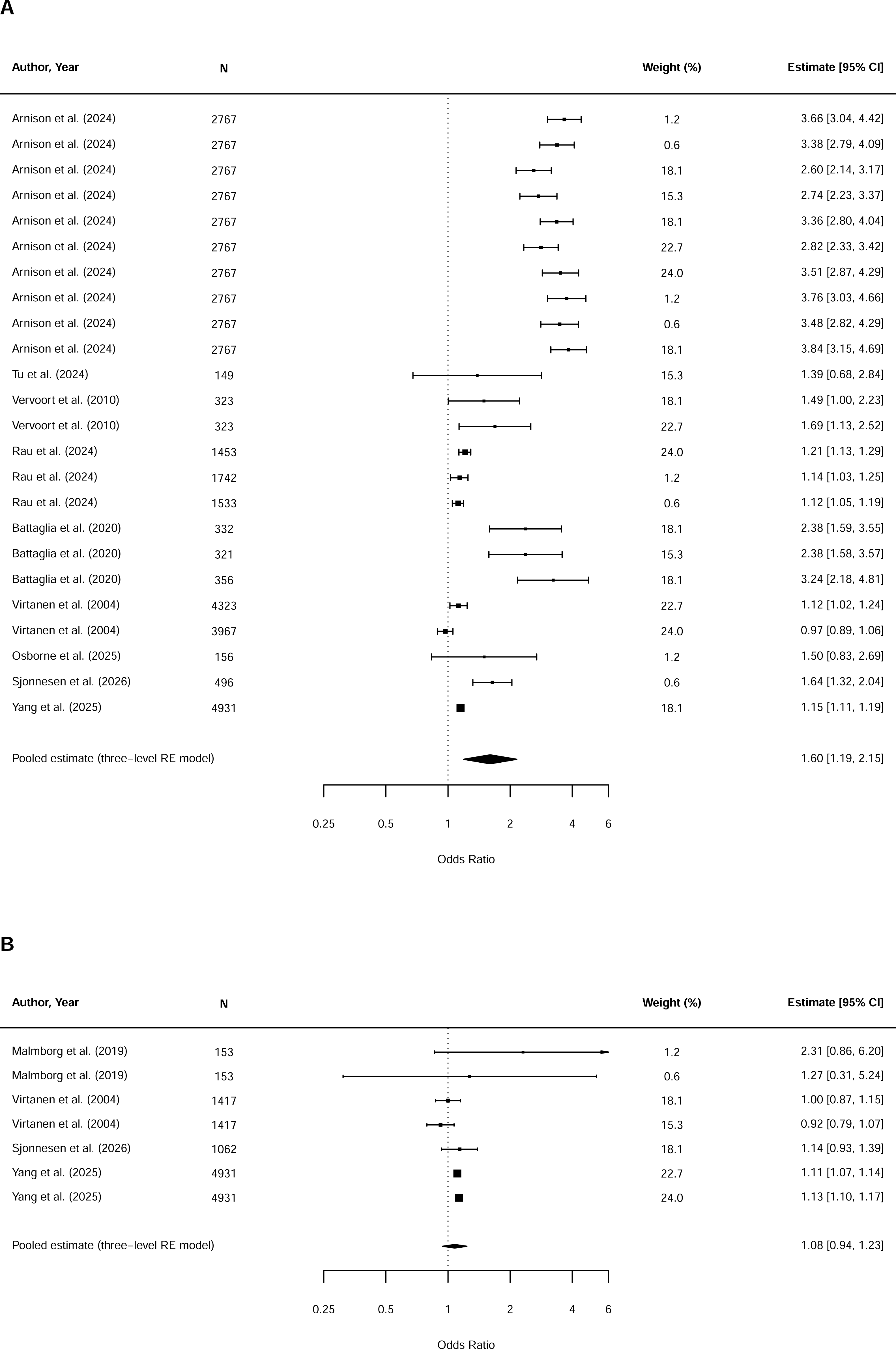
Forest plots of meta-analyses of prospective associations of anxiety and subsequent pain. (A) Unadjusted associations; (B) Adjusted associations.

##### 3.4.1.2. Adjusted association

Meta-analysis of seven adjusted associations from four studies with categorical pain [70,106,127,135] showed no significant association between anxiety and subsequent pain after adjustment for confounders (*n* = 7,563; *k* = 7; OR = 1.08; 95% CI: 0.94-1.23; *p* =.224; Figure 2B). There was no indication of publication bias (Figure S1C, D; Peter’s test: *t(5)* =-0.18, *p* =.864). Heterogeneity was considerable (total *I^2^ =* 83.18%), almost entirely due to between-study variation (*I^2^_between_ =* 83.18%; *τ̂*^2^ = 0.0064; 95% CI: 0.0000-0.1798). Within-study heterogeneity variance was negligible (*I^2^_within_ =* 0.00%; *τ̂*^2^ = 0.0000; 95% CI: 0.0000-0.0095). The prediction interval crossed null (OR = 0.85-1.37), reducing certainty in the pooled effect. Meta-regressions with mean baseline age, anxiety informant, anxiety operationalisation, pain type, and follow-up durations, found no significant moderators (Table S2).

#### 3.4.2. Depression and subsequent pain

##### 3.4.2.1. Unadjusted association

Meta-analysis of 28 unadjusted associations from 11 studies [4,34,68,84,98,106,109,121,127,134,134,135] showed a significant association between depression and subsequent pain in children and adolescents (*n* = 19,452; *k* = 28; OR = 1.88; 95% CI: 1.26-2.82; *p* =.003; Figure 3A). Peters’ test was significant (*t* =-2.17, *p* =.039), but the funnel plots suggested that the asymmetry was not due to small-study effects (Figure S2A, B). Heterogeneity was considerable (total *I^2^* = 99.71%) and was largely due to between-study variation (*I^2^_between_* = 99.68%; *τ̂*^2^ = 0.3991; 95% CI: 0.1774-1.1434). Within-study heterogeneity was minimal (*I^2^_within_* = 0.03%; *τ̂*^2^ = 0.0001; 95% CI: 0.0000-0.0033). The prediction interval crossed the null value (OR = 0.48-7.32), reducing certainty around estimated effect. Meta-regressions were conducted with sex, mean baseline age, follow-up duration, depression informant, pain informant, depression operationalisation, pain operationalisation, pain type, and risk of bias. For depression informant, one study [106] used a combined self-and parent-report measure and therefore was excluded from the meta-regression (Table S3). Results showed that being male, categorical pain, and longer follow-up durations were associated with weaker associations.

**Figure 3.**
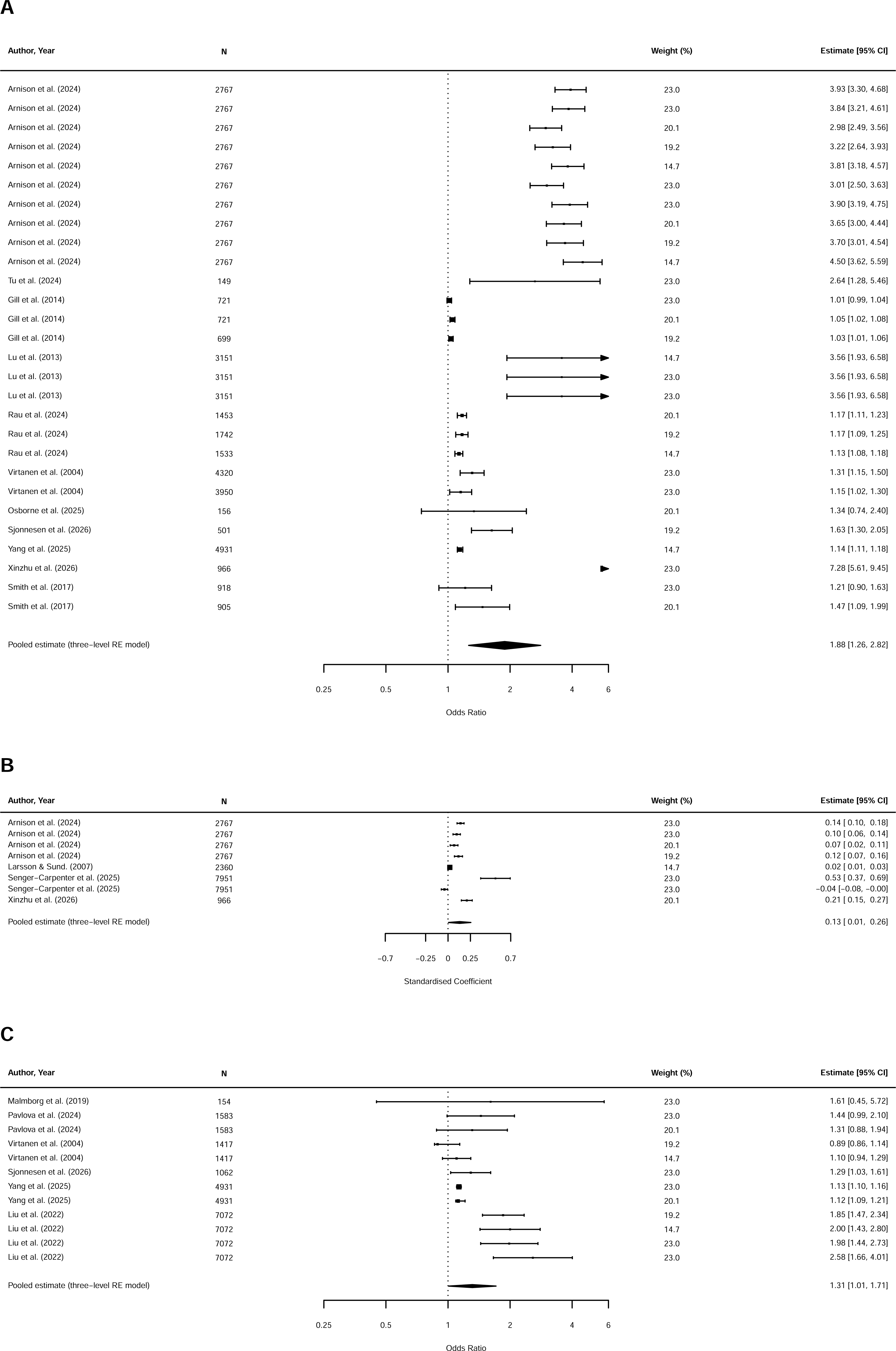
Forest plots of meta-analyses of prospective associations of depression and subsequent pain. (A) Unadjusted associations; (B) Adjusted associations of continuous pain outcomes; (C) Adjusted associations of categorical pain outcomes.

##### 3.4.2.2. Adjusted association

Two meta-analyses were conducted. Meta-analysis with continuous pain outcomes was conducted with eight associations from four studies [4,62,103,134]. It showed depression was significantly associated with subsequent pain symptoms, after adjusting for confounders (*n* = 14,044; *k* = 8; β = 0.13; 95% CI: 0.01-0.26, *p* =.043; Figure 3B). There was no evidence of publication bias (Figure S2C, D; Peters’ test: *t(6)* = 0.93, *p* =.389). Heterogeneity was considerable (total *I^2^* = 98.65%), almost entirely due to within-study variation (*I^2^_within_* = 98.65%; *τ̂*^2^ = 0.0212; 95% CI: 0.0062-0.0878). Between-study heterogeneity was negligible (*I^2^_between_* = 0.00%; *τ̂*^2^= 0.0000; 95% CI: 0.0000-0.0646). The prediction interval crossed the null threshold (β =-0.24-0.50), suggesting greater uncertainty in the estimated effect. Meta-regression with sex, mean baseline age, depression informant, pain informant, pain type, and risk of bias showed no significant moderation (Table S4).

For studies of categorical pain outcomes, meta-analysis of 12 associations from six studies [67,70,87,106,127,135] also showed a significant association between depression and subsequent pain after adjustment for confounders (*n* = 16,219; *k* = 12; OR = 1.31; 95% CI: 1.01-1.71; *p* =.045; Figure 3C). There was no indication of publication bias (Figure S2E, F; Peters’ test: *t(10)* =-0.40, *p* =.700). Heterogeneity was considerable (total *I^2^* = 95.20%), almost all explained by between-study variation(*I^2^_between_*= 95.20%; *τ̂*^2^ = 0.0668; 95% CI: 0.0189-0.3785). Within-study heterogeneity was negligible (*I^2^_within_* = 0.00%, *τ̂*^2^ = 0.0000; 95% CI: 0.0000-0.0188). The prediction interval crossed the null value (OR = 0.70-2.46), decreasing certainty around the estimated effect. Meta-regressions were conducted with mean baseline age, age group, depression informant, depression operationalisation, pain informant, pain type, risk of bias, and follow-up duration (Table S5). Associations were weaker in studies with categorical depression.

#### 3.4.3. Combine Anxiety and depression and subsequent pain

##### 3.4.3.1. Unadjusted association

Meta-analysis of ten unadjusted associations from seven studies (Lenert et al., 2026; Lien et al., 2011; Miadich et al., 2021; Smedbråten et al., 2022; Smith et al., 2017; Virtanen et al., 2004; Wolff et al., 2012) showed a significant association between combined depression and anxiety and subsequent pain in children and adolescents (*n* = 13,352; *k* = 10; OR = 1.32; 95% CI: 1.15-1.51, *p* =.002; Figure 4A). There was no indication of publication bias (Figure S3A, B; Peters’ test: *t(8)* = 2.27, *p* =.053). Heterogeneity was moderate (total *I^2^ =* 55.15%), nearly all attributable to within-study variation (*I^2^_within_* = 55.15%; *τ̂*^2^ = 0.0175; 95% CI: 0.0000-0.0962). Between-study heterogeneity was negligible (*I^2^_between_* = 0.00%; *τ̂*^2^ = 0.0000; 95% CI: 0.0000-0.1651). The prediction interval crossed null (OR = 0.95-1.83), indicating greater uncertainty in the estimated effect. Meta-regressions were conducted with mean baseline age, age group, follow-up duration, anxiety/depression informant, pain type, pain informant, and risk of bias (Table S6). The results showed longer follow-up duration was significantly associated with weaker associations.

**Figure 4.**
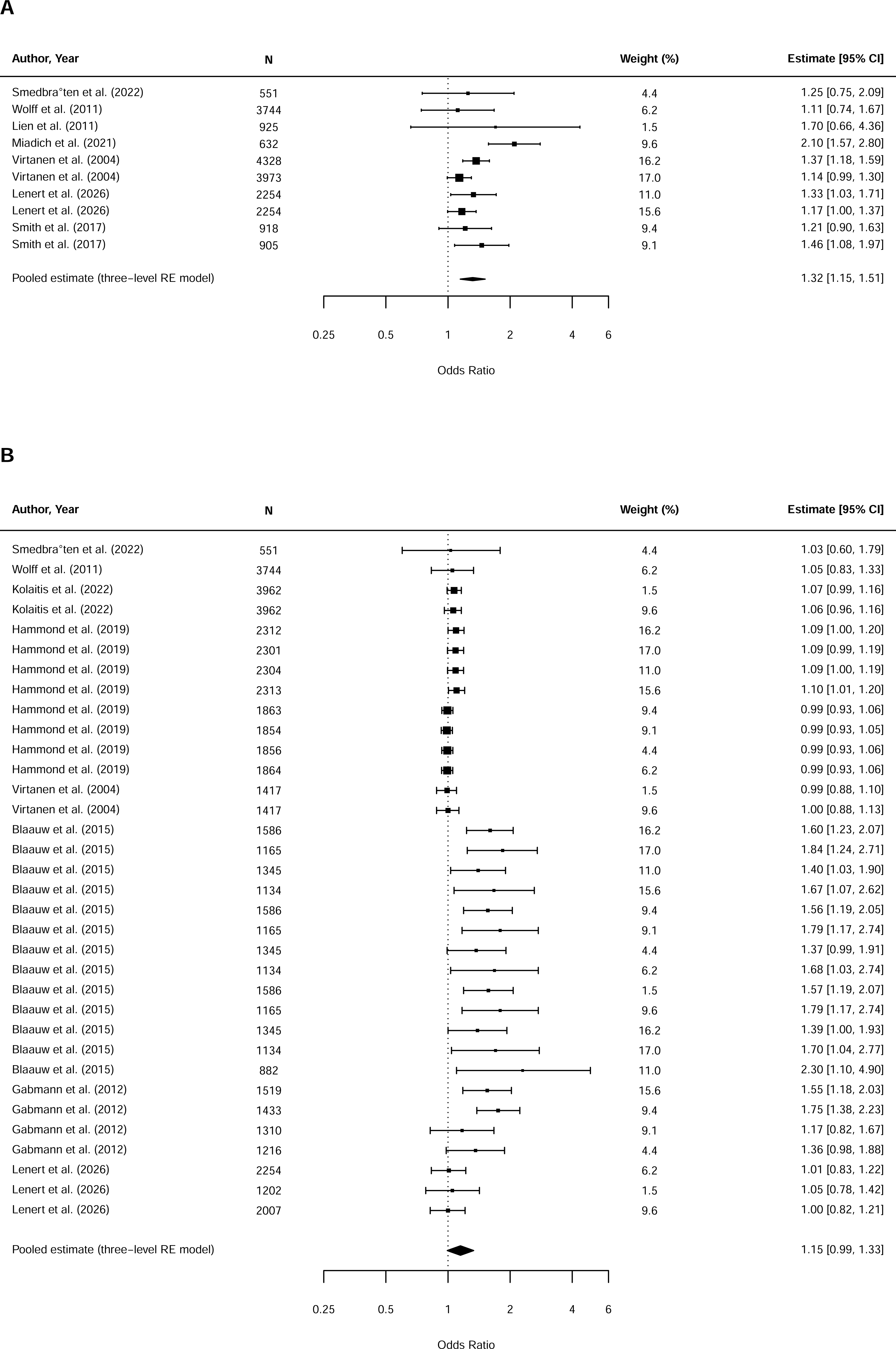
Forest plots of meta-analyses of prospective associations of combined anxiety/depression and subsequent pain. (A) Unadjusted associations; (B) Adjusted associations.

##### 3.4.3.2. Adjusted association

Meta-analysis of 34 associations from eight studies with categorical pain [11,32,39,61,64,107,127,130] showed no significant association between combined anxiety/depression with subsequent pain, after adjustment for confounders (*n* = 17,346; *k* = 34; OR = 1.15; 95% CI: 0.99-1.33; *p* =.063; Figure 4B). There was no indication of publication bias (Figure S3C, D; Peters’ test: *t(32)* = 1.30; *p* =.203). Heterogeneity was considerable (total *I^2^ =* 89.05%), primarily due to between-study variation (*I^2^_between_ =* 87.82%; *τ̂*^2^ = 0.0344; 95% CI: 0.0116-0.1364). Within-study heterogeneity was small (*I^2^_within_ =* 1.23%; *τ̂*^2^ = 0.0005; 95% CI: 0.0000-0.0040). The prediction interval included the null value (OR = 0.76-1.73), indicating greater uncertainty in the estimated effect. Meta-regressions were conducted with mean baseline age, age group, follow-up duration, pain informant, pain type, risk of bias, and anxiety/depression informant (Table S7). Studies with high risk of bias reported significantly stronger associations than those with low risk of bias. Looking at the results of studies with low risk of bias, the association remained non-significant.

#### 3.4.4. Pain and subsequent anxiety

##### 3.4.4.1. Unadjusted association

Meta-analysis of 22 unadjusted associations from four studies [4–6,97] showed that pain was significantly associated with subsequent anxiety in children and adolescents (*n* = 11,529; *k* = 22; OR = 2.78, 95% CI: 2.46-3.13, *p* <.001; Figure 5A). There was no evidence of publication bias (Figure S4A, B; Peters’ test: *t(20)* = 0.16, *p* =.872). Heterogeneity was substantial (total *I^2^ =* 54.37%), mostly attributable to within-study variation (*I^2^_within_* = 49.68%; *τ̂*^2^ = 0.0201; 95% CI: 0.0040-0.0749). Between-study heterogeneity accounted for a small proportion of the total variance (*I^2^_between_* = 4.69%; *τ̂*^2^ = 0.0019; 95% CI: 0.0000-0.1741). The prediction interval range did not cross null threshold (OR = 1.99-3.86), increasing certainty in the estimated effect. Meta-regressions with mean baseline age, follow-up duration, pain type, risk of bias, age group, pain informant, pain operationalisation, anxiety operationalisation, and sex found that the associations were significantly weaker in studies with younger age at baseline, longer follow-up duration, and males (Table S8).

**Figure 5.**
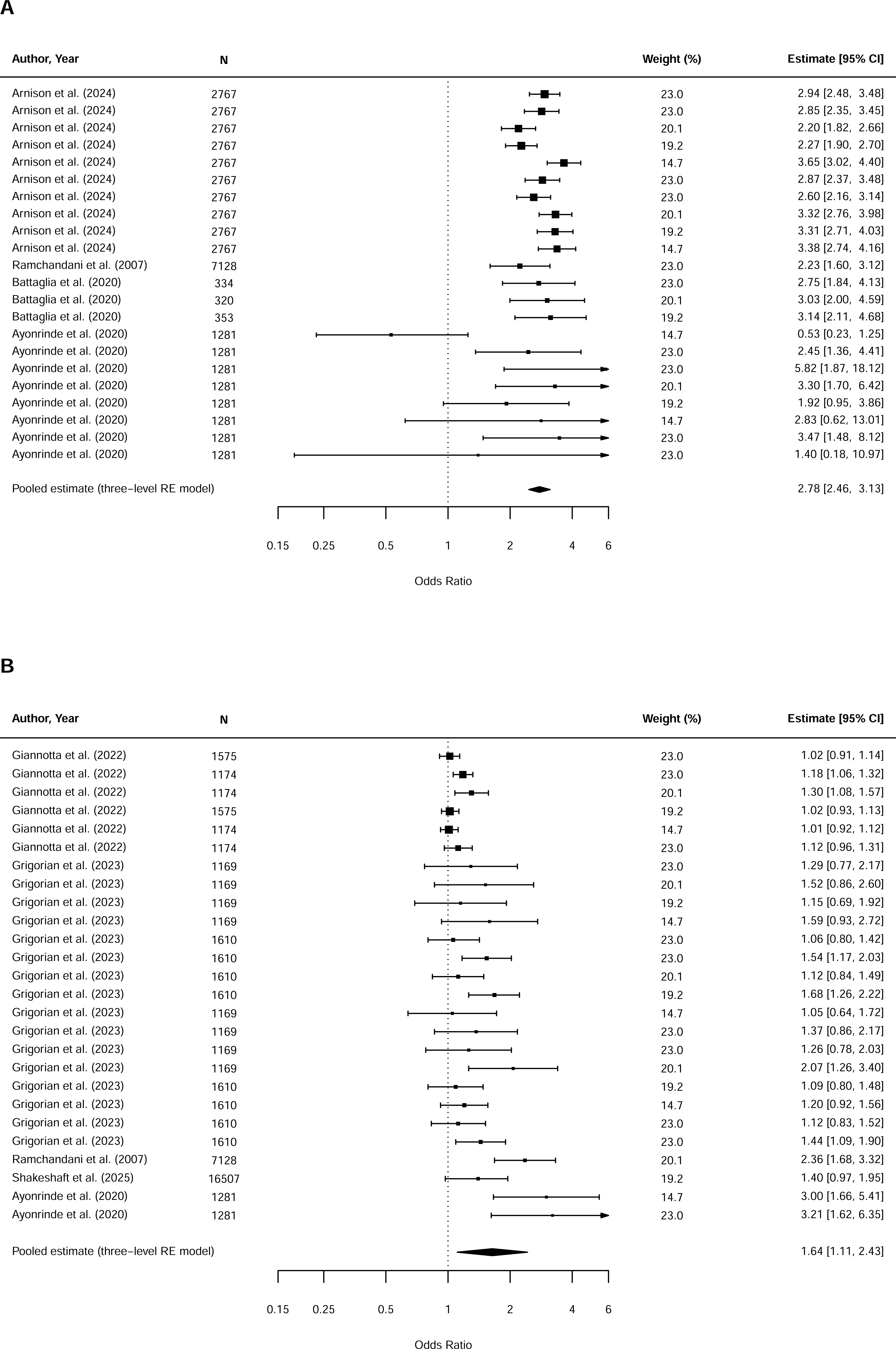
Forest plots of meta-analyses of prospective associations of pain and subsequent anxiety. (A) Unadjusted associations; (B) Adjusted associations.

##### 3.4.4.2. Adjusted association

Meta-analysis of 26 associations from five studies with categorical anxiety outcomes [5,33,36,97,104] showed pain was significantly associated with subsequent anxiety, after adjustment for confounders (*n* = 28,101; *k* = 26; OR = 1.64; 95% CI: 1.11-2.43; *p* =.016; Figure 5B). There was no indication of publication bias (Figure S4C, D; Peters’ test: *t(24)* =-1.15; *p* =.262). Heterogeneity was considerable (total *I^2^ =* 92.93%), mostly due to between-study variation (*I^2^_between_ =* 90.51%; *τ̂*^2^ = 0.1581; 95% CI: 0.0329-1.1013). Within-study heterogeneity was small (*I^2^_within_ =* 2.42%; *τ̂*^2^= 0.0042; 95% CI: 0.0000-0.0245). The prediction interval crossed null (OR = 0.66-4.11), indicating greater uncertainty in the estimated effect. Meta-regressions were conducted with mean baseline age, follow-up duration, age group, pain informant, pain operationalisation, risk of bias, and anxiety informant (Table S9). Results showed that the associations were significantly weaker in adolescents, compared to children, and in studies that used youth-reported instead of other-reported pain.

#### 3.4.5. Pain and subsequent depression

##### 3.4.5.1. Unadjusted association

Meta-analysis of 26 unadjusted associations from five studies [4,5,48,97,134] showed that pain was significantly associated with subsequent depression in children and adolescents (*n* = 18,566; *k* = 26; OR = 2.48; 95% CI: 1.55-3.98; *p* <.001; Figure 6A). Significant Peters’ test (*t(24)* = 3.19, *p =*.004) suggested funnel plot asymmetry, but the funnel plots showed it was not due to small-study effects (Figure S5A, B). Heterogeneity was considerable (total *I^2^ =* 92.72%), primarily explained by between-study variation (*I^2^_between_* = 84.02%; *τ̂*^2^= 0.2164; 95% CI: 0.0391-1.5710). Within-study heterogeneity was small (*I^2^_within_* = 8.70%; *τ̂*^2^ = 0.0224; 95% CI: 0.0045-0.0844). The prediction interval crossed the null value (OR = 0.82-7.54), indicating less certainty in the estimated effect. Meta-regressions were conducted with mean baseline age, follow-up duration, pain type, depression operationalisation, age group, pain informant, pain operationalisation, risk of bias, and sex (Table S10). The associations were weaker when participants were younger at baseline, follow-up duration was longer, and depression was operationalised as categorical outcome.

**Figure 6.**
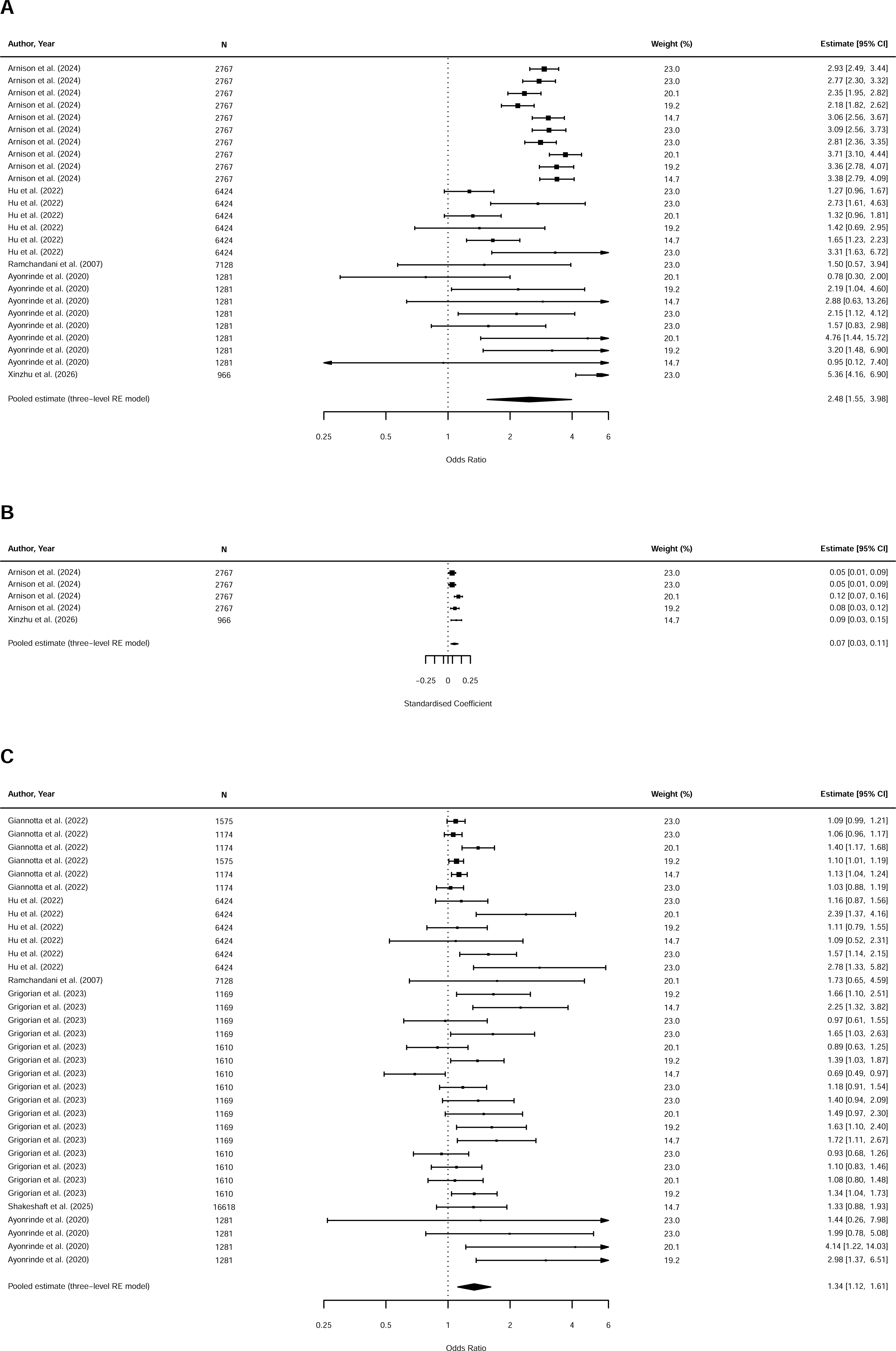
Forest plots of meta-analyses of prospective associations of pain and subsequent depression. (A) Unadjusted associations; (B) Adjusted associations of continuous depression outcomes; (C) Adjusted associations of categorical depression outcomes.

##### 3.4.5.2. Adjusted association

Two meta-analyses were conducted. Meta-analysis of five associations from two studies [4,134] showed that pain was significantly associated with more elevated depressive symptoms subsequently (*n* = 3733; *k* = 5; β = 0.07; 95% CI: 0.03-0.11; *p* =.007; Figure 6B). There was no evidence of publication bias (Figure S5C, D; Peters’ test: *t* =-0.68; *p* =.503). Heterogeneity was moderate (total *I^2^ =* 47.81%), nearly all due to within-study variation (*I^2^_within_* = 47.81%; *τ̂*^2^ = 0.0005; 95% CI: 0.0000-0.0054). Between-study heterogeneity variance was negligible (*I^2^_between_* = 0.00%; *τ̂*^2^ = 0.0000; 95% CI: 0.0000-0.0535). The prediction interval does not cross the null value (β = 0.00-0.15), increasing certainty in the estimated effect. Meta-regression was conducted with sex only and found no significant moderation (Table S11).

Meta-analysis of 34 associations from six studies with categorical depression [5,33,36,48,97,104] also showed pain was significantly associated with subsequent depression (*n* = 34,636; *k* = 34; OR = 1.34; 95% CI: 1.12-1.61; *p* =.003; Figure 6C). There was no evidence of publication bias (Figure S5E, F; Peters’ test: *t(32)* =-0.68; *p* =.503). Heterogeneity was considerable (total *I^2^ =* 77.44%), with majority attributed to between-study variation (*I^2^_between_* = 43.16%; *τ̂*^2^ = 0.0253; 95% CI: 0.0000-0.3345). Within-study heterogeneity variance was smaller (*I^2^_within_* = 34.28%; *τ̂*^2^ = 0.0201; 95% CI: 0.0000-0.0722). The prediction interval crossed the null value (OR = 0.84-2.15), reducing certainty in the estimated effect. Meta-regressions were conducted with mean baseline age, follow-up duration, pain type, risk of bias, age group, depression informant, pain informant, and pain operationalisation, but none were significant (Table S12).

#### 3.4.6. Pain with subsequent combined anxiety and depression

##### 3.4.6.1. Unadjusted association

Meta-analysis of 23 associations from four studies [16,66,108,114] showed that pain was significantly associated with subsequent combined anxiety/depression (*n* = 6,478; *k* = 23; OR = 1.89; 95% CI: 1.46-2.45; *p* <.001; Figure 7A). There was no indication of publication bias (Figure S6A, B; Peters’ test: *t(21)* = 3.38, *p* =.003). Heterogeneity was substantial (total *I^2^ =* 65.59%), primarily attributable to between-study variation (*I^2^_between_* = 63.85%; *τ̂*^2^ = 0.0508; 95% CI: 0.0022-0.6306), Within-study variation was minimal (*I^2^_within_* = 1.74%; *τ̂*^2^ = 0.0014; 95% CI: 0.0000-0.0323). The prediction interval range was OR = 1.10-3.24, suggesting greater certainty in the estimated effect. Meta-regressions were conducted with mean baseline age, follow-up duration, age group, pain informant, pain type, risk of bias, and anxiety/depression operationalisation (Table S13). Associations were found to be weaker when combined anxiety and depression was operationalised as categorical outcomes.

**Figure 7.**
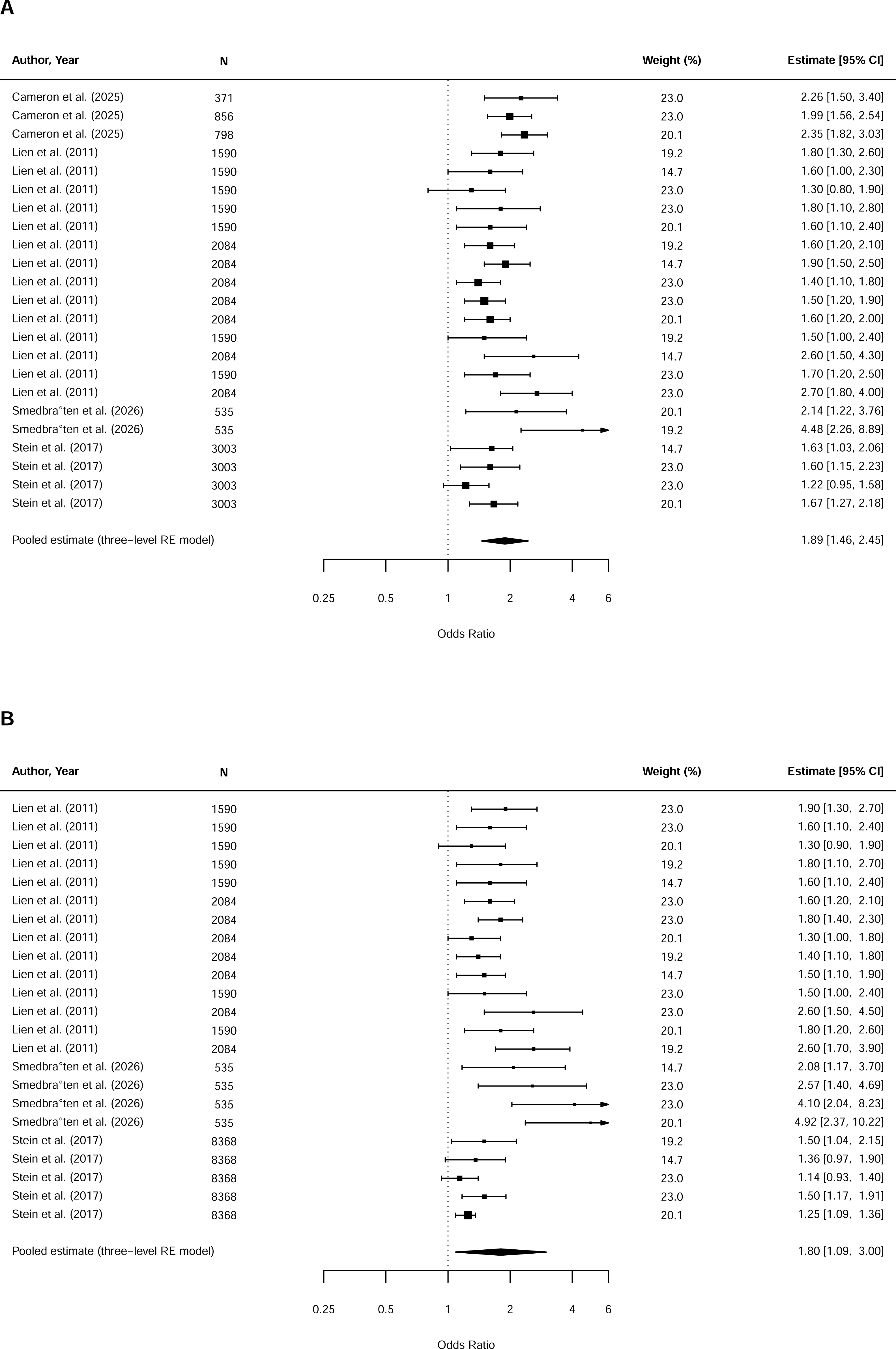
Forest plots of meta-analyses of prospective associations of pain and subsequent combined anxiety/depression. (A) Unadjusted associations; (B) Adjusted associations.

##### 3.4.6.2. Adjusted association

Meta-analysis of 23 associations from three studies with categorical anxiety/depression outcomes [66,108,114] showed that pain was significantly associated with anxiety and depression (*n* = 10,987; *k* = 23; OR = 1.80; 95% CI: 1.09-3.00; *p* =.025; Figure 7B). Significant Peters’ test suggested funnel plot asymmetry (*t(21)* = 5.42; *p* <.001), but the funnel plots indicated it was not due to small-study effects (Figure S6C, D). Heterogeneity was considerable (total *I^2^ =* 87.72%) and mainly driven by between-study variation (*I^2^_between_* = 87.41%; *τ̂*^2^ = 0.1703; 95% CI: 0.0241-3.2844). Within-study heterogeneity variance was small (*I^2^_within_* = 0.31%; *τ̂*^2^= 0.0006; 95% CI: 0.0000-0.0326). The prediction interval crossed the null value (OR = 0.67-4.89), indicating less certainty in the estimated effect. Meta-regressions with mean baseline age, age group, follow-up duration, pain informant, pain operationalisation, pain type, and risk of bias showed that longer follow-up durations were associated with weaker associations (Table S14).

### 3.5. Narrative synthesis of effects not included in meta-analysis

#### 3.5.1. Anxiety and subsequent pain

Associations that could not be meta-analysed were all adjusted associations reported in four studies with continuous pain outcomes [4,6,122,129] and five with categorical outcomes [31,84,87,98,135]. For Yang et al. (2025), the associations that could not be meta-analysed were part of the bidirectional associations between anxiety with functional abdominal pain disorders (FAPD). Results were inconsistent. Among the former four, two studies [4,6] found some significant positive associations between anxiety and subsequent pain symptoms, adjusting for baseline pain, age, and sex, although significance depended on measurement timepoints. The other two [122,129] found no significant association. Among studies with categorical outcomes, three reported no significant association between anxiety and subsequent pain [31,84,87] after adjustment for confounders. One study [98] found a significant positive association for headache, but not abdominal or MSK pain. The other [135] found significant positive associations between anxiety and subsequent FAPD, adjusting for concurrent pain.

#### 3.5.2. Depression and subsequent pain

Regarding unadjusted associations, one study [62] reported significantly higher mean depressive symptom scores in the frequent pain, compared to infrequent pain groups, but the differences occurred to abdominal pain and MSK pain, but not headache. Another study [109] conducted two sets of unadjusted analyses, one with withdrawn/depressed behaviours measured with the Youth Self-Report [1], and one with depression symptoms measured with the Beck Depression Inventory for Youth [8]. The latter was not included in the meta-analysis and showed no significant differences in mean scores among adolescents with no low back pain, those with pain with minimal impact, and those with pain with impact.

For adjusted associations, results were inconsistent. One study with continuous pain [7] reported a significant positive association after adjusting for age, baseline BMI, gynaecological age, contraceptive use, and average duration of menstruation, but the association became non-significant after adjusting additionally for baseline menstrual symptoms and/or somatic complaints. Among five studies with categorical pain [68,84,98,109,135], two studies [98,135] found significant associations between depression and subsequent pain, after adjustment for confounders. Meanwhile, two found no significant associations [84,109]. One [68] found depression was associated with higher risk for incident chronic pain, but not incident total chronic daily headache or tension-type headache.

#### 3.5.3. Combined anxiety and depression and subsequent pain

Results of unadjusted associations were inconsistent. One study found no significant association [66], whilst two others [62,113] found significant positive ones between anxiety and depression with pain, yet the strengths differed by pain type (headache, abdominal pain, or MSK pain) and anxiety/depression informant (self-reported, parent-reported, or teacher-reported).

Findings from adjusted analyses were mixed. Among studies with continuous pain, two studies [79,103] reported some significant associations between anxiety/depression and subsequent pain, whereas one study [62] found none. One study [13] found that anxiety and depression significantly mediated the associations between sleep problems and subsequent severity of abdominal pain, but not headache or MSK pain. One study [113] found inconsistent associations that varied by informant, pain outcomes, age at follow-up, and sex. One study with categorical pain [109] found no significant association between anxiety/depression and having low back pain at follow-up.

#### 3.5.4. Pain and subsequent anxiety

One study [58] reported no significant unadjusted association between headache or abdominal pain with anxiety, but significant ones for MSK pain. For adjusted associations, three studies with continuous anxiety [6,129,135] generally found a significant positive association after adjustment for confounders. However, one study [6] reported that significance depended on age, being significant between pain at age 12 and anxiety at age 13, but not between pain at age 13 and anxiety at age 14.

#### 3.5.5. Pain and subsequent depression

One study [58] reported significant unadjusted associations between MSK pain and depression, but not for headache or abdominal pain. For adjusted associations, one study [109] reported significant associations between FAPD and depression symptoms at three months later.

#### 3.5.6. Pain and subsequent anxiety and depression combined

Two studies with continuous anxiety/depression outcomes [61,130] found no significant association between pain and subsequent anxiety/depression symptoms in children, after adjusting for confounders such as sex, age, ethnicity, parental psychological distress, and familial environment.

#### 3.5.7. Bidirectionality between pain and anxiety/depression

Five studies from five cohorts examined bidirectional associations between pain and anxiety/depression in young people [4,6,129,134,135]. Three included three or more waves [4,6,135], whereas two included two waves [129,134]. Among four studies [4,6,129,135] examined pain and anxiety, but findings were mixed. One [135] reported consistent bidirectional associations across three waves. Two studies [4,6] also reported significant bidirectional associations, but they were not consistent across all ages. One study [4] found consistent associations between MSK pain and anxious mood across ages 14-18. Meanwhile, the other [6] found evidence of a cascading pattern, whereby pain at age 12 predicted anxiety at age 13, which in turn predicted pain at age 14. In contrast, one study [129] found only unidirectional association from pain to anxiety. For pain and depression, both studies [4,134] found bidirectional associations between pain and depression across all waves.

### 3.6. Risk of bias

Overall, associations in 13 studies were rated as low risk of bias; 7 as having some concerns; 19 as high risk; and 3 as very high risk of bias. Detailed risk-of-bias plots for each association, generated using the robvis tool [75], are provided in the Supplementary Materials. The most common reasons for higher risk of bias were inadequate adjustment for important confounders, adjustment for potential mediators, and insufficient handling of missing data. Studies using parental reports of pain were also judged as having some risk of bias because of potential lack of reliable agreement between parent-proxy and child ratings of pain [30].

### 3.7. GRADE

GRADE findings are provided in Table S15. Two out of 14 meta-analyses had high quality of evidence, including the adjusted associations between depression with subsequent continuous pain and between pain with subsequent continuous depression. The other 12 meta-analyses were judged to be of very low to moderate quality, meaning further research may impact on our confidence in the estimated effects. We downgraded most outcomes due to serious/very serious risk of bias, inconsistency resulting from considerable between-study heterogeneity, and imprecision.

## 4. Discussion

This review aimed to examine the direction and strength of the temporal associations between pain and anxiety/depression among young people aged 0-24, using longitudinal, prospective, population-and community-based studies. Meta-analyses demonstrated significant positive associations between anxiety/depression and subsequent pain. After adjustment for confounders, associations were attenuated but remained significant for depression. Pain was also positively associated with subsequent anxiety/depression, and these associations remained significant upon confounder adjustment. There was no evidence of publication bias. Risk of bias significantly moderated the adjusted association between combined anxiety/depression and subsequent pain; however, findings from studies with low risk of bias were consistent with the overall meta-analysis, supporting the robustness of the primary findings. Risk of bias did not significantly moderate effect sizes in the remaining meta-analyses. Nevertheless, study quality varied considerably across the included studies, and the overall certainty of evidence was mostly moderate to very low. These findings should thus be interpreted with caution. In addition to unidirectional associations, we also found evidence for bidirectional associations between pain and anxiety/depression during adolescence. For studies that could not be quantitatively synthesised, narrative evidence generally showed mixed findings.

Our results on the association between anxiety/depression with subsequent pain are consistent with previous meta-analyses showing that emotional problems like anxiety and depression predicted post-operative pain [19], recurrent headache [50], and musculoskeletal pain [51] in children and adolescents. We extended these findings by demonstrating the associations across a broader range of pain types, including abdominal and menstrual pain, and across not only childhood and adolescence but also young adulthood. In contrast, our results diverged from two smaller reviews [45,54], which showed no significant associations between mental distress/depression and subsequent pain. Nevertheless, each was based on a single study, one of which assessed general psychological difficulties rather than anxiety/depression [45], whilst the other included participants beyond our eligible age range [54].

The findings of diminishing associations for anxiety and combined anxiety/depression after confounder adjustment were in line with two previous systematic reviews of emotional problems and recurrent headache [50] or abdominal pain [49], but not with the review of musculoskeletal pain [51]. In contrast, our result of a remaining significant association for depression was consistent with the latter review. Although these discrepancies may reflect differences between pain conditions, pain types did not significantly moderate associations in our analyses. However, the small number of effects included in the meta-regressions may have limited statistical power to detect true differences by pain types. Alternatively, the discrepancies could stem from different studies and numbers of effect sizes included across reviews.

The results that pain was associated with subsequent anxiety/depression aligned with previous reviews showing elevated rates of anxiety and depression in young people with pain as compared to those without [23,27]. They also aligned with findings from clinical populations [12]. We added to the literature by showing that pain may also increase the risk for elevated anxiety and depression symptoms or disorders, and this risk exists above the effect of other factors such as age or sex. Furthermore, whilst previous studies focused on clinical pain, we showed that such risk may exist even when pain is subclinical.

The diminishing associations between anxiety and combined anxiety/depression with subsequent pain after confounder adjustment suggest that these relationships may be partly explained by shared antecedents. Existing models such as shared vulnerability model [71] and mutual maintenance model [111] have proposed that the co-occurrence of pain and anxiety/depression is driven by overlapping psychological (e.g., anxiety sensitivity, uncertainty tolerance, avoidance, emotional dysregulation, sleep disturbances), familial (e.g., parental pain, mental health, parenting styles), and environmental (e.g., stress exposure) factors. Empirical studies have also supported these models, showing that factors such as sleep disturbances [41,81,95], early life adversity [21,101], and parental history of pain and/or mental health problems [17,72,98] are linked to the development of both pain and anxiety/depression in young people, and therefore, may be their shared antecedents. In addition, they have also demonstrated the role of shared genetic liabilities [73,102].

We note that upon adjustment for confounders, depression remained significantly associated with subsequent pain, suggesting that, unlike anxiety, shared antecedents may not fully account for this relationship. Instead, depression may also contribute to later pain, perhaps via additional neurobiological and emotional regulation pathways. For example, a recent neuroimaging study found altered activity and functional connectivity in several brain regions, including the amygdala, middle frontal gyrus, insular lobe, temporoparietal, and occipital lobes among depressed adolescents with co-occurring pain, compared with their pain-free counterparts [120]. In addition, suicidality, a potential symptom of severe depression, has also been shown to predict persistent pain in young people [46]. It has also been shown that elevated depression symptoms contribute to later pain through increased dysregulation, especially greater difficulties accessing effective emotion regulation strategies [135].

The aforementioned models and mechanisms may also explain the associations between pain and subsequent anxiety/depression. In addition, neurobiological factors, such as low levels of serotonin and brain-derived neurotrophic factor, increased inflammation, and altered brain connectivity in the limbic regions have been found to be associated with these problems and theorised to underlie the development of mental health, including anxiety/depression, in youths with chronic pain [125].

Importantly, many studies did not report sufficient data to be meta-analysed and therefore could only be synthesised narratively. This likely reduced the statistical power of the three-level meta-analyses, particularly of the adjusted associations between anxiety/depression and subsequent pain, and between pain and subsequent continuous depression. The smaller number of studies and effect sizes included also limited the ability to reliably detect potential publication bias due to small-study effects [115]. In addition, most meta-analyses of adjusted associations with continuous outcomes could not be conducted, therefore, findings were limited to categorical pain and/or anxiety/depression and may not have captured the full nature of these relationships. Further, unadjusted and adjusted associations were not always based on the same studies. Even when the same studies were included, they often differed in sample size. For these reasons, our findings must be interpreted with caution.

The meta-regressions showed that the unadjusted associations between anxiety/depression and subsequent pain, and between pain and subsequent anxiety tend to be stronger in females than males. This is consistent with evidence that females are at greater risk for both pain [18,50] and anxiety/depression [3,69], and may therefore also be more vulnerable to the co-occurrence. Potential explanations include higher anxiety sensitivity [9], lower pain threshold and tolerance [55], and higher susceptibility to inflammation [60] in women. In addition, higher testosterone levels in men appears protective against pain and anxiety/depression [55,76,110]. However, sex moderation could not be tested on all associations, particularly adjusted associations. Furthermore, a few meta-regressions were based on a small number of studies, potentially limiting power to detect true sex differences.

Age also moderated the effect of pain on subsequent anxiety and depression, with stronger associations observed in studies with older participants at baseline. Nevertheless, these analyses were conducted exclusively with adolescent studies. We could compare between children and adolescents in a subset of meta-regressions and found significant age-group moderation effect only for the adjusted association between pain and subsequent anxiety. Notably, for most age-group meta-regressions, the number of effects for adolescents were substantially higher than those for children. For example, the significant meta-regression included 24 effects for adolescents but only two for children. Such small and unequal subgroup sizes may have reduced statistical power and the reliability of their estimates. Therefore, evidence for age group differences remains tentative.

Previous research has shown that pain, anxiety/depression, and their co-occurrences increase in adolescence [56,82,117]. Puberty may be a key driver, as changes in sex hormones, physical maturation, and brain structure/functions during this period have been implicated in the development of these problems [82,91,126]. Consequently, studies with older adolescents at baseline may have captured more participants further along in puberty. Notably, sex differences in pain and anxiety/depression have also been shown to emerge with puberty [43,110], though we were unable to test an interaction between age and sex due to insufficient data. Future studies should clarify how the relationships between pain and anxiety/depression may shift across age and sex.

This review provides the first and most comprehensive synthesis to date of longitudinal, prospective, population and community-based studies examining the direction and strength of the temporal associations between pain and anxiety/depression in children, adolescents, and young adults. Restricting inclusion to longitudinal, prospective studies strengthened inferences about temporality and reduced recall bias, whilst focusing on population-and community-based samples improved generalisability beyond clinical settings. This review is also the first to synthesise evidence on bidirectional associations between pain and anxiety/depression in young people, offering preliminary evidence of a reciprocal relationship. We further included a broad range of pain and anxiety/depression symptoms and conditions, providing a more comprehensive understanding of these relationships over time. Methodologically, we used three-level meta-analysis to account for correlated effect sizes and improve statistical power. We also used ROBINS-E, which is tailored to epidemiologically observational studies, ensuring rigorous evaluation of study quality.

Despite these strengths, this review has several limitations. First, there was considerable heterogeneity across the meta-analyses, mainly at the between-study level. Several key variables, such as sex, mean baseline age, follow-up duration, pain type, pain informant, pain and anxiety/depression operationalisation, and risk of bias partly explained heterogeneity in some meta-analyses, though for others, no significant moderators were found. We note that the number of effect sizes included in some meta-regressions were relatively small and imbalance among subgroups, limiting their statistical power. In addition, because of the limited information in the original studies, we were unable to examine all moderators. Secondly, pain and anxiety/depression’s definitions and measurements varied considerably across studies, making it difficult to synthesise and determine how associations differ across their phenotypes. Due to limited information in the original studies, we could only conduct meta-regressions using broad categories (e.g., continuous versus categorical measures), which could not capture differences across more nuanced pain and anxiety/depression phenotypes. Finally, we excluded non-English articles, so may have missed relevant studies published in other languages and potentially introduced language bias. Nevertheless, we conducted an extensive search across multiple databases and included both published and unpublished literature. Doing this helped maximise the completeness of our review and minimise publication bias.

This review highlights several important gaps and directions for future research. First, additional high-quality longitudinal studies are needed to increase confidence in the available evidence, especially studies with young adults due to limited existing evidence. New studies should ensure sufficient adjustment for important confounders and utilise appropriate data missingness mechanisms to reduce risk of bias.

Second, more studies are needed to elucidate bidirectional associations between anxiety/depression and pain in young people. Whilst we have found some evidence, it came from only five studies, of which only three included three or more waves of data, limiting our understanding of reciprocal relationships. Further, all studies examined anxiety and depression separately, without adjusting for the comorbid mental health symptom, despite evidence that anxiety and depression are strongly associated in young people [28]. This makes it difficult to distinguish whether observed effects are direct or operate indirectly through the other mental health condition, limiting our understanding of the specific mechanisms linking pain and anxiety/depression over time. Moreover, all five studies were conducted with adolescents, leaving bidirectional associations in children and young adults and how they may develop across childhood, adolescence, young adulthood largely unexplored. Future research should examine bidirectional associations across developmental stages whilst accounting for the correlation between anxiety and depression.

Third, future studies should improve reporting of associations within moderator subgroups, allowing future synthesis to explain observed heterogeneity better. In addition, other unexamined variables that may have explained observed heterogeneity, such as race and ethnicity, household socioeconomic status, parenting environment, and pain chronicity [51,111,125] could not be examined because of insufficient information. Therefore, they should be explored in future research.

Fourth, many studies used pain measures that had not been validated in population-and/or community-based samples of young people. Given that pain is a multifaceted experience [128], future studies should use validated measures and adopt more refined approaches to pain phenotyping. Suggestions include harmonising related pain phenotypes or integrating data collected through various sources to characterise pain more fully [65]. More broadly, the development of standardised frameworks for classifying pain phenotypes in young people, similar to the pyramid model of phenotyping chronic neuropathic pain proposed by Liang and colleagues [65], would facilitate more meaningful comparisons across studies and improve future evidence synthesis.

Finally, most included studies were conducted in high-income countries in Europe, North America, and Australia, with few studies in other regions. As culture can influence the experience and expression of pain and anxiety/depression [59,88], and the prevalence and risk of these conditions vary across countries/regions [14,29,136], their longitudinal associations may also differ across cultural contexts. Future studies should examine country/region as a potential moderator of these associations. They should also consider how lower-order factors such as socioeconomic status [93,99] and immigration status [14,100] can affect pain and anxiety/depression within each country. Such work may help inform more culturally appropriate interventions for young people.

Our findings have important implications for clinical practice and public health. Clinically, mental health screening has been recommended among young people with pain [133]. Our findings suggest it is also important for clinicians to consider associated pain symptoms among youths with elevated anxiety/depression. Importantly, a recent study found that children and adolescents took longer to receive a pain diagnosis after a mental disorder diagnosis, than vice versa [12], suggesting potential clinical oversight. Although this study was conducted within the Swedish healthcare system, similar oversight may occur elsewhere. To reduce this and improve the quality of patient management, healthcare professionals across disciplines, therefore, need to be informed of the potential associations between pain and anxiety/depression, and to recognise young people’s vulnerability to pain when they present with mental health difficulties, and vice versa.

Our review also highlights the need for treatments targeting both pain and anxiety/depression in young people. It has been noted that many psychological treatments for pain focus only on changing pain-related cognitive processes, without addressing elevated anxiety and depression [125], demonstrating a limitation in existing treatments. Transdiagnostic treatment that targets emotional processes underlying both pain and anxiety/depression has shown preliminary positive results [2], but larger controlled trials are needed to confirm its overall effectiveness.

On a public health level, our findings demonstrate the need to improve public understanding of the relationship between pain and anxiety/depression in young people. It is especially important for parents and educators to be aware of the associations so that they can observe early signs of comorbidity and seek timely support for the young individuals. This may matter most when symptoms do not yet reach clinical thresholds. It has been found that persistent high levels of subclinical anxiety and depression symptoms in childhood and adolescence increased the risk of psychiatric disorders in adulthood [80]. In addition, unmanaged or undertreated acute pain can transition into chronic pain, increasing physical complications, slowing wound healing, heightening distress, and raising healthcare costs [52]. Research that informs early prevention efforts and intervention is therefore essential.

## Conflict of interest statement

The authors have no conflicts of interest to declare.

## Data Availability

All data produced in the present study are available upon reasonable request to the authors

## Acknowledgement

The authors want to thank Tomasz Kowalczyk (University of Sussex’s Library) for his advice and guidance on developing search strategy. This research was supported by the Psychology Doctoral Studentship from the School of Psychology, Faculty of Science, Engineering and Medicine, University of Sussex, awarded to Thi Viet Hai Tran.

## Notes

### Competing Interest Statement

The authors have declared no competing interest.

