## Supplementary Material for "Temporal associations between pain and anxiety/depression in young people: A systematic review and meta-analysis of longitudinal, prospective, population- and community-based studies"

### 1. Search strategies

**PsycInfo**

1. tiab(pain OR pains OR painful OR somatoform OR somatic OR headache OR "stomachache" OR "stomach ache" OR backache OR "back ache" OR "toothache" OR "tooth ache" OR "ear ache" OR "earache" OR "muscle ache" OR allodynia OR neuralgia OR myalgia OR fibromyalgia OR arthralgia OR odontalgia OR coxalgia OR cephalalgia OR otalgia OR lumbalgia OR gastralgia OR causalgia OR vulvodynia OR mastodynia OR mastalgia OR arthrodynia OR polyarthralgia OR migraine OR angina OR zoster OR "irritable bowel" OR ulcer OR arthritis OR musculoskeletal OR temporomandibular OR "TMJ disorder*" OR dysmenorrhea OR endometriosis OR arthritis) OR summary(pain OR pains OR painful OR somatoform OR somatic OR headache OR "stomachache" OR "stomach ache" OR backache OR "back ache" OR "toothache" OR "tooth ache" OR "ear ache" OR "earache" OR "muscle ache" OR allodynia OR neuralgia OR myalgia OR fibromyalgia OR arthralgia OR odontalgia OR coxalgia OR cephalalgia OR otalgia OR lumbalgia OR gastralgia OR causalgia OR vulvodynia OR mastodynia OR mastalgia OR arthrodynia OR polyarthralgia OR migraine OR angina OR zoster OR "irritable bowel" OR ulcer OR arthritis OR musculoskeletal OR temporomandibular OR "TMJ disorder*" OR dysmenorrhea OR endometriosis OR arthritis) OR subject(pain OR pains OR painful OR somatoform OR somatic OR headache OR "stomachache" OR "stomach ache" OR backache OR "back ache" OR "toothache" OR "tooth ache" OR "ear ache" OR "earache" OR "muscle ache" OR allodynia OR neuralgia OR myalgia OR fibromyalgia OR arthralgia OR odontalgia OR coxalgia OR cephalalgia OR otalgia OR lumbalgia OR gastralgia OR causalgia OR vulvodynia OR mastodynia OR mastalgia OR arthrodynia OR polyarthralgia OR migraine OR angina OR zoster OR "irritable bowel" OR ulcer OR arthritis OR musculoskeletal OR temporomandibular OR "TMJ disorder*" OR dysmenorrhea OR endometriosis OR arthritis)

2. tiab(anxi* OR "separation anxiety" OR "selective mutism" OR phobia OR "sociophobia" OR "sociophobias" OR "social anxiety" OR "panic disorder" OR agoraphobia OR "generalized anxiety disorder" OR "generalised anxiety disorder" OR "anxiety disorder" OR "anxious distress" OR "overanxious") OR summary(anxi* OR "separation anxiety" OR "selective mutism" OR phobia OR "sociophobia" OR "sociophobias" OR "social anxiety" OR "panic disorder" OR agoraphobia OR "generalized anxiety disorder" OR "generalised anxiety disorder" OR "anxiety disorder" OR "anxious distress" OR "overanxious") OR subject(anxi* OR "separation anxiety" OR "selective mutism" OR phobia OR "sociophobia" OR "sociophobias" OR "social anxiety" OR "panic disorder" OR agoraphobia OR "generalized anxiety disorder" OR "generalised anxiety disorder" OR "anxiety disorder" OR "anxious distress" OR "overanxious")
3. tiab(depression OR depress* OR "disruptive mood dysregulation disorder" OR "major depressive disorder" OR MDD OR "persistent depressive disorder" OR PDD OR "premenstrual dysphoric disorder" OR PMDD OR "depressive disorder" OR dysthymia OR dysthymic OR "dysthymic disorder" OR melancholia OR melancholic OR "melancholic depression") OR summary(depression OR depress* OR "disruptive mood dysregulation disorder" OR "major depressive disorder" OR MDD OR "persistent depressive disorder" OR PDD OR "premenstrual dysphoric disorder" OR PMDD OR "depressive disorder" OR dysthymia OR dysthymic OR "dysthymic disorder" OR melancholia OR melancholic OR "melancholic depression") OR subject(depression OR depress* OR "disruptive mood dysregulation disorder" OR "major depressive disorder" OR MDD OR "persistent depressive disorder" OR PDD OR "premenstrual dysphoric disorder" OR PMDD OR "depressive disorder" OR dysthymia OR dysthymic OR "dysthymic disorder" OR melancholia OR melancholic OR "melancholic depression")

4. tiab("mood disorder*" OR "affective disorder*" OR "emotional disorder*" OR internalizing OR internalising OR "psychological distress") OR summary("mood disorder*" OR "affective disorder*" OR "emotional disorder*" OR internalizing OR internalising OR "psychological distress") OR subject("mood disorder*" OR "affective disorder*" OR "emotional disorder*" OR internalizing OR internalising OR "psychological distress")
5. tiab(longitudinal OR longitudinally OR prospective OR prospectively OR cohort OR "follow-up" OR "follow up" OR panel OR "followed" OR "follow" OR "followed-up" OR temporal) OR summary(longitudinal OR longitudinally OR prospective OR prospectively OR cohort OR "follow-up" OR "follow up" OR panel OR "followed" OR "follow" OR "followed-up" OR temporal) OR subject(longitudinal OR longitudinally OR prospective OR prospectively OR cohort OR "follow-up" OR "follow up" OR panel OR "followed" OR "follow" OR "followed-up" OR temporal)
6. tiab("early life" OR adolescence OR adolescent* OR teenager* OR teen* OR youth* OR "young adulthood" OR "young adult*" OR "infant*" OR "toddler" OR "preschooler" OR child*) OR summary("early life" OR adolescence OR adolescent* OR teenager* OR teen* OR youth* OR "young adulthood" OR "young adult*" OR "infant*" OR "toddler" OR "preschooler" OR child*) OR subject("early life" OR adolescence OR adolescent* OR teenager* OR teen* OR youth* OR "young adulthood" OR "young adult*" OR "infant*" OR "toddler" OR "preschooler" OR child*)
7. 2 OR 3 OR 4
8. 1 AND 5 AND 6 AND 7
9. 8 filtering by English

Full final search terms:

((tiab("mood disorder*" OR "affective disorder*" OR "emotional disorder*" OR internalizing OR internalising OR "psychological distress") OR summary("mood disorder*" OR "affective disorder*" OR "emotional disorder*" OR internalizing OR internalising OR "psychological distress") OR subject("mood disorder*" OR "affective disorder*" OR "emotional disorder*" OR internalizing OR internalising OR "psychological distress")) OR (tiab(depression OR depress* OR "disruptive mood dysregulation disorder" OR "major depressive disorder" OR MDD OR "persistent depressive disorder" OR PDD OR "premenstrual dysphoric disorder" OR PMDD OR "depressive disorder" OR dysthymia OR dysthymic OR "dysthymic disorder" OR melancholia OR melancholic OR "melancholic depression") OR summary(depression OR depress* OR "disruptive mood dysregulation disorder" OR "major depressive disorder" OR MDD OR "persistent depressive disorder" OR PDD OR "premenstrual dysphoric disorder" OR PMDD OR "depressive disorder" OR dysthymia OR dysthymic OR "dysthymic disorder" OR melancholia OR melancholic OR "melancholic depression") OR subject(depression OR depress* OR "disruptive mood dysregulation disorder" OR "major depressive disorder" OR MDD OR "persistent depressive disorder" OR PDD OR "premenstrual dysphoric disorder" OR PMDD OR "depressive disorder" OR dysthymia OR dysthymic OR "dysthymic disorder" OR melancholia OR melancholic OR "melancholic depression")) OR (tiab(anxi* OR "separation anxiety" OR "selective mutism" OR phobia OR "sociophobia" OR "sociophobias" OR "social anxiety" OR "panic disorder" OR agoraphobia OR "generalized anxiety disorder" OR "generalised anxiety disorder" OR "anxiety disorder" OR "anxious distress" OR "overanxious") OR summary(anxi* OR "separation anxiety" OR "selective mutism" OR phobia OR "sociophobia" OR "sociophobias" OR "social anxiety" OR "panic disorder" OR agoraphobia OR "generalized anxiety disorder" OR "generalised anxiety disorder" OR "anxiety disorder" OR "anxious distress" OR "overanxious") OR subject(anxi* OR "separation anxiety" OR "selective mutism" OR phobia OR "sociophobia" OR "sociophobias" OR "social anxiety" OR "panic disorder" OR agoraphobia OR "generalized anxiety disorder" OR "generalised anxiety disorder" OR "anxiety disorder" OR "anxious distress" OR "overanxious"))) AND (tiab("early life" OR adolescence OR adolescent* OR teenager* OR teen* OR youth* OR "young adulthood" OR "young adult*" OR "infant*" OR "toddler" OR "preschooler" OR child*) OR summary("early life" OR adolescence OR adolescent* OR teenager* OR teen* OR youth* OR "young adulthood" OR "young adult*" OR "infant*" OR "toddler" OR "preschooler" OR child*) OR subject("early life" OR adolescence OR adolescent* OR teenager* OR teen* OR youth* OR "young adulthood" OR "young adult*" OR "infant*" OR "toddler" OR "preschooler" OR child*)) AND (tiab(longitudinal OR longitudinally OR prospective OR prospectively OR cohort OR "follow-up" OR "follow up" OR panel OR "followed" OR "follow" OR "followed-up" OR temporal) OR summary(longitudinal OR longitudinally OR prospective OR prospectively OR cohort OR "follow-up" OR "follow up" OR panel OR "followed" OR "follow" OR "followed-up" OR temporal) OR subject(longitudinal OR longitudinally OR prospective OR prospectively OR cohort OR "follow-up" OR "follow up" OR panel OR "followed" OR "follow" OR "followed-up" OR temporal)) AND (tiab(pain OR pains OR painful OR somatoform OR somatic OR headache OR "stomachache" OR "stomach ache" OR backache OR "back ache" OR "toothache" OR "tooth ache" OR "ear ache" OR "earache" OR "muscle ache" OR allodynia OR neuralgia OR myalgia OR fibromyalgia OR arthralgia OR odontalgia OR coxalgia OR cephalalgia OR otalgia OR lumbalgia OR gastralgia OR causalgia OR vulvodynia OR mastodynia OR mastalgia OR arthrodynia OR polyarthralgia OR migraine OR angina OR zoster OR "irritable bowel" OR ulcer OR arthritis OR musculoskeletal OR temporomandibular OR "TMJ disorder*" OR dysmenorrhea OR endometriosis OR arthritis) OR summary(pain OR pains OR painful OR somatoform OR somatic OR headache OR "stomachache" OR "stomach ache" OR backache OR "back ache" OR "toothache" OR "tooth ache" OR "ear ache" OR "earache" OR "muscle ache" OR allodynia OR neuralgia OR myalgia OR fibromyalgia OR arthralgia OR odontalgia OR coxalgia OR cephalalgia OR otalgia OR lumbalgia OR gastralgia OR causalgia OR vulvodynia OR mastodynia OR mastalgia OR arthrodynia OR polyarthralgia OR migraine OR angina OR zoster OR "irritable bowel" OR ulcer OR arthritis OR musculoskeletal OR temporomandibular OR "TMJ disorder*" OR dysmenorrhea OR endometriosis OR arthritis) OR subject(pain OR pains OR painful OR somatoform OR somatic OR headache OR "stomachache" OR "stomach ache" OR backache OR "back ache" OR "toothache" OR "tooth ache" OR "ear ache" OR "earache" OR "muscle ache" OR allodynia OR neuralgia OR myalgia OR fibromyalgia OR arthralgia OR odontalgia OR coxalgia OR cephalalgia OR otalgia OR lumbalgia OR gastralgia OR causalgia OR vulvodynia OR mastodynia OR mastalgia OR arthrodynia OR polyarthralgia OR migraine OR angina OR zoster OR "irritable bowel" OR ulcer OR arthritis OR musculoskeletal OR temporomandibular OR "TMJ disorder*" OR dysmenorrhea OR endometriosis OR arthritis)) AND la.exact("ENG")

**Web of Science**

1. TS=(pain OR pains OR painful OR somatoform OR somatic OR headache OR "stomachache" OR "stomach ache" OR backache OR "back ache" OR "toothache" OR "tooth ache" OR "ear ache" OR "earache" OR "muscle ache" OR allodynia OR neuralgia OR myalgia OR fibromyalgia OR arthralgia OR odontalgia OR coxalgia OR cephalalgia OR otalgia OR lumbalgia OR gastralgia OR causalgia OR vulvodynia OR mastodynia OR mastalgia OR arthrodynia OR polyarthralgia OR migraine OR angina OR zoster OR "irritable bowel" OR ulcer OR arthritis OR musculoskeletal OR temporomandibular OR "TMJ disorder*" OR dysmenorrhea OR endometriosis OR arthritis)
2. TS=(anxi* OR "separation anxiety" OR "selective mutism" OR phobia OR "sociophobia" OR "sociophobias" OR "social anxiety" OR "panic disorder" OR agoraphobia OR "generalized anxiety disorder" OR "generalised anxiety disorder" OR "anxiety disorder" OR "anxious distress" OR "overanxious")
3. TS=(depression OR depress* OR "disruptive mood dysregulation disorder" OR "major depressive disorder" OR MDD OR "persistent depressive disorder" OR PDD OR "premenstrual dysphoric disorder" OR PMDD OR "depressive disorder" OR dysthymia OR dysthymic OR "dysthymic disorder" OR melancholia OR melancholic OR "melancholic depression")
4. TS=("mood disorder*" OR "affective disorder*" OR "emotional disorder*" OR internalizing OR internalising OR "psychological distress")
5. #2 OR #3 OR #4

6. TS=(longitudinal OR longitudinally OR prospective OR prospectively OR cohort OR "follow-up" OR "follow up" OR panel OR "followed" OR "follow" OR "followed-up" OR temporal)
7. TS=("early life" OR adolescence OR adolescent* OR teenager* OR teen* OR youth* OR "young adulthood" OR "young adult*" OR "infant*" OR "toddler" OR "preschooler" OR child*)
8. #1 AND #5 AND #6 AND #7
9. Refine #8 results by English language

**Scopus**

1. TITLE-ABS-KEY ( pain OR pains OR painful OR somatoform OR somatic OR headache OR "stomachache" OR "stomach ache" OR backache OR "back ache" OR "toothache" OR "tooth ache" OR "ear ache" OR "earache" OR "muscle ache" OR allodynia OR neuralgia OR myalgia OR fibromyalgia OR arthralgia OR odontalgia OR coxalgia OR cephalalgia OR otalgia OR lumbalgia OR gastralgia OR causalgia OR vulvodynia OR mastodynia OR mastalgia OR arthrodynia OR polyarthralgia OR migraine OR angina OR zoster OR "irritable bowel" OR ulcer OR arthritis OR musculoskeletal OR temporomandibular OR "TMJ disorder*" OR dysmenorrhea OR endometriosis OR arthritis ) AND (TITLE-ABS-KEY ( anxi* OR "separation anxiety" OR "selective mutism" OR phobia OR "sociophobia" OR "sociophobias" OR "social anxiety" OR "panic disorder" OR agoraphobia OR "generalized anxiety disorder" OR "generalised anxiety disorder" OR "anxiety disorder" OR "anxious distress" OR "overanxious" ) OR TITLE-ABS-KEY ( depression OR depress* OR "disruptive mood dysregulation disorder" OR "major depressive disorder" OR mdd OR "persistent depressive disorder" OR pdd OR "premenstrual dysphoric disorder" OR pmdd OR "depressive disorder" OR dysthymia OR dysthymic OR "dysthymic disorder" OR melancholia OR melancholic OR "melancholic depression" ) OR TITLE-ABS-KEY ( "mood disorder*" OR "affective disorder*" OR "emotional disorder*" OR internalizing OR internalising OR "psychological distress" ) ) AND TITLE-ABS-KEY(longitudinal OR longitudinally OR prospective OR prospectively OR cohort OR "follow-up" OR "follow up" OR panel OR "followed" OR "follow" OR "followed-up" OR temporal) AND TITLE-ABS-KEY("early life" OR adolescence OR adolescent* OR teenager* OR teen* OR youth* OR "young adulthood" OR "young adult*" OR "infant*" OR "toddler" OR "preschooler" OR child*)
2. Limit #1 on English results

**PubMed**

1. pain[Title/Abstract] OR pains[Title/Abstract] OR painful[Title/Abstract] OR somatoform[Title/Abstract] OR somatic[Title/Abstract] OR headache[Title/Abstract] OR "stomachache"[Title/Abstract] OR "stomach ache"[Title/Abstract] OR backache[Title/Abstract] OR "back ache"[Title/Abstract] OR "toothache"[Title/Abstract] OR "tooth ache"[Title/Abstract] OR "ear ache"[Title/Abstract] OR "earache"[Title/Abstract] OR "muscle ache"[Title/Abstract] OR allodynia[Title/Abstract] OR neuralgia[Title/Abstract] OR myalgia[Title/Abstract] OR fibromyalgia[Title/Abstract] OR arthralgia[Title/Abstract] OR odontalgia[Title/Abstract] OR coxalgia[Title/Abstract] OR cephalalgia[Title/Abstract] OR otalgia[Title/Abstract] OR lumbalgia[Title/Abstract] OR gastralgia[Title/Abstract] OR causalgia[Title/Abstract] OR vulvodynia[Title/Abstract] OR mastodynia[Title/Abstract] OR mastalgia[Title/Abstract] OR arthrodynia[Title/Abstract] OR polyarthralgia[Title/Abstract] OR migraine[Title/Abstract] OR angina[Title/Abstract] OR zoster[Title/Abstract] OR "irritable bowel"[Title/Abstract] OR ulcer[Title/Abstract] OR arthritis[Title/Abstract] OR musculoskeletal[Title/Abstract] OR temporomandibular[Title/Abstract] OR "TMJ disorder*"[Title/Abstract] OR dysmenorrhea[Title/Abstract] OR endometriosis[Title/Abstract] OR arthritis[Title/Abstract]
2. anxi*[Title/Abstract] OR "separation anxiety"[Title/Abstract] OR "selective mutism"[Title/Abstract] OR phobia[Title/Abstract] OR "sociophobia"[Title/Abstract] OR "social anxiety"[Title/Abstract] OR "panic disorder"[Title/Abstract] OR agoraphobia[Title/Abstract] OR "generalized anxiety disorder"[Title/Abstract] OR "generalised anxiety disorder"[Title/Abstract] OR "anxiety disorder"[Title/Abstract] OR "anxious distress"[Title/Abstract] OR "overanxious"[Title/Abstract]
3. depression[Title/Abstract] OR depress*[Title/Abstract] OR "disruptive mood dysregulation disorder"[Title/Abstract] OR "major depressive disorder"[Title/Abstract] OR MDD[Title/Abstract] OR "persistent depressive disorder"[Title/Abstract] OR PDD[Title/Abstract] OR "premenstrual

dysphoric disorder"[Title/Abstract] OR PMDD[Title/Abstract] OR "depressive disorder"[Title/Abstract] OR dysthymia[Title/Abstract] OR dysthymic[Title/Abstract] OR "dysthymic disorder"[Title/Abstract] OR melancholia[Title/Abstract] OR melancholic[Title/Abstract] OR "melancholic depression"[Title/Abstract]

4. "mood disorder*"[Title/Abstract] OR "affective disorder*"[Title/Abstract] OR "emotional disorder*"[Title/Abstract] OR internalizing[Title/Abstract] OR internalising[Title/Abstract] OR "psychological distress"[Title/Abstract]
5. #2 OR #3 OR #4
6. longitudinal[Title/Abstract] OR longitudinally[Title/Abstract] OR prospective[Title/Abstract] OR prospectively[Title/Abstract] OR cohort[Title/Abstract] OR "follow-up"[Title/Abstract] OR "follow up"[Title/Abstract] OR panel[Title/Abstract] OR "followed"[Title/Abstract] OR "follow"[Title/Abstract] OR "followed-up"[Title/Abstract] OR temporal[Title/Abstract]
7. "early life"[Title/Abstract] OR adolescence[Title/Abstract] OR adolescent*[Title/Abstract] OR teenager*[Title/Abstract] OR teen*[Title/Abstract] OR youth*[Title/Abstract] OR "young adulthood"[Title/Abstract] OR "young adult*"[Title/Abstract] OR "infant*"[Title/Abstract] OR "toddler"[Title/Abstract] OR "preschooler"[Title/Abstract] OR child*[Title/Abstract]
8. #1 AND #5 AND #6 AND #7
9. Filtered #8 by English results

Full search terms:

(((pain[Title/Abstract] OR pains[Title/Abstract] OR painful[Title/Abstract] OR somatoform[Title/Abstract] OR somatic[Title/Abstract] OR headache[Title/Abstract] OR "stomachache"[Title/Abstract] OR "stomach ache"[Title/Abstract] OR backache[Title/Abstract] OR "back ache"[Title/Abstract] OR "toothache"[Title/Abstract] OR "tooth ache"[Title/Abstract] OR "ear ache"[Title/Abstract] OR "earache"[Title/Abstract] OR "muscle ache"[Title/Abstract] OR allodynia[Title/Abstract] OR neuralgia[Title/Abstract] OR myalgia[Title/Abstract] OR fibromyalgia[Title/Abstract] OR arthralgia[Title/Abstract] OR odontalgia[Title/Abstract] OR coxalgia[Title/Abstract] OR cephalalgia[Title/Abstract] OR otalgia[Title/Abstract] OR lumbalgia[Title/Abstract] OR gastralgia[Title/Abstract] OR causalgia[Title/Abstract] OR vulvodynia[Title/Abstract] OR mastodynia[Title/Abstract] OR mastalgia[Title/Abstract] OR arthrodynia[Title/Abstract] OR polyarthralgia[Title/Abstract] OR migraine[Title/Abstract] OR angina[Title/Abstract] OR zoster[Title/Abstract] OR "irritable bowel"[Title/Abstract] OR ulcer[Title/Abstract] OR arthritis[Title/Abstract] OR musculoskeletal[Title/Abstract] OR temporomandibular[Title/Abstract] OR "TMJ disorder*"[Title/Abstract] OR dysmenorrhea[Title/Abstract] OR endometriosis[Title/Abstract] OR arthritis[Title/Abstract]) AND (((anxi*[Title/Abstract] OR "separation anxiety"[Title/Abstract] OR "selective mutism"[Title/Abstract] OR phobia[Title/Abstract] OR "sociophobia"[Title/Abstract] OR "social anxiety"[Title/Abstract] OR "panic disorder"[Title/Abstract] OR agoraphobia[Title/Abstract] OR "generalized anxiety disorder"[Title/Abstract] OR "generalised anxiety disorder"[Title/Abstract] OR "anxiety disorder"[Title/Abstract] OR "anxious distress"[Title/Abstract] OR "overanxious"[Title/Abstract]) OR (depression[Title/Abstract] OR depress*[Title/Abstract] OR "disruptive mood dysregulation disorder"[Title/Abstract] OR "major depressive disorder"[Title/Abstract] OR MDD[Title/Abstract] OR "persistent depressive disorder"[Title/Abstract] OR PDD[Title/Abstract] OR "premenstrual dysphoric disorder"[Title/Abstract] OR PMDD[Title/Abstract] OR "depressive disorder"[Title/Abstract] OR dysthymia[Title/Abstract] OR dysthymic[Title/Abstract] OR "dysthymic disorder"[Title/Abstract] OR melancholia[Title/Abstract] OR melancholic[Title/Abstract] OR "melancholic depression"[Title/Abstract])) OR ("mood disorder*"[Title/Abstract] OR "affective disorder*"[Title/Abstract] OR "emotional disorder*"[Title/Abstract] OR internalizing[Title/Abstract] OR internalising[Title/Abstract] OR "psychological distress"[Title/Abstract]))) AND (longitudinal[Title/Abstract] OR longitudinally[Title/Abstract] OR prospective[Title/Abstract] OR prospectively[Title/Abstract] OR cohort[Title/Abstract] OR "follow-up"[Title/Abstract] OR "follow up"[Title/Abstract] OR panel[Title/Abstract] OR "followed"[Title/Abstract] OR "follow"[Title/Abstract] OR "followed-up"[Title/Abstract] OR temporal[Title/Abstract])) AND ("early life"[Title/Abstract] OR adolescence[Title/Abstract] OR adolescent*[Title/Abstract] OR teenager*[Title/Abstract] OR teen*[Title/Abstract] OR youth*[Title/Abstract] OR "young adulthood"[Title/Abstract] OR "young adult*"[Title/Abstract] OR "infant*"[Title/Abstract] OR "toddler"[Title/Abstract] OR "preschooler"[Title/Abstract] OR child*[Title/Abstract]) Filters: English

**ProQuest Dissertations & Theses Global**

1. title(pain OR pains OR painful OR somatoform OR somatic OR headache OR "stomachache" OR "stomach ache" OR backache OR "back ache" OR "toothache" OR "tooth ache" OR "ear ache" OR "earache" OR "muscle ache" OR allodynia OR neuralgia OR myalgia OR fibromyalgia OR arthralgia OR odontalgia OR coxalgia OR cephalalgia OR otalgia OR lumbalgia OR gastralgia OR causalgia OR vulvodynia OR mastodynia OR mastalgia OR arthrodynia OR polyarthralgia OR migraine OR angina OR zoster OR "irritable bowel" OR ulcer OR arthritis OR musculoskeletal OR temporomandibular OR "TMJ disorder*" OR dysmenorrhea OR endometriosis OR arthritis) OR summary(pain OR pains OR painful OR somatoform OR somatic OR headache OR "stomachache" OR "stomach ache" OR backache OR "back ache" OR "toothache" OR "tooth ache" OR "ear ache" OR "earache" OR "muscle ache" OR allodynia OR neuralgia OR myalgia OR fibromyalgia OR arthralgia OR odontalgia OR coxalgia OR cephalalgia OR otalgia OR lumbalgia OR gastralgia OR causalgia OR vulvodynia OR mastodynia OR mastalgia OR arthrodynia OR polyarthralgia OR migraine OR angina OR zoster OR "irritable bowel" OR ulcer OR arthritis OR musculoskeletal OR temporomandibular OR "TMJ disorder*" OR dysmenorrhea OR endometriosis OR arthritis) OR subject(pain OR

pains OR painful OR somatoform OR somatic OR headache OR "stomachache" OR "stomach ache" OR backache OR "back ache" OR "toothache" OR "tooth ache" OR "ear ache" OR "earache" OR "muscle ache" OR allodynia OR neuralgia OR myalgia OR fibromyalgia OR arthralgia OR odontalgia OR coxalgia OR cephalalgia OR otalgia OR lumbalgia OR gastralgia OR causalgia OR vulvodynia OR mastodynia OR mastalgia OR arthrodynia OR polyarthralgia OR migraine OR angina OR zoster OR "irritable bowel" OR ulcer OR arthritis OR musculoskeletal OR temporomandibular OR "TMJ disorder*" OR dysmenorrhea OR endometriosis OR arthritis)

2. title(anxi* OR "separation anxiety" OR "selective mutism" OR phobia OR "sociophobia" OR "sociophobias" OR "social anxiety" OR "panic disorder" OR agoraphobia OR "generalized anxiety disorder" OR "generalised anxiety disorder" OR "anxiety disorder" OR "anxious distress" OR "overanxious") OR summary(anxi* OR "separation anxiety" OR "selective mutism" OR phobia OR "sociophobia" OR "sociophobias" OR "social anxiety" OR "panic disorder" OR agoraphobia OR "generalized anxiety disorder" OR "generalised anxiety disorder" OR "anxiety disorder" OR "anxious distress" OR "overanxious") OR subject(anxi* OR "separation anxiety" OR "selective mutism" OR phobia OR "sociophobia" OR "sociophobias" OR "social anxiety" OR "panic disorder" OR agoraphobia OR "generalized anxiety disorder" OR "generalised anxiety disorder" OR "anxiety disorder" OR "anxious distress" OR "overanxious")
3. title(depression OR depress* OR "disruptive mood dysregulation disorder" OR "major depressive disorder" OR MDD OR "persistent depressive disorder" OR PDD OR "premenstrual dysphoric disorder" OR PMDD OR "depressive disorder" OR dysthymia OR dysthymic OR "dysthymic disorder" OR melancholia OR melancholic OR "melancholic depression") OR summary(depression OR depress* OR "disruptive mood dysregulation disorder" OR "major depressive disorder" OR MDD OR "persistent depressive disorder" OR PDD OR "premenstrual dysphoric disorder" OR PMDD OR "depressive disorder" OR dysthymia OR dysthymic OR "dysthymic disorder" OR melancholia OR melancholic OR "melancholic depression") OR subject(depression

OR depress* OR "disruptive mood dysregulation disorder" OR "major depressive disorder" OR MDD OR "persistent depressive disorder" OR PDD OR "premenstrual dysphoric disorder" OR PMDD OR "depressive disorder" OR dysthymia OR dysthymic OR "dysthymic disorder" OR melancholia OR melancholic OR "melancholic depression")

4. title("mood disorder*" OR "affective disorder*" OR "emotional disorder*" OR internalizing OR internalising OR "psychological distress") OR summary("mood disorder*" OR "affective disorder*" OR "emotional disorder*" OR internalizing OR internalising OR "psychological distress") OR subject("mood disorder*" OR "affective disorder*" OR "emotional disorder*" OR internalizing OR internalising OR "psychological distress")
5. title(longitudinal OR longitudinally OR prospective OR prospectively OR cohort OR "follow-up" OR "follow up" OR panel OR "followed" OR "follow" OR "followed-up" OR temporal) OR summary(longitudinal OR longitudinally OR prospective OR prospectively OR cohort OR "follow-up" OR "follow up" OR panel OR "followed" OR "follow" OR "followed-up" OR temporal) OR subject(longitudinal OR longitudinally OR prospective OR prospectively OR cohort OR "follow-up" OR "follow up" OR panel OR "followed" OR "follow" OR "followed-up" OR temporal)
6. title("early life" OR adolescence OR adolescent* OR teenager* OR teen* OR youth* OR "young adulthood" OR "young adult*" OR "infant*" OR "toddler" OR "preschooler" OR child*) OR summary("early life" OR adolescence OR adolescent* OR teenager* OR teen* OR youth* OR "young adulthood" OR "young adult*" OR "infant*" OR "toddler" OR "preschooler" OR child*) OR subject("early life" OR adolescence OR adolescent* OR teenager* OR teen* OR youth* OR "young adulthood" OR "young adult*" OR "infant*" OR "toddler" OR "preschooler" OR child*)
7. 2 OR 3 OR 4
8. 1 AND 5 AND 6 AND 7
9. Filtered #8 by English

Full final search terms:

((title("mood disorder*" OR "affective disorder*" OR "emotional disorder*" OR internalizing OR internalising OR "psychological distress") OR summary("mood disorder*" OR "affective disorder*" OR "emotional disorder*" OR internalizing OR internalising OR "psychological distress") OR subject("mood disorder*" OR "affective disorder*" OR "emotional disorder*" OR internalizing OR internalising OR "psychological distress")) OR (title(depression OR depress* OR "disruptive mood dysregulation disorder" OR "major depressive disorder" OR MDD OR "persistent depressive disorder" OR PDD OR "premenstrual dysphoric disorder" OR PMDD OR "depressive disorder" OR dysthymia OR dysthymic OR "dysthymic disorder" OR melancholia OR melancholic OR "melancholic depression") OR summary(depression OR depress* OR "disruptive mood dysregulation disorder" OR "major depressive disorder" OR MDD OR "persistent depressive disorder" OR PDD OR "premenstrual dysphoric disorder" OR PMDD OR "depressive disorder" OR dysthymia OR dysthymic OR "dysthymic disorder" OR melancholia OR melancholic OR "melancholic depression") OR subject(depression OR depress* OR "disruptive mood dysregulation disorder" OR "major depressive disorder" OR MDD OR "persistent depressive disorder" OR PDD OR "premenstrual dysphoric disorder" OR PMDD OR "depressive disorder" OR dysthymia OR dysthymic OR "dysthymic disorder" OR melancholia OR melancholic OR "melancholic depression")) OR (title(anxi* OR "separation anxiety" OR "selective mutism" OR phobia OR "sociophobia" OR "sociophobias" OR "social anxiety" OR "panic disorder" OR agoraphobia OR "generalized anxiety disorder" OR "generalised anxiety disorder" OR "anxiety disorder" OR "anxious distress" OR "overanxious") OR summary(anxi* OR "separation anxiety" OR "selective mutism" OR phobia OR "sociophobia" OR "sociophobias" OR "social anxiety" OR "panic disorder" OR agoraphobia OR "generalized anxiety disorder" OR "generalised anxiety disorder" OR "anxiety disorder" OR "anxious distress" OR "overanxious") OR subject(anxi* OR "separation anxiety" OR "selective mutism" OR phobia OR "sociophobia" OR "sociophobias" OR "social anxiety" OR "panic disorder" OR agoraphobia OR "generalized anxiety disorder" OR "generalised anxiety disorder" OR "anxiety disorder" OR "anxious distress" OR "overanxious"))) AND (title(pain OR pains OR painful OR somatoform OR somatic OR headache OR "stomachache" OR "stomach ache" OR backache OR "back ache" OR "toothache" OR "tooth ache" OR "ear ache" OR "earache" OR "muscle ache" OR allodynia OR neuralgia OR myalgia OR fibromyalgia OR arthralgia OR odontalgia OR coxalgia OR cephalalgia OR otalgia OR lumbalgia OR gastralgia OR causalgia OR vulvodynia OR mastodynia OR mastalgia OR arthrodynia OR polyarthralgia OR migraine OR angina OR zoster OR "irritable bowel" OR ulcer OR arthritis OR musculoskeletal OR temporomandibular OR "TMJ disorder*" OR dysmenorrhea OR endometriosis OR arthritis) OR summary(pain OR pains OR painful OR somatoform OR somatic OR headache OR "stomachache" OR "stomach ache" OR backache OR "back ache" OR "toothache" OR "tooth ache" OR "ear ache" OR "earache" OR "muscle ache" OR allodynia OR neuralgia OR myalgia OR fibromyalgia OR arthralgia OR odontalgia OR coxalgia OR cephalalgia OR otalgia OR lumbalgia OR gastralgia OR causalgia OR vulvodynia OR mastodynia OR mastalgia OR arthrodynia OR polyarthralgia OR migraine OR angina OR zoster OR "irritable bowel" OR ulcer OR arthritis OR musculoskeletal OR temporomandibular OR "TMJ disorder*" OR dysmenorrhea OR endometriosis OR arthritis) OR subject(pain OR pains OR painful OR somatoform OR somatic OR headache OR "stomachache" OR "stomach ache" OR backache OR "back ache" OR "toothache" OR "tooth ache" OR "ear ache" OR "earache" OR "muscle ache" OR allodynia OR neuralgia OR myalgia OR fibromyalgia OR arthralgia OR odontalgia OR coxalgia OR cephalalgia OR otalgia OR lumbalgia OR gastralgia OR causalgia OR vulvodynia OR mastodynia OR mastalgia OR arthrodynia OR polyarthralgia OR migraine OR angina OR zoster OR "irritable bowel" OR ulcer OR arthritis OR musculoskeletal OR temporomandibular OR "TMJ disorder*" OR dysmenorrhea OR endometriosis OR arthritis)) AND (title("early life" OR adolescence OR adolescent* OR teenager* OR teen* OR youth* OR "young adulthood" OR "young adult*" OR "infant*" OR "toddler" OR "preschooler" OR child*) OR summary("early life" OR adolescence OR adolescent* OR teenager* OR teen* OR youth* OR "young adulthood" OR "young adult*" OR "infant*" OR "toddler" OR "preschooler" OR child*) OR subject("early life" OR adolescence OR adolescent* OR teenager* OR teen* OR youth* OR "young adulthood" OR "young adult*" OR "infant*" OR "toddler" OR "preschooler" OR child*)) AND (title(longitudinal OR longitudinally OR prospective OR prospectively OR cohort OR "follow-up" OR "follow up" OR panel OR "followed" OR "follow" OR "followed-up" OR temporal) OR summary(longitudinal OR longitudinally OR prospective OR prospectively OR cohort OR "follow-up" OR "follow up" OR panel OR "followed" OR "follow" OR "followed-up" OR temporal) OR subject(longitudinal OR longitudinally OR prospective OR prospectively OR cohort OR "follow-up" OR "follow up" OR panel OR "followed" OR "follow" OR "followed-up" OR temporal)) AND la.exact("ENG")

**Google Scholar**

(pain OR pains OR painful OR somatoform OR somatic OR headache OR "stomachache" OR "stomach ache" OR backache OR "back ache" OR "toothache" OR "tooth ache" OR "ear ache" OR "earache" OR "muscle ache" OR allodynia OR neuralgia OR myalgia OR fibromyalgia OR arthralgia OR odontalgia OR coxalgia OR cephalalgia OR otalgia OR lumbalgia OR gastralgia OR causalgia OR vulvodynia OR mastodynia OR mastalgia OR arthrodynia OR polyarthralgia OR migraine OR angina OR zoster OR "irritable bowel" OR ulcer OR arthritis OR musculoskeletal OR temporomandibular OR "TMJ disorder" OR "TMJ disorders" OR dysmenorrhea OR endometriosis OR arthritis) AND ((anxiety OR anxious OR "separation anxiety" OR "selective mutism" OR phobia OR phobias OR "sociophobia" OR "social anxiety" OR "panic disorder" OR agoraphobia OR "generalized anxiety disorder" OR "generalised anxiety disorder" OR "anxiety disorder" OR "anxious distress" OR "overanxious") OR (depression OR depressive OR depressed OR "disruptive mood dysregulation disorder" OR "major depressive disorder" OR MDD OR "persistent depressive disorder" OR PDD OR "premenstrual dysphoric disorder" OR PMDD OR "depressive disorder" OR dysthymia OR dysthymic OR "dysthymic disorder" OR melancholia OR melancholic OR "melancholic depression") OR ("mood disorder" OR "mood disorders" OR "affective disorder" OR "affective disorders" OR "emotional disorder" OR "emotional disorders" OR internalizing OR internalising OR "psychological distress")) AND (longitudinal OR longitudinally OR prospective OR prospectively OR cohort OR "follow-up" OR "follow up" OR panel OR "followed" OR "follow" OR "followed-up" OR temporal) AND("early life" OR adolescence OR adolescent OR adolescents OR teenager OR teenagers OR teen OR teens OR youth OR youths OR "young adulthood" OR "young adult" OR "young adults" OR "infant" OR "infants" OR "toddler" OR "preschooler" OR child OR children)

Sort by relevance

**OSF Preprints**

Each search syntax was performed individually. Their results were aggregated together.

1. "pain" "anxiety" longitudinal
2. "pain" "depression" longitudinal
3. "pain" "psychological distress" longitudinal
4. "pain" "anxiety" temporal
5. "pain" "depression" temporal
6. "pain" "psychological distress" temporal
7. "pain" "anxiety" follow
8. "pain" "anxiety" followed
9. "pain" "depression" follow
10. "pain" "depression" followed
11. "pain" "psychological distress" follow
12. "pain" "psychological distress" followed
13. "pain" "anxiety" panel
14. "pain" "depression" panel

15. "pain" "psychological distress" panel
16. "pain" "anxiety" prospective
17. "pain" "depression" prospective
18. "pain" "psychological distress" prospective
19. "pain" "internalizing" temporal
20. "pain" "internalising" temporal
21. "pain" "internalizing" "longitudinal"
22. "pain" "internalising" "longitudinal"
23. "pain" "internalizing" follow
24. "pain" "internalizing" followed
25. "pain" "internalizing" panel
26. "pain" "internalising" follow
27. "pain" "internalising" followed
28. "pain" "internalising" panel
29. "pain" "internalizing" prospective
30. "pain" "internalising" prospective

### 2. Data extraction

The data extracted included:

(1) study characteristics (author, year, publication type, funding source, conflict of interest, open access data).

(2) study design (cohort name, sampling method, sampling location, study timeline, recruitment and sampling setting, data collection process, sampling selection, sample size).

(3) participant demographics (i.e., age, gender, socio-economic status, ethnicity, country).

(4) the relationship being studied, including the exposure(s) and outcome(s).

(5) measures of pain, anxiety, and depression (i.e., measurement type, informants, time lag, treatment of variable, cut-off points, psychometric information).

(6) data analysis (confounders, analytical approach, missing data treatment).

(7) key results, including univariable and multivariable associations (descriptive results and/or effect sizes and 95% confidence intervals showing the strength and direction of the associations). When effect sizes and CIs were not provided, if possible, they were calculated from the reported raw data.

### 3. Supplementary figure legends

**Figure S1. Funnel plots of meta-analyses of prospective associations of anxiety and subsequent pain. (A) Funnel plot of unadjusted associations; (B) Contour-enhanced funnel plot of unadjusted associations; (C) Funnel plot of adjusted associations; (D) Contour-enhanced funnel plot of adjusted associations.**

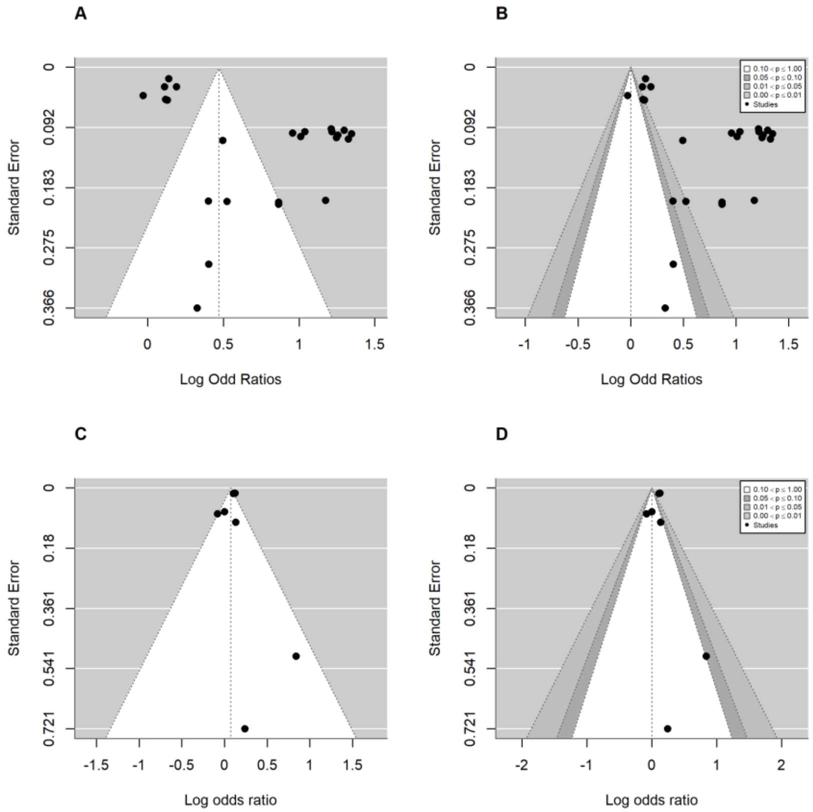

**Figure S2. Funnel plots of meta-analyses of prospective associations of depression and subsequent pain. (A) Funnel plot of unadjusted associations; (B) Contour-enhanced funnel plot of unadjusted associations; (C) Funnel plot of adjusted associations of continuous pain outcomes; (D) Contour-enhanced funnel plot of adjusted associations of continuous pain outcomes; (E) Funnel plot of adjusted associations of categorical pain outcomes; (F) Contour-enhanced funnel plot of adjusted associations of categorical pain outcomes.**

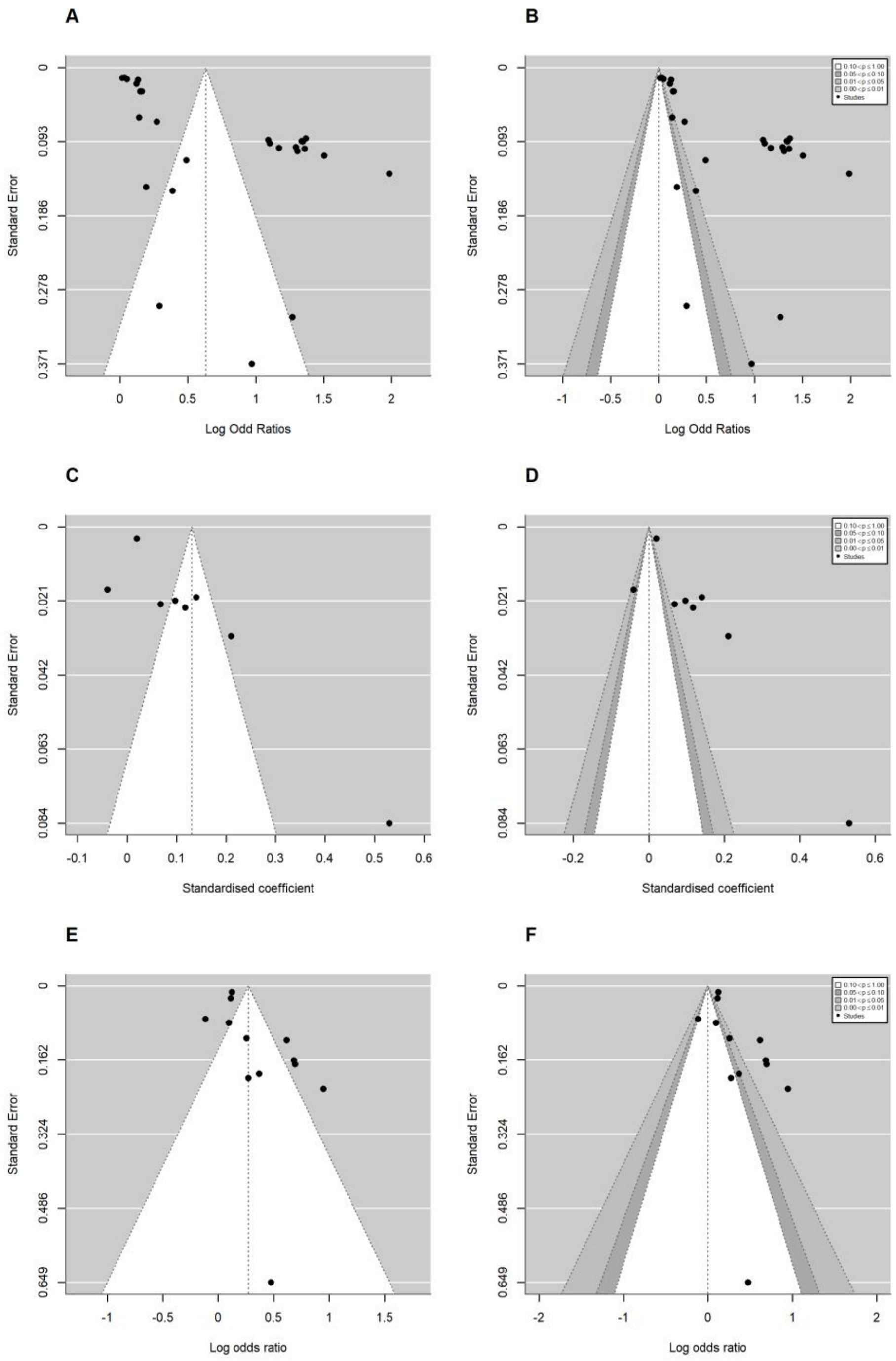
A
B
C
D
E
F
Standard Error
Log Odd Ratios
Standardised coefficient
Log odds ratio
0
0.093
0.186
0.278
0.371
0.021
0.042
0.063
0.084
0.162
0.324
0.486
0.649
Studies

**Figure S3. Funnel plots of meta-analyses of prospective associations of combined anxiety/depression and subsequent pain. (A) Funnel plot of unadjusted associations; (B) Contour-enhanced funnel plot of unadjusted associations; (C) Funnel plot of adjusted associations; (D) Contour-enhanced funnel plot of adjusted associations.**

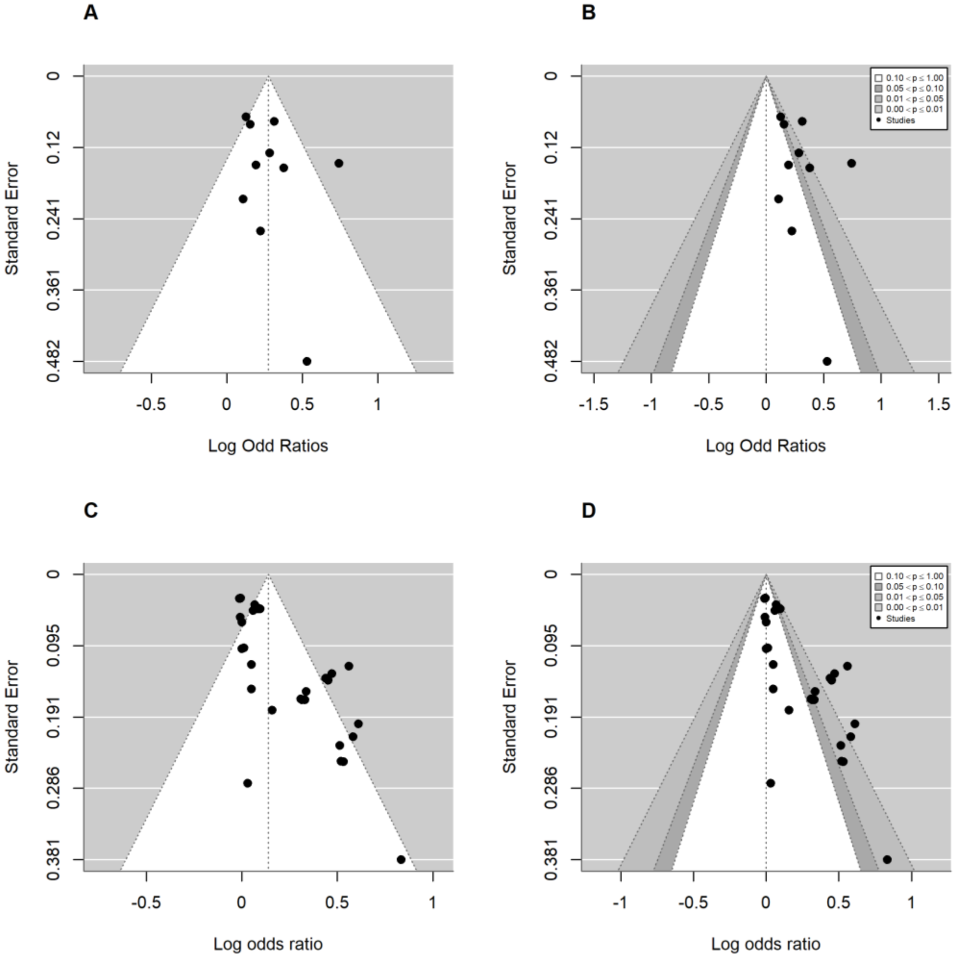

**Figure S4. Funnel plots of meta-analyses of prospective associations of pain and subsequent anxiety. (A) Funnel plot of unadjusted associations; (B) Contour-enhanced funnel plot of unadjusted associations; (C) Funnel plot of adjusted associations; (D) Contour-enhanced funnel plot of adjusted associations.**

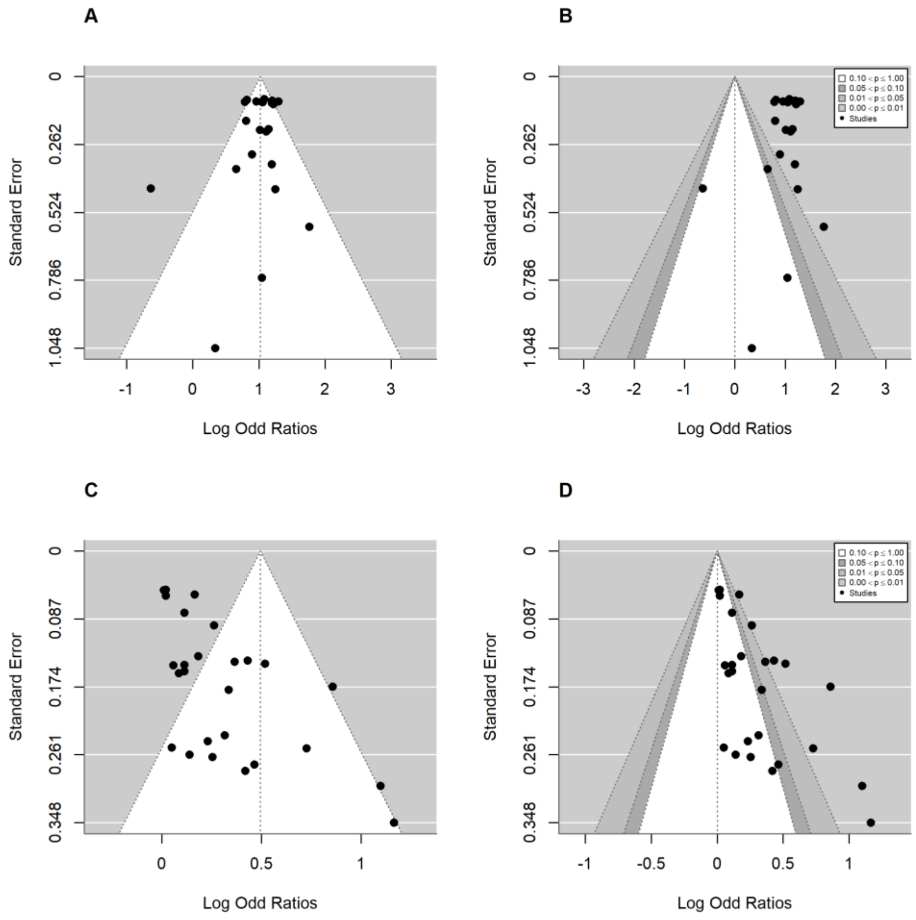

**Figure S5. Funnel plots of meta-analyses of prospective associations of pain and subsequent depression. (A) Funnel plot of unadjusted associations; (B) Contour-enhanced funnel plot of unadjusted associations; (C) Funnel plot of adjusted associations of continuous depression outcomes; (D) Contour-enhanced funnel plot of adjusted associations of continuous depression outcomes; (E) Funnel plot of adjusted associations of categorical depression outcomes; (F) Contour-enhanced funnel plot of adjusted associations of categorical depression outcomes.**

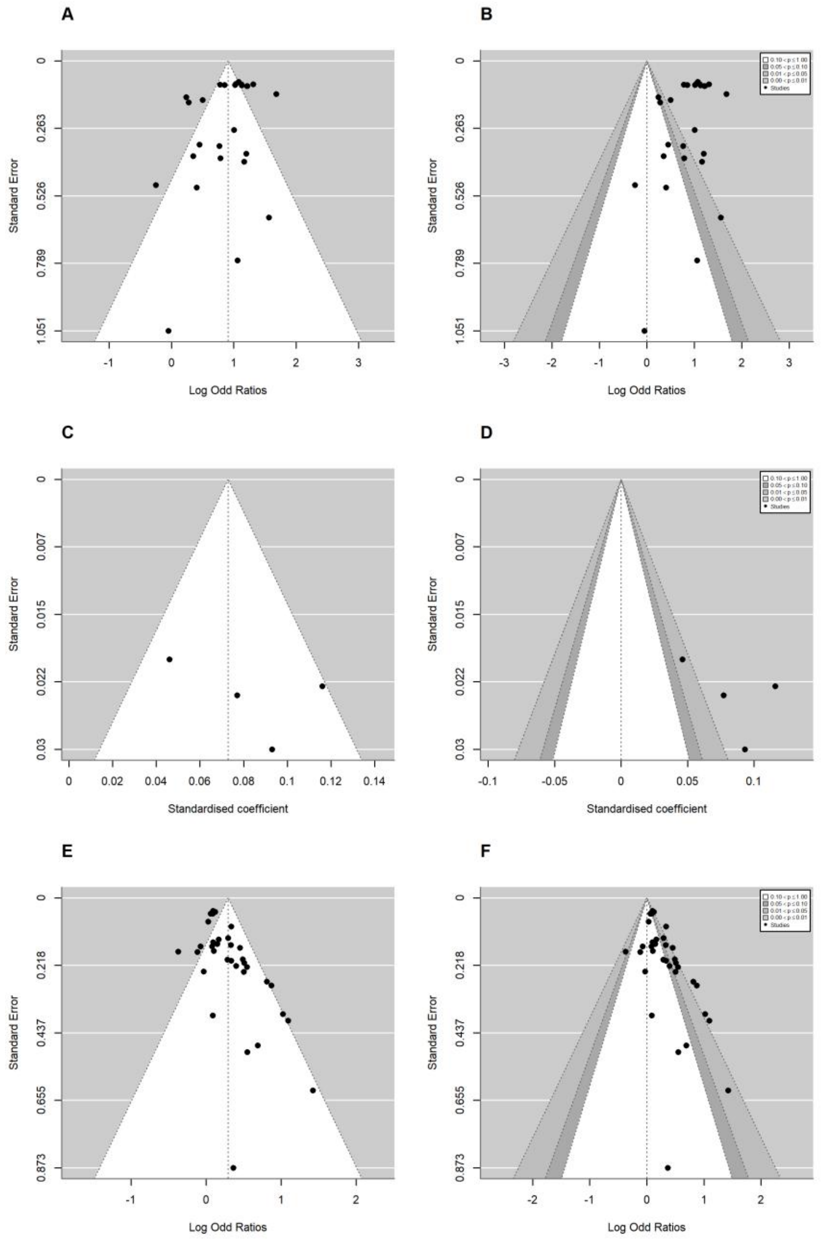
A
B
C
D
E
F
Standard Error
Log Odd Ratios
Standardised coefficient
0.10 < p ≤ 1.00
0.05 < p ≤ 0.10
0.01 < p ≤ 0.05
0.00 < p ≤ 0.01
Studies

**Figure S6. Funnel plots of meta-analyses of prospective associations of pain and subsequent combined anxiety/depression. (A) Funnel plot of unadjusted associations; (B) Contour-enhanced funnel plot of unadjusted associations; (C) Funnel plot of adjusted associations; (D) Contour-enhanced funnel plot of adjusted associations.**

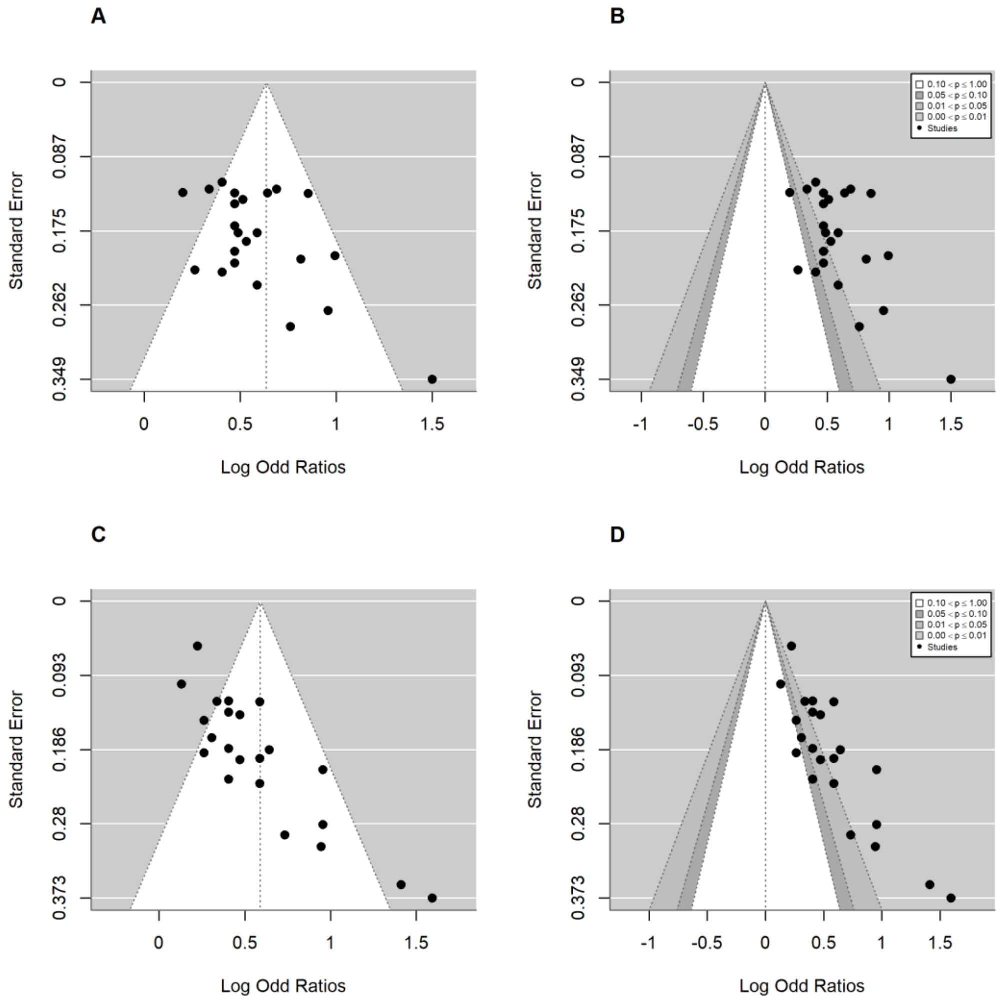

### 4. Meta regression results

**Table S1**

*Meta-Regression Results for Unadjusted Association between Anxiety and Subsequent Pain*

| Moderator | Subgroup | Number of effects | *b* | *SE* | *t-value* | *df* | *p* | *95% CI* | Between-group F-test |
|---|---|---|---|---|---|---|---|---|---|
| Pain type | | | | | | | | | *F(3, 20) = 0.36, p = 0.784* |
| | MSK | 12 | 0.40 | 0.18 | 2.19 | 20 | 0.041 | 0.02, 0.78 | |
| | Others | 3 | -0.04 | 0.13 | -0.27 | 20 | 0.787 | -0.31, 0.24 | |
| | Mixed pain | 5 | 0.32 | 0.36 | 0.91 | 20 | 0.375 | -0.42, 1.06 | |
| | Headache | 4 | 0.02 | 0.13 | 0.18 | 20 | 0.862 | -0.24, 0.29 | |
| Risk of bias | | | | | | | | | *F(2, 21) = 1.41, p = 0.266* |
| | low | 11 | 0.86 | 0.27 | 3.17 | 21 | 0.005 | 0.30, 1.43 | |
| | some concerns | 2 | -0.40 | 0.49 | -0.82 | 21 | 0.422 | -1.41, 0.61 | |
| | very high or high | 11 | -0.54 | 0.32 | -1.68 | 21 | 0.108 | -1.20, 0.13 | |

| | | | | | | | | | |
|---|---|---|---|---|---|---|---|---|---|
| Anxiety informant | | | | | | | | | *F(1, 21) = 1.17, p = 0.292* |
| | youth | 21 | 0.54 | 0.17 | 3.11 | 21 | 0.005 | 0.18, 0.9 | |
| | other | 2 | -0.50 | 0.46 | -1.08 | 21 | 0.292 | -1.45, 0.46 | |
| **Pain operationalisation** | | | | | | | | | ***F(1, 22) = 14.49, p = 0.001*** |
| | **continuous** | **15** | **0.91** | **0.15** | **6.27** | **22** | **<.001** | **0.61, 1.22** | |
| | **categorical** | **9** | **-0.70** | **0.18** | **-3.81** | **22** | **0.001** | **-1.08, -0.32** | |
| Mean baseline age | | | | | | | | | *F(1, 22) = 4.22, p = 0.052* |
| | intercept | | -0.24 | 0.37 | -0.63 | 22 | 0.535 | -1.01, 0.54 | |
| | mean baseline age | 24 | 0.06 | 0.03 | 2.05 | 22 | 0.052 | 0.00, 0.12 | |
| **Follow-up duration** | | | | | | | | | ***F(1, 22) = 6.34, p = 0.020*** |
| | **intercept** | | **0.62** | **0.16** | **3.79** | **22** | **0.001** | **0.28, 0.96** | |

| | | | | | | | | | |
|---|---|---|---|---|---|---|---|---|---|
| | **follow-up duration** | **24** | **-0.08** | **0.03** | **-2.52** | **22** | **0.020** | **-0.15, -0.01** | |
| **Sex** | | | | | | | | | ***F(1, 30) = 7.27, p = 0.011*** |
| | **female** | **17** | **0.62** | **0.21** | **2.93** | **30** | **0.006** | **0.19, 1.05** | |
| | **male** | **15** | **-0.22** | **0.08** | **-2.70** | **30** | **0.011** | **-0.38, -0.05** | |

**Table S2**

*Meta-Regression Results for Adjusted Association between Anxiety and Subsequent Categorical Pain*

| Moderator | Subgroup | Number of effects | *b* | *SE* | *t-value* | *df* | *p* | *95% CI* | Between-group F-test |
|---|---|---|---|---|---|---|---|---|---|
| Mean baseline age | | | | | | | | | *F(1, 5) = 0.02, p = 0.891* |
| | intercept | | 0.05 | 0.25 | 0.20 | 5 | 0.853 | -0.59, 0.68 | |
| | mean baseline age | 7 | 0.00 | 0.02 | 0.14 | 5 | 0.891 | -0.05, 0.06 | |
| Anxiety informant | | | | | | | | | *F(1, 4) = 0.64, p = 0.467* |
| | youth | 4 | 0.22 | 0.21 | 1.03 | 4 | 0.359 | -0.36, 0.79 | |
| | other | 2 | -0.25 | 0.32 | -0.80 | 4 | 0.467 | -1.13, 0.62 | |
| Anxiety operationalisation | | | | | | | | | *F(1, 5) = 1.84, p = 0.233* |
| | continuous | 5 | 0.07 | 0.06 | 1.18 | 5 | 0.292 | -0.08, 0.21 | |

| | | | | | | | | | |
|---|---|---|---|---|---|---|---|---|---|
| | categorical | 2 | 0.58 | 0.42 | 1.36 | 5 | 0.233 | -0.51, 1.67 | |
| Pain type | | | | | | | | | *F(2, 4) = 1.19, p = 0.392* |
| | MSK | 2 | 0.64 | 0.42 | 1.52 | 4 | 0.204 | -0.53, 1.81 | |
| | Headache | 3 | -0.62 | 0.43 | -1.44 | 4 | 0.224 | -1.81, 0.58 | |
| | Abdominal pain | 2 | -0.53 | 0.43 | -1.22 | 4 | 0.289 | -1.73, 0.67 | |
| Follow-up duration | | | | | | | | | *F(1, 5) = 0, p = 0.997* |
| | intercept | | 0.09 | 0.13 | 0.64 | 5 | 0.552 | -0.26, 0.43 | |
| | follow-up duration | 7 | 0.00 | 0.04 | 0.00 | 5 | 0.997 | -0.11, 0.10 | |

**Table S3**

*Meta-Regression Results for Unadjusted Association between Depression and Subsequent Pain*

| Moderator | Subgroup | Number of effects | *b* | *SE* | *t-value* | *df* | *p* | *95% CI* | Between-group F-test |
|---|---|---|---|---|---|---|---|---|---|
| Mean baseline age | | | | | | | | | *F(1, 26) = 3.51, p = 0.072* |
| | intercept | | -0.02 | 0.40 | -0.05 | 26 | 0.964 | -0.84, 0.8 | |
| | mean baseline age | 28 | 0.05 | 0.03 | 1.87 | 26 | 0.072 | -0.01, 0.11 | |
| **Follow-up duration** | | | | | | | | | ***F(1, 26) = 5.85, p = 0.023*** |
| | **intercept** | | **0.78** | **0.20** | **3.88** | **26** | **0.001** | **0.37, 1.2** | |
| | **follow-up duration** | **28** | **-0.07** | **0.03** | **-2.42** | **26** | **0.023** | **-0.14, -0.01** | |

| | | | | | | | | | |
|---|---|---|---|---|---|---|---|---|---|
| Depression informant | | | | | | | | | *F(1, 25) = 0.47, p = 0.498* |
| | youth | 25 | 0.70 | 0.24 | 2.97 | 25 | 0.007 | 0.22, 1.19 | |
| | other | 2 | -0.50 | 0.73 | -0.69 | 25 | 0.498 | -2.00, 1.00 | |
| Depression operationalisation | | | | | | | | | *F(1, 26) = 1.04, p = 0.317* |
| | continuous | 25 | 0.57 | 0.20 | 2.78 | 26 | 0.010 | 0.15, 0.99 | |
| | categorical | 3 | 0.70 | 0.69 | 1.02 | 26 | 0.317 | -0.71, 2.11 | |
| Pain informant | | | | | | | | | *F(1, 26) = 1.16, p = 0.291* |
| | youth | 24 | 0.64 | 0.20 | 3.24 | 26 | 0.003 | 0.23, 1.04 | |
| | other | 4 | -0.03 | 0.03 | -1.08 | 26 | 0.291 | -0.10, 0.03 | |
| **Pain operationalisation** | | | | | | | | | ***F(1, 26) = 16.1, p = 0.001*** |
| | **continuous** | **11** | **1.61** | **0.28** | **5.83** | **26** | **<.001** | **1.05, 2.18** | |
| | **categorical** | **17** | **-1.24** | **0.31** | **-4.01** | **26** | **0.001** | **-1.87, -0.60** | |
| Pain type | | | | | | | | | *F(2, 24) = 0.6, p = 0.555* |

| | | | | | | | | |
|---|---|---|---|---|---|---|---|---|
| | MSK | 17 | 0.47 | 0.16 | 3.00 | 24 | 0.006 | 0.15, 0.79 | |
| | Others | 3 | 0.04 | 0.04 | 0.99 | 24 | 0.334 | -0.04, 0.12 | |
| | Headache | 7 | 0.03 | 0.04 | 0.79 | 24 | 0.436 | -0.05, 0.11 | |
| Risk of bias | | | | | | | | | *F(2, 25) = 1.86, p = 0.177* |
| | low | 18 | 1.00 | 0.26 | 3.80 | 25 | 0.001 | 0.46, 1.54 | |
| | some concerns | 2 | -0.71 | 0.64 | -1.11 | 25 | 0.278 | -2.04, 0.61 | |
| | very high or high | 8 | -0.69 | 0.38 | -1.83 | 25 | 0.080 | -1.47, 0.09 | |
| **Sex** | | | | | | | | | ***F(1, 26) = 12.31, p = 0.002*** |
| | **female** | **15** | **0.94** | **0.31** | **3.00** | **26** | **0.006** | **0.30, 1.58** | |
| | **male** | **13** | **-0.24** | **0.07** | **-3.51** | **26** | **0.002** | **-0.38, -0.10** | |

**Table S4**

*Meta-Regression Results for Adjusted Association between Depression and Subsequent Continuous Pain*

| Moderator | Subgroup | Number of effects | *b* | *SE* | *t-value* | *df* | *p* | *95% CI* | Between-group F-test |
|---|---|---|---|---|---|---|---|---|---|
| Mean baseline age | | | | | | | | | *F(1, 6) = 0.52, p = 0.496* |
| | intercept | | 0.41 | 0.38 | 1.06 | 6 | 0.330 | -0.53, 1.35 | |
| | mean baseline age | 8 | -0.02 | 0.03 | -0.72 | 6 | 0.496 | -0.09, 0.05 | |
| Depression informant | | | | | | | | | *F(1, 6) = 0.58, p = 0.474* |
| | youth | 6 | 0.11 | 0.06 | 1.67 | 6 | 0.147 | -0.05, 0.27 | |
| | other | 2 | 0.10 | 0.14 | 0.76 | 6 | 0.474 | -0.23, 0.43 | |
| Pain informant | | | | | | | | | *F(1, 6) = 0.58, p = 0.474* |
| | youth | 6 | 0.11 | 0.06 | 1.67 | 6 | 0.147 | -0.05, 0.27 | |

| | | | | | | | | | |
|---|---|---|---|---|---|---|---|---|
| | other | 2 | 0.10 | 0.14 | 0.76 | 6 | 0.474 | -0.23, 0.43 | |
| Pain type | | | | | | | | | *F(1, 6) = 0.22, p = 0.654* |
| | MSK | 4 | 0.11 | 0.08 | 1.30 | 6 | 0.240 | -0.09, 0.30 | |
| | Mixed pain | 4 | 0.05 | 0.12 | 0.47 | 6 | 0.654 | -0.23, 0.34 | |
| Risk of bias | | | | | | | | | *F(1, 6) = 0.58, p = 0.474* |
| | low | 6 | 0.11 | 0.06 | 1.67 | 6 | 0.147 | -0.05, 0.27 | |
| | very high | 2 | 0.10 | 0.14 | 0.76 | 6 | 0.474 | -0.23, 0.43 | |
| Sex | | | | | | | | | *F(1, 8) = 0.01, p = 0.934* |
| | female | 5 | 0.13 | 0.06 | 2.05 | 8 | 0.075 | -0.02, 0.28 | |
| | male | 5 | 0.00 | 0.02 | -0.09 | 8 | 0.934 | -0.05, 0.05 | |

**Table S5**

*Meta-Regression Results for Adjusted Association between Depression and Subsequent Categorical Pain*

| Moderator | Subgroup | Number of effects | *b* | *SE* | *t-value* | *df* | *p* | *95% CI* | Between-group F-test |
|---|---|---|---|---|---|---|---|---|---|
| Mean baseline age | | | | | | | | | *F(1, 10) = 0.06, p = 0.813* |
| | intercept | | 0.19 | 0.36 | 0.53 | 10 | 0.607 | -0.61, 1.00 | |
| | mean baseline age | 12 | 0.01 | 0.03 | 0.24 | 10 | 0.813 | -0.06, 0.08 | |
| Age group | | | | | | | | | *F(1, 10) = 0, p = 0.946* |
| | adolescents | 9 | 0.27 | 0.17 | 1.60 | 10 | 0.141 | -0.11, 0.64 | |
| | child | 3 | 0.02 | 0.28 | 0.07 | 10 | 0.946 | -0.61, 0.65 | |
| Depression informant | | | | | | | | | *F(1, 9) = 0.74, p = 0.413* |
| | youth | 7 | 0.40 | 0.21 | 1.91 | 9 | 0.088 | -0.07, 0.88 | |

| | | | | | | | | | |
|---|---|---|---|---|---|---|---|---|
| | other | 4 | -0.27 | 0.31 | -0.86 | 9 | 0.413 | -0.97, 0.44 | |
| **Depression operationalisation** | | | | | | | | | ***F(1, 10) = 7.68, p = 0.020*** |
| | **continuous** | **6** | **0.11** | **0.09** | **1.22** | **10** | **0.251** | **-0.09, 0.31** | |
| | **categorical** | **6** | **0.43** | **0.16** | **2.77** | **10** | **0.020** | **0.08, 0.78** | |
| Pain informant | | | | | | | | | *F(1, 10) = 0.23, p = 0.645* |
| | youth | 10 | 0.26 | 0.12 | 2.07 | 10 | 0.065 | -0.02, 0.54 | |
| | other | 2 | 0.11 | 0.24 | 0.48 | 10 | 0.645 | -0.42, 0.65 | |
| Pain type | | | | | | | | | *F(2, 8) = 0.16, p = 0.859* |
| | Headache | 4 | 0.23 | 0.15 | 1.53 | 8 | 0.165 | -0.12, 0.58 | |
| | Abdominal pain | 3 | 0.03 | 0.19 | 0.18 | 8 | 0.862 | -0.41, 0.48 | |
| | Mixed pain | 4 | 0.09 | 0.17 | 0.54 | 8 | 0.601 | -0.29, 0.47 | |
| Risk of bias | | | | | | | | | *F(1, 10) = 4.78, p = 0.054* |
| | low | 7 | 0.44 | 0.12 | 3.72 | 10 | 0.004 | 0.18, 0.71 | |
| | high | 5 | -0.38 | 0.17 | -2.19 | 10 | 0.054 | -0.76, 0.01 | |

| | | | | | | | | | |
|---|---|---|---|---|---|---|---|---|---|
| Follow-up duration | | | | | | | | | *F(1, 10) = 0.15, p = 0.704* |
| | intercept | | 0.35 | 0.23 | 1.49 | 10 | 0.167 | -0.17, 0.87 | |
| | follow-up duration | 12 | -0.03 | 0.08 | -0.39 | 10 | 0.704 | -0.20, 0.14 | |

**Table S6**

*Meta-Regression Results for Unadjusted Association between Combined Anxiety/Depression and Subsequent Pain*

| Moderator | Subgroup | Number of effects | *b* | *SE* | *t-value* | *df* | *p* | *95% CI* | Between-group F-test |
|---|---|---|---|---|---|---|---|---|---|
| Mean baseline age | | | | | | | | | *F(1, 8) = 0.01, p = 0.930* |
| | intercept | | 0.27 | 0.25 | 1.10 | 8 | 0.304 | -0.30, 0.85 | |
| | mean baseline age | 10 | 0.00 | 0.02 | 0.09 | 8 | 0.930 | -0.05, 0.05 | |
| Age group | | | | | | | | | *F(1, 8) = 0.25, p = 0.632* |
| | adolescents | 6 | 0.26 | 0.12 | 2.18 | 8 | 0.061 | -0.02, 0.53 | |
| | child | 4 | 0.09 | 0.17 | 0.50 | 8 | 0.632 | -0.31, 0.49 | |
| **Follow-up duration** | | | | | | | | | ***F(1, 8) = 5.44, p = 0.048*** |
| | **intercept** | | **0.73** | **0.21** | **3.53** | **8** | **0.008** | **0.25, 1.21** | |
| | **follow-up duration** | **10** | **-0.17** | **0.07** | **-2.33** | **8** | **0.048** | **-0.34, 0.00** | |

| | | | | | | | | | |
|---|---|---|---|---|---|---|---|---|---|
| Anxiety/depression informant | | | | | | | | | *F(1, 8) = 0, p = 0.964* |
| | youth | 4 | 0.29 | 0.16 | 1.79 | 8 | 0.111 | -0.08, 0.67 | |
| | other | 6 | 0.01 | 0.19 | 0.05 | 8 | 0.964 | -0.44, 0.46 | |
| Pain type | | | | | | | | | *F(3, 6) = 0.3, p = 0.825* |
| | MSK | 4 | 0.30 | 0.19 | 1.54 | 6 | 0.174 | -0.17, 0.77 | |
| | Mixed pain | 2 | 0.17 | 0.28 | 0.59 | 6 | 0.580 | -0.53, 0.86 | |
| | Headache | 2 | -0.08 | 0.30 | -0.26 | 6 | 0.800 | -0.82, 0.66 | |
| | Dysmenorrhea | 2 | -0.10 | 0.31 | -0.31 | 6 | 0.768 | -0.85, 0.66 | |
| Pain informant | | | | | | | | | *F(1, 8) = 4.03, p = 0.080* |
| | youth | 8 | 0.23 | 0.05 | 4.46 | 8 | 0.002 | 0.11, 0.34 | |
| | other | 2 | 0.29 | 0.14 | 2.01 | 8 | 0.080 | -0.04, 0.61 | |
| Risk of bias | | | | | | | | | *F(1, 8) = 3.91, p = 0.083* |
| | some concerns | 3 | 0.44 | 0.10 | 4.35 | 8 | 0.003 | 0.21, 0.67 | |
| | high | 7 | -0.23 | 0.12 | -1.98 | 8 | 0.083 | -0.50, 0.04 | |

**Table S7**

*Meta-Regression Results for Adjusted Association between Combined Anxiety/Depression and Subsequent Pain*

| Moderator | Subgroup | Number of effects | *b* | *SE* | *t-value* | *df* | *p* | *95% CI* | Between-group F-test |
|---|---|---|---|---|---|---|---|---|---|
| Mean baseline age | | | | | | | | | *F(1, 32) = 1.41, p = 0.244* |
| | intercept | | -0.03 | 0.16 | -0.18 | 32 | 0.860 | -0.35, 0.29 | |
| | mean baseline age | 34 | 0.02 | 0.02 | 1.19 | 32 | 0.244 | -0.01, 0.05 | |
| Age group | | | | | | | | | *F(1, 32) = 3.2, p = 0.083* |
| | adolescents | 20 | 0.26 | 0.09 | 2.84 | 32 | 0.008 | 0.07, 0.45 | |
| | child | 14 | -0.22 | 0.12 | -1.79 | 32 | 0.083 | -0.48, 0.03 | |
| Follow-up duration | | | | | | | | | *F(1, 32) = 2.27, p = 0.142* |
| | intercept | | 0.33 | 0.14 | 2.33 | 32 | 0.027 | 0.04, 0.61 | |
| | follow-up duration | 34 | -0.07 | 0.04 | -1.51 | 32 | 0.142 | -0.15, 0.02 | |
| Pain informant | | | | | | | | | *F(1, 32) = 0, p = 0.965* |

| | | | | | | | | | |
|---|---|---|---|---|---|---|---|---|---|
| | youth | 19 | 0.14 | 0.11 | 1.25 | 32 | 0.221 | -0.09, 0.37 | |
| | other | 15 | -0.01 | 0.16 | -0.04 | 32 | 0.965 | -0.33, 0.31 | |
| Pain type | | | | | | | | | *F(2, 30) = 0.55, p = 0.583* |
| | Headache | 27 | 0.22 | 0.11 | 2.00 | 30 | 0.054 | 0.00, 0.44 | |
| | Mixed pain | 3 | -0.16 | 0.19 | -0.82 | 30 | 0.417 | -0.55, 0.24 | |
| | Dysmenorrhea | 3 | -0.20 | 0.24 | -0.83 | 30 | 0.413 | -0.70, 0.30 | |
| **Risk of bias** | | | | | | | | | ***F(1, 32) = 14.57, p = 0.001*** |
| | **low** | **10** | **0.04** | **0.04** | **1.02** | **32** | **0.316** | **-0.04, 0.13** | |
| | **high** | **24** | **0.22** | **0.06** | **3.82** | **32** | **0.001** | **0.10, 0.34** | |
| Anxiety/depression informant | | | | | | | | | *F(1, 32) = 3.81, p = 0.060* |
| | youth | 14 | 0.38 | 0.13 | 2.80 | 32 | 0.009 | 0.10, 0.65 | |
| | other | 20 | -0.29 | 0.15 | -1.95 | 32 | 0.060 | -0.59, 0.01 | |

**Table S8**

*Meta-Regression Results for Unadjusted Association between Pain and Subsequent Anxiety*

| Moderator | Subgroup | Number of effects | *b* | *SE* | *t-value* | *df* | *p* | *95% CI* | Between-group F-test |
|---|---|---|---|---|---|---|---|---|---|
| **Mean baseline age** | | | | | | | | | ***F(1, 20) = 13.33, p = 0.002*** |
| | **intercept** | | **-0.01** | **0.29** | **-0.03** | **20** | **0.979** | **-0.62, 0.60** | |
| | **mean baseline age** | **22** | **0.09** | **0.02** | **3.65** | **20** | **0.002** | **0.04, 0.14** | |
| **Follow-up duration** | | | | | | | | | ***F(1, 20) = 28.07, p < .001*** |
| | **intercept** | | **1.36** | **0.22** | **6.26** | **20** | **<.001** | **0.91, 1.81** | |
| | **follow-up duration** | **22** | **-0.13** | **0.02** | **-5.30** | **20** | **<.001** | **-0.18, -0.08** | |
| Pain type | | | | | | | | | *F(2, 19) = 1.67, p = 0.214* |

| | | | | | | | | |
|---|---|---|---|---|---|---|---|---|
| | MSK | 10 | 1.06 | 0.05 | 19.86 | 19 | <.001 | 0.95, 1.17 | |
| | Mixed pain | 3 | 0.03 | 0.15 | 0.17 | 19 | 0.867 | -0.30, 0.35 | |
| | Abdominal pain | 9 | -0.25 | 0.14 | -1.78 | 19 | 0.091 | -0.55, 0.04 | |
| Risk of bias | | | | | | | | | *F(1, 20) = 0.93, p = 0.346* |
| | low | 10 | 1.06 | 0.09 | 12.39 | 20 | <.001 | 0.88, 1.24 | |
| | high | 12 | -0.13 | 0.14 | -0.97 | 20 | 0.346 | -0.41, 0.15 | |
| Age group | | | | | | | | | *F(1, 20) = 4.32, p = 0.051* |
| | adolescents | 18 | 1.06 | 0.05 | 22.29 | 20 | <.001 | 0.96, 1.16 | |
| | child | 4 | -0.35 | 0.17 | -2.08 | 20 | 0.051 | -0.70, 0.00 | |
| Pain informant | | | | | | | | | *F(1, 20) = 3.38, p = 0.081* |
| | youth | 13 | 1.07 | 0.05 | 21.77 | 20 | <.001 | 0.96, 1.17 | |
| | other | 9 | -0.25 | 0.14 | -1.84 | 20 | 0.081 | -0.54, 0.03 | |
| Pain operationalisation | | | | | | | | | *F(1, 20) = 3.38, p = 0.081* |
| | continuous | 13 | 1.07 | 0.05 | 21.77 | 20 | <.001 | 0.96, 1.17 | |

| | | | | | | | | | |
|---|---|---|---|---|---|---|---|---|---|
| | categorical | 9 | -0.25 | 0.14 | -1.84 | 20 | 0.081 | -0.54, 0.03 | |
| Anxiety operationalisation | | | | | | | | | *F(1, 20) = 3.38, p = 0.081* |
| | continuous | 13 | 1.07 | 0.05 | 21.77 | 20 | <.001 | 0.96, 1.17 | |
| | categorical | 9 | -0.25 | 0.14 | -1.84 | 20 | 0.081 | -0.54, 0.03 | |
| **Sex** | | | | | | | | | ***F(1, 24) = 7.81, p = 0.010*** |
| | **female** | **13** | **1.02** | **0.08** | **12.16** | **24** | **<.001** | **0.84, 1.19** | |
| | **male** | **13** | **-0.18** | **0.06** | **-2.79** | **24** | **0.010** | **-0.31, -0.05** | |

**Table S9**

*Meta-Regression Results for Adjusted Association between Pain and Subsequent Anxiety*

| Moderator | Subgroup | Number of effects | *b* | *SE* | *t-value* | *df* | *p* | *95% CI* | Between-group F-test |
|---|---|---|---|---|---|---|---|---|---|
| Mean baseline age | | | | | | | | | *F(1, 24) = 0.87, p = 0.361* |
| | intercept | | 0.84 | 0.42 | 2.01 | 24 | 0.056 | -0.02, 1.69 | |
| | mean baseline age | 26 | -0.03 | 0.03 | -0.93 | 24 | 0.361 | -0.09, 0.04 | |
| Follow-up duration | | | | | | | | | *F(1, 24) = 0.42, p = 0.522* |
| | intercept | | 0.43 | 0.21 | 2.03 | 24 | 0.054 | -0.01, 0.86 | |
| | follow-up duration | 26 | 0.02 | 0.03 | 0.65 | 24 | 0.522 | -0.04, 0.08 | |
| **Age group** | | | | | | | | | ***F(1, 24) = 5.45, p = 0.028*** |

| | | | | | | | | |
|---|---|---|---|---|---|---|---|---|
| | **adolescents** | **24** | **0.29** | **0.12** | **2.38** | **24** | **0.026** | **0.04, 0.55** | |
| | **child** | **2** | **0.57** | **0.25** | **2.33** | **24** | **0.028** | **0.07, 1.08** | |
| **Pain informant** | | | | | | | | | ***F(1, 24) = 5.49, p = 0.028*** |
| | **youth** | **22** | **0.17** | **0.17** | **0.99** | **24** | **0.331** | **-0.18, 0.52** | |
| | **other** | **4** | **0.58** | **0.25** | **2.34** | **24** | **0.028** | **0.07, 1.10** | |
| Pain operationalisation | | | | | | | | | *F(1, 24) = 1.54, p = 0.226* |
| | continuous | 6 | 0.08 | 0.38 | 0.22 | 24 | 0.831 | -0.69, 0.86 | |
| | categorical | 20 | 0.53 | 0.43 | 1.24 | 24 | 0.226 | -0.35, 1.42 | |
| Pain type | | | | | | | | | *F(3, 22) = 1.45, p = 0.256* |
| | MSK | 3 | 0.04 | 0.35 | 0.11 | 22 | 0.914 | -0.68, 0.76 | |
| | Mixed pain | 3 | 0.09 | 0.07 | 1.30 | 22 | 0.208 | -0.05, 0.24 | |
| | Headache | 9 | 0.52 | 0.40 | 1.30 | 22 | 0.208 | -0.31, 1.35 | |
| | Abdominal pain | 11 | 0.60 | 0.40 | 1.52 | 22 | 0.142 | -0.22, 1.43 | |
| Risk of bias | | | | | | | | | *F(1, 24) = 1.99, p = 0.171* |

| | | | | | | | | | |
|---|---|---|---|---|---|---|---|---|---|
| | some concerns | 14 | 0.40 | 0.18 | 2.23 | 24 | 0.036 | 0.03, 0.77 | |
| | high | 12 | 0.13 | 0.09 | 1.41 | 24 | 0.171 | -0.06, 0.33 | |
| Anxiety informant | | | | | | | | | *F(1, 24) = 0.12, p = 0.730* |
| | youth | 24 | 0.44 | 0.27 | 1.63 | 24 | 0.116 | -0.12, 1.01 | |
| | other | 2 | 0.15 | 0.44 | 0.35 | 24 | 0.730 | -0.76, 1.07 | |

**Table S10**

*Meta-Regression Results for Unadjusted Association between Pain and Subsequent Depression*

| Moderator | Subgroup | Number of effects | *b* | *SE* | *t-value* | *df* | *p* | *95% CI* | Between-group F-test |
|---|---|---|---|---|---|---|---|---|---|
| **Mean baseline age** | | | | | | | | | ***F(1, 24) = 15.14, p = 0.001*** |
| | **intercept** | | **-0.44** | **0.42** | **-1.05** | **24** | **0.306** | **-1.32, 0.43** | |
| | **mean baseline age** | **26** | **0.11** | **0.03** | **3.89** | **24** | **0.001** | **0.05, 0.17** | |
| **Follow-up duration** | | | | | | | | | ***F(1, 24) = 16.62, p < .001*** |
| | **intercept** | | **1.22** | **0.27** | **4.45** | **24** | **<.001** | **0.65, 1.78** | |
| | **follow-up duration** | **26** | **-0.12** | **0.03** | **-4.08** | **24** | **<.001** | **-0.17, -0.06** | |
| Pain type | | | | | | | | | *F(3, 22) = 1.31, p = 0.297* |

| | | | | | | | | | |
|---|---|---|---|---|---|---|---|---|---|
| | MSK | 10 | 1.07 | 0.42 | 2.58 | 22 | 0.017 | 0.21, 1.94 | |
| | Mixed pain | 3 | 0.06 | 0.50 | 0.12 | 22 | 0.903 | -0.98, 1.10 | |
| | Headache | 2 | -0.22 | 0.52 | -0.42 | 22 | 0.679 | -1.29, 0.86 | |
| | Abdominal pain | 11 | -0.42 | 0.49 | -0.85 | 22 | 0.406 | -1.44, 0.60 | |
| **Depression operationalisation** | | | | | | | | | ***F(1, 24) = 7.07, p = 0.014*** |
| | **continuous** | **11** | **1.32** | **0.21** | **6.38** | **24** | **<.001** | **0.89, 1.74** | |
| | **categorical** | **15** | **-0.76** | **0.28** | **-2.66** | **24** | **0.014** | **-1.34, -0.17** | |
| Age group | | | | | | | | | *F(1, 24) = 1.67, p = 0.209* |
| | adolescents | 22 | 0.98 | 0.22 | 4.38 | 24 | <.001 | 0.52, 1.44 | |
| | child | 4 | -0.41 | 0.32 | -1.29 | 24 | 0.209 | -1.07, 0.25 | |
| Pain informant | | | | | | | | | *F(1, 24) = 0.79, p = 0.382* |
| | youth | 17 | 1.06 | 0.30 | 3.58 | 24 | 0.002 | 0.45, 1.67 | |
| | other | 9 | -0.46 | 0.51 | -0.89 | 24 | 0.382 | -1.52, 0.60 | |

| | | | | | | | | | |
|---|---|---|---|---|---|---|---|---|---|
| Pain operationalisation | | | | | | | | | *F(1, 24) = 0.79, p = 0.382* |
| | continuous | 17 | 1.06 | 0.30 | 3.58 | 24 | 0.002 | 0.45, 1.67 | |
| | categorical | 9 | -0.46 | 0.51 | -0.89 | 24 | 0.382 | -1.52, 0.60 | |
| Risk of bias | | | | | | | | | *F(1, 24) = 0.79, p = 0.382* |
| | low | 17 | 1.06 | 0.30 | 3.58 | 24 | 0.002 | 0.45, 1.67 | |
| | high | 9 | -0.46 | 0.51 | -0.89 | 24 | 0.382 | -1.52, 0.60 | |
| Sex | | | | | | | | | *F(1, 20) = 2.39, p = 0.137* |
| | female | 13 | 0.33 | 0.08 | 4.11 | 20 | 0.001 | 0.16, 0.49 | |
| | male | 13 | -0.02 | 0.02 | -1.55 | 20 | 0.137 | -0.06, 0.01 | |

**Table S11**

*Meta-Regression Results for Adjusted Association between Pain and Subsequent Continuous Depression*

| Moderator | Subgroup | Number of effects | *b* | *SE* | *t-value* | *df* | *p* | *95% CI* | Between-group F-test |
|---|---|---|---|---|---|---|---|---|---|
| Sex | | | | | | | | | *F(1, 8) = 0.13, p = 0.733* |
| | female | 5 | 0.07 | 0.02 | 3.55 | 8 | 0.008 | 0.02, 0.12 | |
| | male | 5 | 0.01 | 0.03 | 0.35 | 8 | 0.733 | -0.05, 0.07 | |

**Table S12**

*Meta-Regression Results for Adjusted Association between Pain and Subsequent Categorical Depression*

| Moderator | Subgroup | Number of effects | *b* | *SE* | *t-value* | *df* | *p* | *95% CI* | Between-group F-test |
|---|---|---|---|---|---|---|---|---|---|
| Mean baseline age | | | | | | | | | *F(1, 32) = 3.15, p = 0.086* |
| | intercept | | 1.10 | 0.48 | 2.32 | 32 | 0.027 | 0.13, 2.07 | |
| | mean baseline age | 34 | -0.06 | 0.03 | -1.77 | 32 | 0.086 | -0.12, 0.01 | |
| Follow-up duration | | | | | | | | | *F(1, 32) = 0.88, p = 0.356* |
| | intercept | | 0.20 | 0.15 | 1.36 | 32 | 0.183 | -0.10, 0.50 | |
| | follow-up duration | 34 | 0.03 | 0.04 | 0.94 | 32 | 0.356 | -0.04, 0.10 | |
| Pain type | | | | | | | | | *F(3, 30) = 0.33, p = 0.805* |

| | | | | | | | | | |
|---|---|---|---|---|---|---|---|---|---|
| | MSK | 3 | 0.30 | 0.20 | 1.49 | 30 | 0.148 | -0.11, 0.72 | |
| | Mixed pain | 5 | 0.11 | 0.14 | 0.77 | 30 | 0.447 | -0.18, 0.39 | |
| | Headache | 11 | 0.03 | 0.22 | 0.15 | 30 | 0.879 | -0.42, 0.49 | |
| | Abdominal pain | 15 | -0.02 | 0.23 | -0.08 | 30 | 0.937 | -0.48, 0.44 | |
| Risk of bias | | | | | | | | | *F(2, 31) = 0.02, p = 0.984* |
| | low | 6 | 0.36 | 0.26 | 1.37 | 31 | 0.182 | -0.18, 0.89 | |
| | some concerns | 14 | -0.04 | 0.30 | -0.14 | 31 | 0.887 | -0.66, 0.57 | |
| | high | 14 | -0.03 | 0.30 | -0.09 | 31 | 0.926 | -0.64, 0.58 | |
| Age group | | | | | | | | | *F(1, 32) = 0.13, p = 0.720* |
| | adolescents | 32 | 0.28 | 0.09 | 3.16 | 32 | 0.003 | 0.10, 0.47 | |
| | child | 2 | 0.17 | 0.47 | 0.36 | 32 | 0.720 | -0.79, 1.13 | |
| Depression informant | | | | | | | | | *F(1, 32) = 0.02, p = 0.883* |
| | youth | 32 | 0.31 | 0.11 | 2.66 | 32 | 0.012 | 0.07, 0.54 | |

| | | | | | | | | | |
|---|---|---|---|---|---|---|---|---|---|
| | other | 2 | 0.04 | 0.30 | 0.15 | 32 | 0.883 | -0.56, 0.65 | |
| Pain informant | | | | | | | | | *F(1, 32) = 3.96, p = 0.055* |
| | youth | 28 | 0.21 | 0.06 | 3.26 | 32 | 0.003 | 0.08, 0.33 | |
| | other | 6 | 0.38 | 0.19 | 1.99 | 32 | 0.055 | -0.01, 0.77 | |
| Pain operationalisation | | | | | | | | | *F(1, 32) = 3.96, p = 0.055* |
| | continuous | 28 | 0.21 | 0.06 | 3.26 | 32 | 0.003 | 0.08, 0.33 | |
| | categorical | 6 | 0.38 | 0.19 | 1.99 | 32 | 0.055 | -0.01, 0.77 | |

**Table S13**

*Meta-Regression Results for Unadjusted Association between Pain and Subsequent Combined Anxiety/Depression*

| Moderator | Subgroup | Number of effects | *b* | *SE* | *t-value* | *df* | *p* | *95% CI* | Between-group F-test |
|---|---|---|---|---|---|---|---|---|---|
| Mean baseline age | | | | | | | | | *F(1, 21) = 0.75, p = 0.397* |
| | intercept | | 0.39 | 0.30 | 1.32 | 21 | 0.201 | -0.22, 1.01 | |
| | mean baseline age | 23 | 0.02 | 0.02 | 0.86 | 21 | 0.397 | -0.03, 0.07 | |
| Follow-up duration | | | | | | | | | *F(1, 21) = 0.83, p = 0.372* |
| | intercept | | 0.74 | 0.16 | 4.49 | 21 | <.001 | 0.40, 1.08 | |
| | follow-up duration | 23 | -0.02 | 0.02 | -0.91 | 21 | 0.372 | -0.06, 0.02 | |
| Age group | | | | | | | | | *F(1, 21) = 1.43, p = 0.246* |
| | adolescents | 19 | 0.73 | 0.14 | 5.01 | 21 | <.001 | 0.42, 1.03 | |
| | child | 4 | -0.33 | 0.27 | -1.19 | 21 | 0.246 | -0.90, 0.24 | |

| | | | | | | | | | |
|---|---|---|---|---|---|---|---|---|---|
| Pain informant | | | | | | | | | *F(1, 21) = 1.43, p = 0.246* |
| | youth | 19 | 0.73 | 0.14 | 5.01 | 21 | <.001 | 0.42, 1.03 | |
| | other | 4 | -0.33 | 0.27 | -1.19 | 21 | 0.246 | -0.90, 0.24 | |
| Pain type | | | | | | | | | *F(2, 20) = 0.33, p = 0.720* |
| | MSK | 12 | 0.70 | 0.15 | 4.79 | 20 | <.001 | 0.39, 1.00 | |
| | Headache | 2 | -0.02 | 0.13 | -0.14 | 20 | 0.889 | -0.30, 0.26 | |
| | Others | 9 | -0.10 | 0.12 | -0.82 | 20 | 0.424 | -0.34, 0.15 | |
| Risk of bias | | | | | | | | | *F(1, 21) = 0.01, p = 0.926* |
| | some concerns | 6 | 0.67 | 0.24 | 2.75 | 21 | 0.012 | 0.16, 1.18 | |
| | high | 17 | -0.03 | 0.33 | -0.09 | 21 | 0.926 | -0.72, 0.66 | |
| **Anxiety/depression operationalisation** | | | | | | | | | ***F(1, 21) = 11.02, p = 0.003*** |
| | **continuous** | **5** | **0.82** | **0.09** | **9.03** | **21** | **<.001** | **0.63, 1.01** | |
| | **categorical** | **18** | **-0.35** | **0.11** | **-3.32** | **21** | **0.003** | **-0.57, -0.13** | |

**Table S14**

*Meta-Regression Results for Adjusted Association between Pain and Subsequent Combined Anxiety/Depression*

| Moderator | Subgroup | Number of effects | *b* | *SE* | *t-value* | *df* | *p* | *95% CI* | Between-group F-test |
|---|---|---|---|---|---|---|---|---|---|
| Mean baseline age | | | | | | | | | *F(1, 21) = 0.56, p = 0.461* |
| | intercept | | 0.34 | 0.39 | 0.88 | 21 | 0.387 | -0.46, 1.15 | |
| | mean baseline age | 23 | 0.02 | 0.03 | 0.75 | 21 | 0.461 | -0.04, 0.08 | |
| Age group | | | | | | | | | *F(1, 21) = 1.02, p = 0.325* |
| | adolescents | 18 | 0.77 | 0.31 | 2.50 | 21 | 0.021 | 0.13, 1.42 | |
| | child | 5 | -0.53 | 0.52 | -1.01 | 21 | 0.325 | -1.62, 0.56 | |
| **Follow-up duration** | | | | | | | | | ***F(1, 21) = 9.94, p = 0.005*** |

| | | | | | | | | |
|---|---|---|---|---|---|---|---|---|
| | **intercept** | | **0.64** | **0.07** | **8.73** | **21** | **<.001** | **0.49, 0.79** | |
| | **follow-up duration** | **23** | **-0.03** | **0.01** | **-3.15** | **21** | **0.005** | **-0.05, -0.01** | |
| Pain informant | | | | | | | | | *F(1, 21) = 9.39, p = 0.006* |
| | youth | 18 | 0.55 | 0.05 | 10.22 | 21 | <.001 | 0.44, 0.66 | |
| | other | 5 | -0.28 | 0.09 | -3.07 | 21 | 0.006 | -0.47, -0.09 | |
| Pain operationalisation | | | | | | | | | *F(1, 21) = 9.39, p = 0.006* |
| | continuous | 5 | 0.27 | 0.07 | 3.62 | 21 | 0.002 | 0.11, 0.42 | |
| | categorical | 18 | 0.28 | 0.09 | 3.07 | 21 | 0.006 | 0.09, 0.47 | |
| Pain type | | | | | | | | | *F(2, 20) = 0.67, p = 0.521* |
| | MSK | 14 | 0.64 | 0.22 | 2.92 | 20 | 0.009 | 0.18, 1.10 | |
| | Headache | 2 | 0.03 | 0.13 | 0.22 | 20 | 0.828 | -0.24, 0.30 | |
| | Abdominal pain | 7 | -0.13 | 0.13 | -1.07 | 20 | 0.295 | -0.40, 0.13 | |
| Risk of bias | | | | | | | | | *F(1, 21) = 0.06, p = 0.813* |

| | | | | | | | | |
|---|---|---|---|---|---|---|---|---|
| | some concerns | 9 | 0.66 | 0.43 | 1.54 | 21 | 0.138 | -0.23, 1.55 |
| | high | 14 | -0.18 | 0.73 | -0.24 | 21 | 0.813 | -1.70, 1.35 |

### 5. Risk-of-bias plots

**Figure S7. Risk-of-bias plots for studies on the prospective associations between anxiety and subsequent pain.**

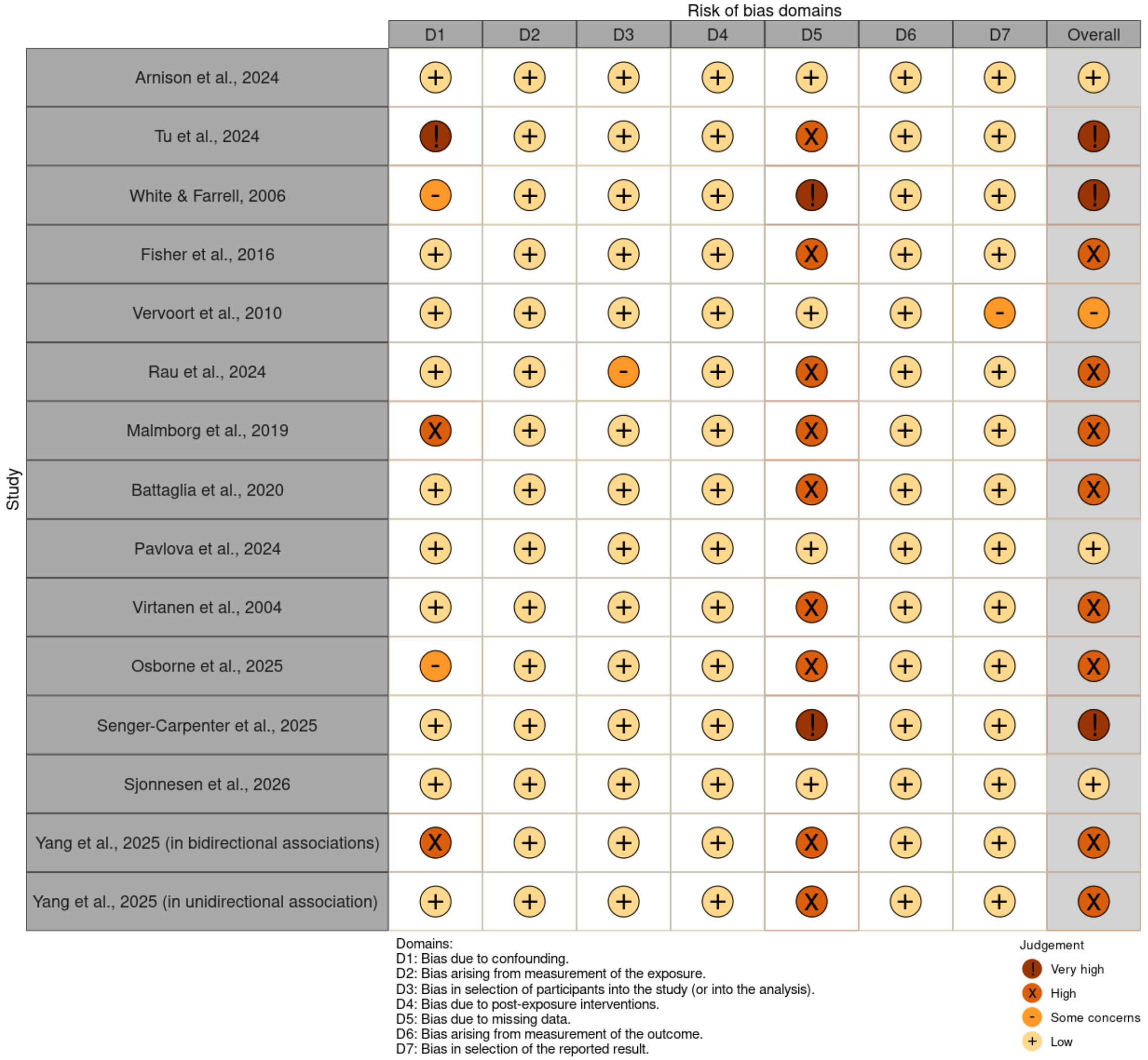

**Figure S8. Risk-of-bias plots for studies on the prospective associations between depression and subsequent pain.**

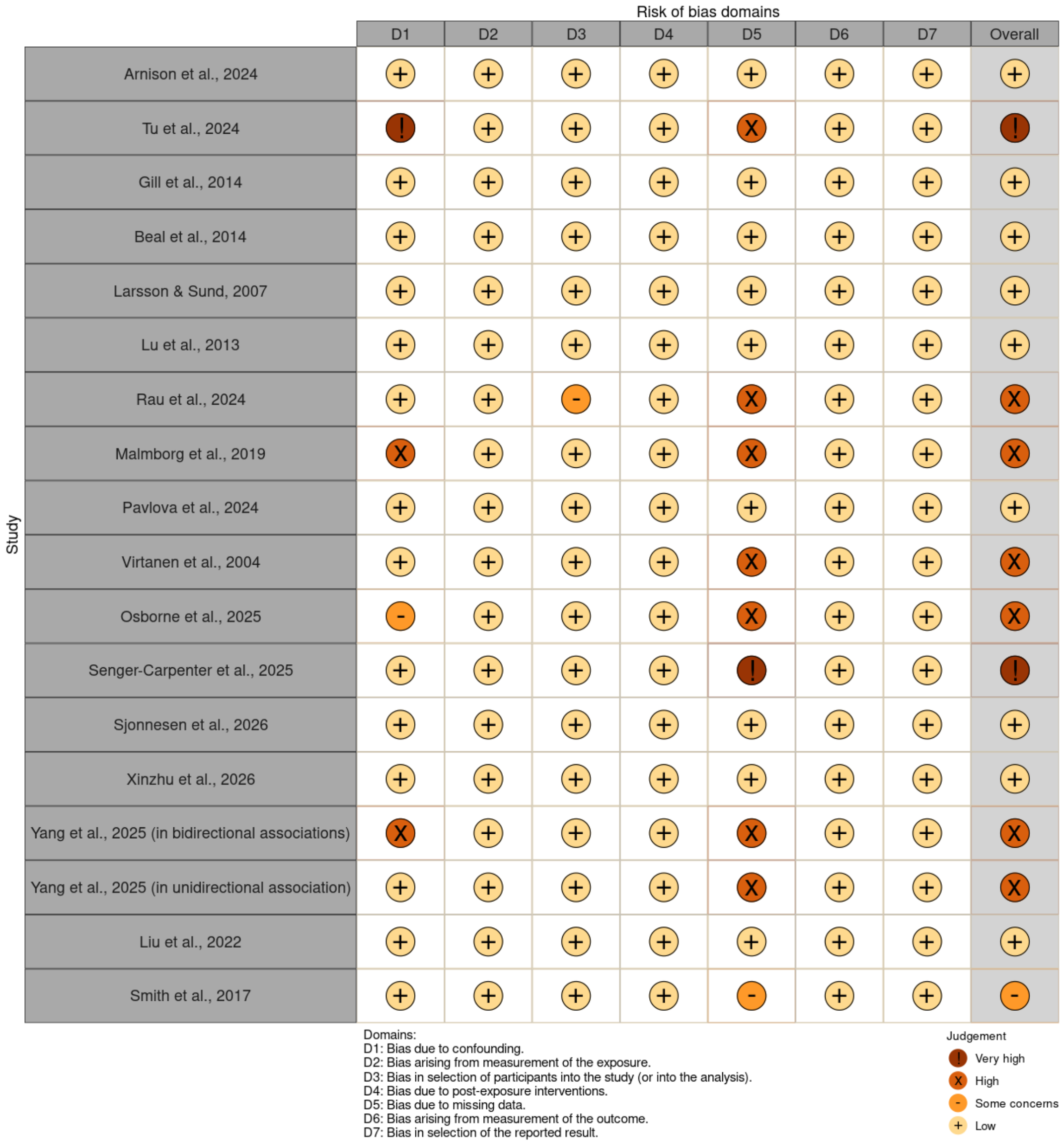

**Figure S9. Risk-of-bias plots for studies on the prospective associations between combined anxiety/depression and subsequent pain.**

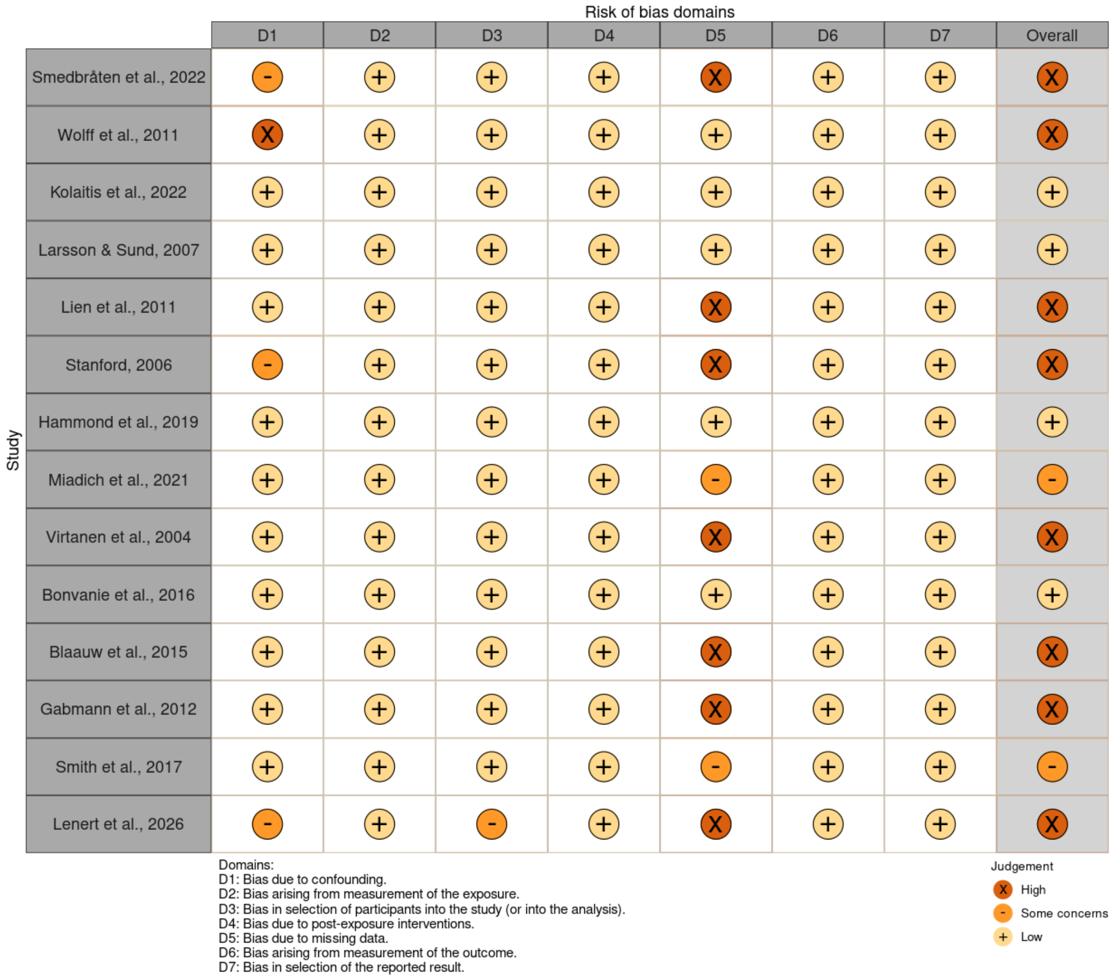

**Figure S10. Risk-of-bias plots for studies on the prospective associations between pain and subsequent anxiety.**

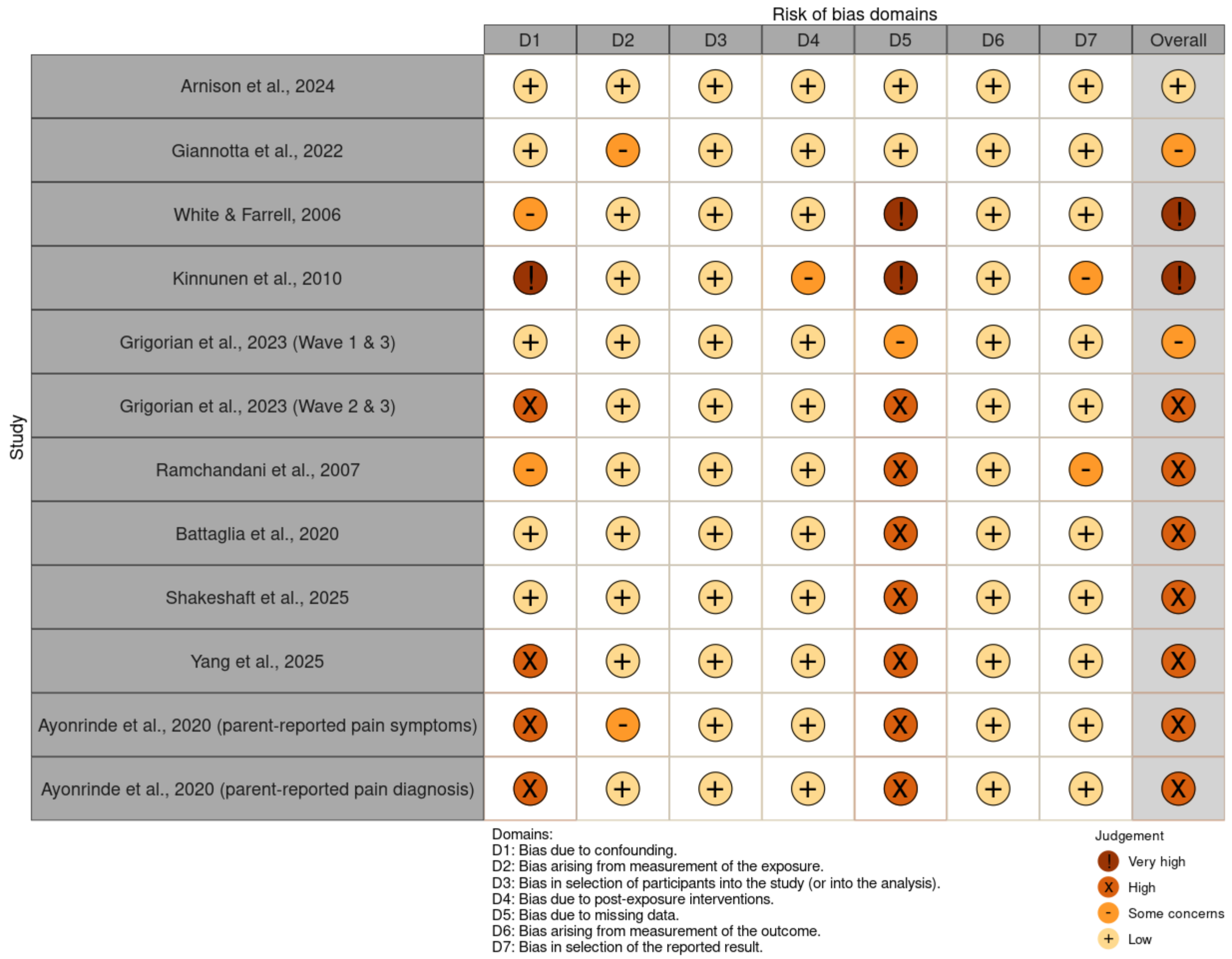

**Figure S11. Risk-of-bias plots for studies on the prospective associations between pain and subsequent depression.**

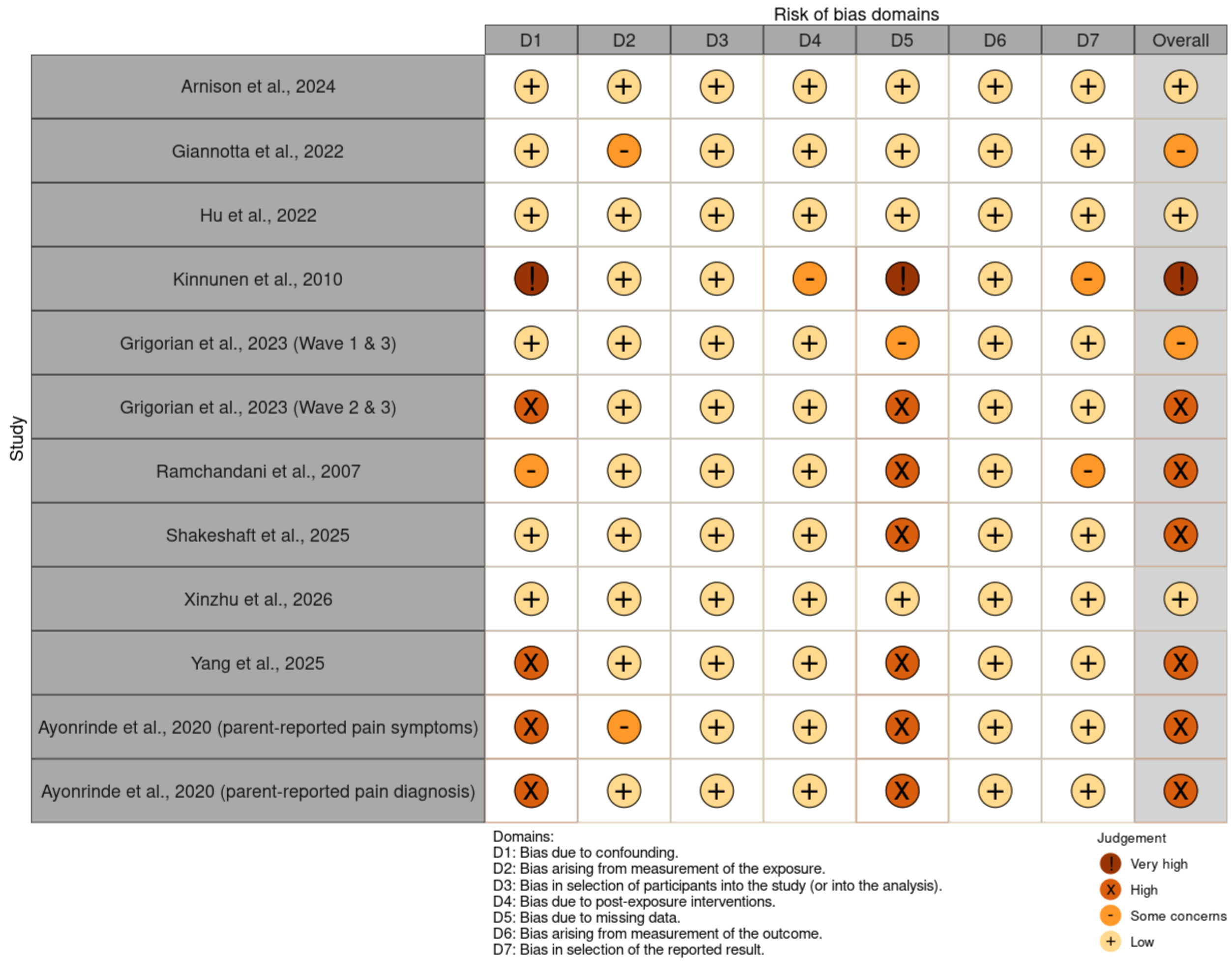

**Figure S12. Risk-of-bias plots for studies on the prospective associations between pain and subsequent combined anxiety/depression.**

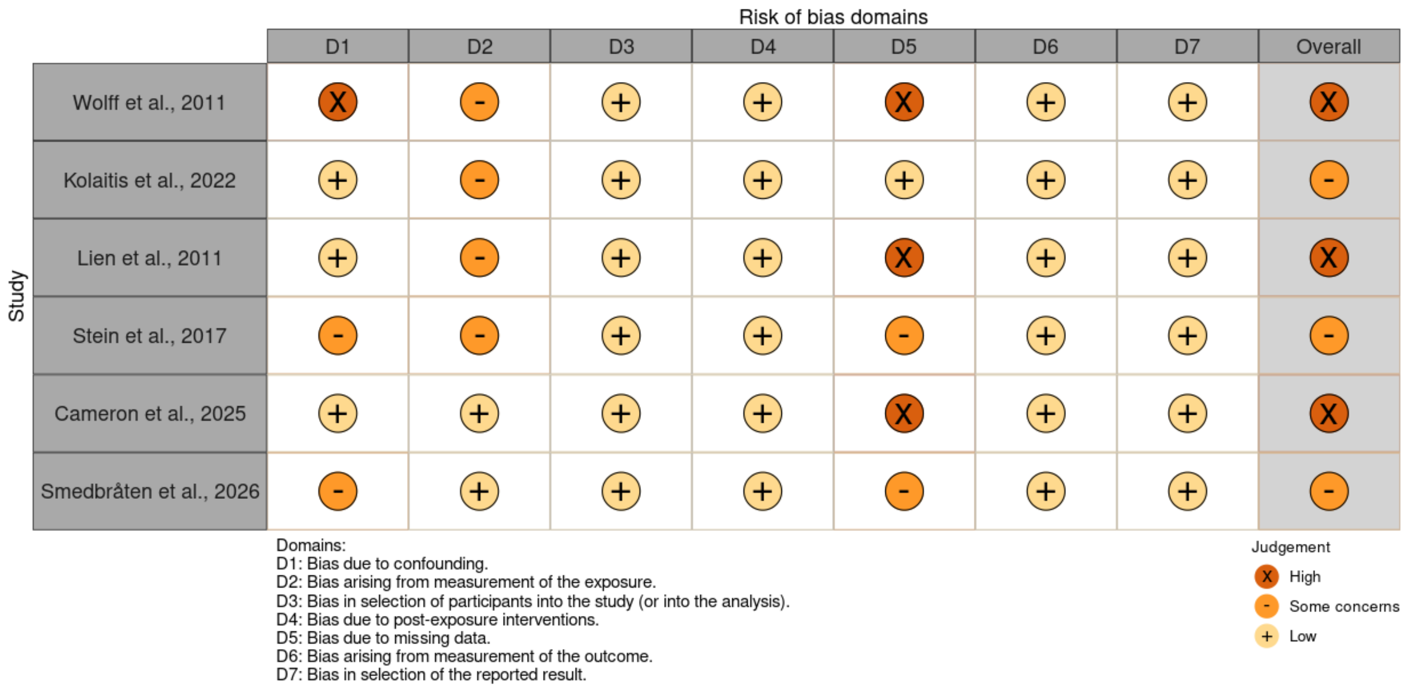

### 6. GRADE rating criteria

As recommended by ROBINS-E research group (Higgins et al., 2024), all outcomes will start out with high quality of evidence. Quality of evidence will be rated down using the following criteria, as per Cochrane guidelines (Cochrane, Chapter 14):

1. Risk of bias: Rate down one (serious limitation) if most information was from studies with some concerns. Rate down two (very serious limitation) if most information was from studies with high or very high risk of bias. Where the amount of information from studies with low risk and high/very risk of bias, rate down one (serious limitation) if there was information from studies with some concerns.
2. Inconsistency: Rate down one for each of these:
    a. Point estimates varied widely across studies, in which that confidence intervals (CIs) show minimal or no overlap, especially when the CIs are in opposite sides of null results.
    b. Considerable between-study heterogeneity ($I^2_{between}$ > 75%).
3. Indirectness of evidence: All studies met this criterion because no indirect comparisons were allowed based on the eligibility criteria.
4. Imprecision of results*:* Rate down one if the CIs are considered wide and suggest different meaningful clinical decisions should the boundaries represent the true effect.
5. Publication bias: Rate down one if publication bias assessments indicate bias. This occurs when contour-enhanced funnel plot indicate small-study effects and Peter's test is significant. Three studies (Tu et al., Osborne et al., Lu et al., 2013) reported that some authors received commercial funding

for their works; however, given that the longitudinal studies were not designed to test a commercially motivated hypothesis, the relevance of this to publication bias risk was judged to be minimal.

Quality of evidence will be rated up one if the effects were deemed to be relatively large (OR > 2.0 or standardised coefficient > 0.5) and the confidence interval was appreciably within the large effect thresholds.

**Table S15.**

*GRADE rating results*

| **Outcome** | **Type of outcome** | **Effect estimate (95% CI)** | **No. of studies** | **Quality Assessment** | | | | | | |
|---|---|---|---|---|---|---|---|---|---|---|
| | | | | **Risk of bias** | **Inconsistency** | **Indirectness** | **Imprecision** | **Publication bias** | **Large effect** | **Overall rating** |
| Anxiety with subsequent pain | Unadjusted association | OR = 1.60; 95% CI: 1.19-2.15 | 9 | Serious | Serious | NA | Not serious | Not serious | No large effect | Low |
| | Adjusted association (binary) | OR = 1.08; 95% CI: 0.94-1.23 | 4 | Very serious | Serious | NA | Serious | Not serious | No large effect | Very low |

| Outcome | Type of outcome | Effect estimate (95% CI) | No. of studies | Quality Assessment | | | | | | |
|---|---|---|---|---|---|---|---|---|---|---|
| | | | | Risk of bias | Inconsistency | Indirectness | Imprecision | Publication bias | Large effect | Overall rating |
| Depression with subsequent pain | Unadjusted association | OR = 1.88; 95% CI: 1.26-2.82 | 11 | Not serious | Serious | NA | Not serious | Not serious | No large effect | Moderate |
| | Adjusted association (continuous) | β = 0.13; 95% CI: 0.01-0.26 | 4 | Not serious | Not serious | NA | Not serious | Not serious | No large effect | High |
| | Adjusted association (binary) | OR = 1.31; 95% CI: 1.01-1.71 | 6 | Not serious | Serious | NA | Serious | Not serious | No large effect | Low |
| Anxiety and depression combined | Unadjusted association | OR = 1.32; 95% CI: 1.15-1.51 | 7 | Very serious | Not serious | NA | Not serious | Not serious | No large effect | Low |

| Outcome | Type of outcome | Effect estimate (95% CI) | No. of studies | Quality Assessment | | | | | | |
|---|---|---|---|---|---|---|---|---|---|---|
| | | | | Risk of bias | Inconsistency | Indirectness | Imprecision | Publication bias | Large effect | Overall rating |
| with subsequent pain | Adjusted association | OR = 1.15; 95% CI: 0.99-1.33 | 8 | Very serious | Serious | NA | Serious | Not serious | No large effect | Very low |
| Pain with subsequent anxiety | Unadjusted association | OR = 2.78, 95% CI: 2.46-3.13 | 4 | Very serious | Not serious | NA | Not serious | Not serious | Large effect | Moderate |
| | Adjusted association | OR = 1.64; 95% CI: 1.11-2.43 | 5 | Serious | Serious | NA | Not serious | Not serious | No large effect | Low |
| Pain with subsequent depression | Unadjusted association | OR = 2.48; 95% CI: 1.55-3.98 | 5 | Not serious | Very serious | NA | Not serious | Not serious | Large effect | Moderate |

| Outcome | Type of outcome | Effect estimate (95% CI) | No. of studies | Quality Assessment | | | | | | |
|---|---|---|---|---|---|---|---|---|---|---|
| | | | | Risk of bias | Inconsistency | Indirectness | Imprecision | Publication bias | Large effect | Overall rating |
| | Adjusted association (continuous) | β = 0.07; 95% CI: 0.03-0.11 | 2 | Not serious | Not serious | NA | Not serious | Not serious | No large effect | High |
| | Adjusted association (binary) | OR = 1.34; 95% CI: 1.12-1.61 | 6 | Serious | Not serious | NA | Not serious | Not serious | No large effect | Moderate |
| Pain with subsequent anxiety and depression combined | Unadjusted association | OR = 1.89; 95% CI: 1.46-2.45 | 4 | Very serious | Serious | NA | Not serious | Not serious | No large effect | Very low |
| | Adjusted association | OR = 1.80; 95% CI: 1.09-3.00 | 3 | Very serious | Serious | NA | Serious | Not serious | No large effect | Very low |
